# Targeted Hybridization Capture of SARS-CoV-2 and Metagenomics Enables Genetic Variant Discovery and Nasal Microbiome Insights

**DOI:** 10.1101/2021.03.16.21252988

**Authors:** Dorottya Nagy-Szakal, Mara Couto-Rodriguez, Heather L. Wells, Joseph Barrows, Marilyne Debieu, Kristin Butcher, Siyuan Chen, Agnes Berki, Courteny Hager, Robert J. Boorstein, Mariah K. Taylor, Colleen B. Jonsson, Christopher E. Mason, Niamh B. O’Hara

**Affiliations:** Biotia, Inc., New York, NY, USA; SUNY Downstate Health Sciences University, The Department Cell Biology/College of Medicine, New York, NY, USA; Twist Bioscience, South San Francisco, CA, USA; Caldwell University, The School of Natural Sciences, College of Natural, Behavioral and Health Sciences, Caldwell, NJ, USA; Lenco Diagnostic Laboratories, Inc., New York, NY, USA; The University of Tennessee Health Science Center, Memphis, TN, USA; Tri-Institutional Computational Biology & Medicine Program, Weill Cornell Medicine of Cornell University, New York, NY, USA; The HRH Prince Alwaleed Bin Talal Bin Abdulaziz Alsaud Institute for Computational Biomedicine, Weill Cornell Medicine, New York, NY, USA; The WorldQuant Initiative for Quantitative Prediction, Weill Cornell Medicine, New York, NY, USA; The Feil Family Brain and Mind Research Institute, Weill Cornell Medicine, New York, NY, USA; Jacobs Technion-Cornell Institute, Cornell Tech, New York, NY, USA

## Abstract

The emergence of novel SARS-CoV-2 genetic variants that may alter viral fitness highlights the urgency of widespread next-generation sequencing (NGS) surveillance. To profile genetic variants, we developed and clinically validated a hybridization capture SARS-CoV-2 NGS assay, integrating novel methods for panel design using dsDNA biotin-labeled probes, and built accompanying software. The positive and negative percent agreement were defined in comparison to an orthogonal RT-PCR assay (PPA and NPA: both 96.7%). The limit of detection was established to be 800 copies/ml with an average fold-enrichment of 46,791x. We identified novel 107 mutations, including 24 in the functionally-important spike protein. Further, we profiled the full nasopharyngeal microbiome using metagenomics and found overrepresentation of 7 taxa and macrolide resistance in SARS-CoV-2-positive patients. This hybrid capture NGS assay, coupled with optimized software, is a powerful approach to detect and comprehensively map SARS-CoV-2 genetic variants for tracking viral evolution and guiding vaccine updates.

**TEASER:** This is the first target hybridization capture-based NGS assay to detect SARS-CoV-2 genetic variants for tracking viral evolution.

## INTRODUCTION

The Coronavirus Disease 2019 (COVID-19) pandemic has resulted in an unprecedented disruption of life across the globe, and an unparalleled death toll, currently reported at over 2M worldwide and climbing (1, 2). To guide public and personal health decisions and control the pandemic, a number of tests for severe acute respiratory syndrome coronavirus 2 (SARS-CoV-2), the virus that causes COVID-19, have been developed and implemented globally (3). Currently, as a rapid and relatively inexpensive technique, real-time reverse transcription-polymerase chain reaction (RT-PCR) is the most commonly used method for the detection of SARS-CoV-2 infection. RT-PCR uses primers to target and amplify one to three regions of the viral genome to determine infection status. Despite the convenience they provide, RT-PCR-based methods do not characterize the virus beyond its presence or absence, and moreover, they are prone to failure when viruses acquire mutations during antigenic shift, since mutations at the primers and/or probe-binding sites may result in false-negative outcomes (4). As underscored by reports of functional genetic variants emerging (including the B.1.1.7 lineage discovered in the UK, B.1.351 lineage discovered in South Africa, and the P.1 variant in Brazil), there is a vital need for tests that tolerate mutations and which are also capable of characterizing the viral genome, including genetic variants, to monitor viral spread and evolution over time.

As an alternative to PCR-based approaches, sequencing-based approaches have increasingly been used to detect the SARS-CoV-2 virus and characterize its genomic variation (5). Sequencing-based approaches include direct metagenomic sequencing, or targeted enrichment such as a hybridization capture-based approach and PCR amplicon-based approach. Direct metagenomic sequencing does not require prior knowledge of the genomic sequence, but has limited sensitivity due to the small viral genome, low abundance of the virus of interest, and the presence of high background from the human host and microbes. Without deep sequencing, which is costly, shotgun sequencing approaches may miss genetic variants due to low or variegated coverage across the viral genome. A targeted approach, which enriches for specific sequences out of a mixed genomic sample, overcomes these difficulties, improving the sensitivity and specificity of NGS-based efforts and achieving higher coverage across the genome (6). The widely used PCR amplicon-based approach [*e.g.,* ARTIC (7)] uses many pairs of overlapping PCR primers to amplify viral genome prior to sequencing. However, it has limited tolerance of mismatch between the targeted sequences and the primers that are used, resulting in an increased risk of amplicon failure as the virus continues to evolve over time. Unlike the PCR amplicon-based approach, the hybrid capture-based method relies on hybridization between the target genome and relatively long probes (up to 120bp) that are designed to be complementary to the target genome. The method can tolerate large target sequence differences from the probe sequences of ∼10% or more (8, 9). The higher tolerance to mismatches makes the successful enrichment of highly divergent SARS-CoV-2 sequences possible (10).

Since the beginning of the COVID-19 pandemic, an unprecedented number of samples have been sequenced by researchers internationally and deposited in public databases. The GISAID consortium has built one of the leading databases (over 400,000 submissions as of January 2021), which has been used to inform the response to the pandemic, vaccine development, and surveillance of novel genetic variants (11). Additionally, the SARS-CoV-2 Sequencing for Public Health Emergency Response, Epidemiology and Surveillance (SPHERES) coalition, another publicly available database, has been sharing genomic sequencing data from over 160 institutions (12). However, these databases have been built using mostly amplicon-based NGS, which may have resulted in missed variants from primer failures or when sufficient sequencing depth was not met. Currently, only around 50,000 full-genome sequences from the United States have been reported, while the United Kingdom has uploaded over 125,000 full-genome sequences, thus giving them an advantage for better surveillance of novel variants. According to the Johns Hopkins COVID-19 database, as of January 2021, the US currently leads the number of COVID-19 positive cases worldwide (25 million cases) but has roughly sequenced only 0.3% of its reported infections, while currently reporting an alarming number of daily cases (∼200,000) (2). There is clearly a great need for the implementation of genomic tools to generate robust and accurate sequence data to assess circulating variants in order to assist in potential challenges that could impact disease outcomes, transmissibility, traceability of the virus, and mitigation efforts.

It is also of interest to ascertain if infection by SARS-CoV-2 may be associated with an altered respiratory tract microbiome and influence disease outcome, or if patients with specific microbial profiles may be more susceptible to SARS-CoV-2 infection (13). Overrepresentation of not only respiratory pathogens, but also opportunistic respiratory microbes (such as *Prevotella spp*.), could affect the inflammatory response and enhance the disease severity (14). Understanding the microbiome of the respiratory tract, potential pathogens, virulence factors, and antimicrobial resistance in SARS-CoV-2 positive patients may elucidate the pathogenicity of COVID-19 in relation to the microbiome and other respiratory pathogens. Characterization of the role and rise of antimicrobial resistance is additionally underscored given the dramatic increase in antibiotics treating pneumonia during the pandemic.

Here, we developed and clinically validated the SARS-CoV-2 NGS Assay and COVID-DX software, a highly sensitive hybrid capture NGS-based assay used for targeting and amplifying the viral sequence for detection of the SARS-CoV-2 viral genome and characterization of genetic variants (Figure 1). Complete genetic variant detection is essential as the SARS-CoV-2 virus continues to evolve and as countries embark on a global vaccination program imparting strong directional selective pressures on the virus, potentially necessitating altered public health responses, and frequent efficacy testing of therapeutics and vaccines. Additionally, we profiled the nasal microbiome of SARS-CoV-2 positive and negative samples. Sequence-based profiling and elucidation of the nasal microbiome allow for a greater understanding of co-infections and drug resistance that contribute to the morbidity and mortality of SARS-CoV-2.

**Fig. 1.**
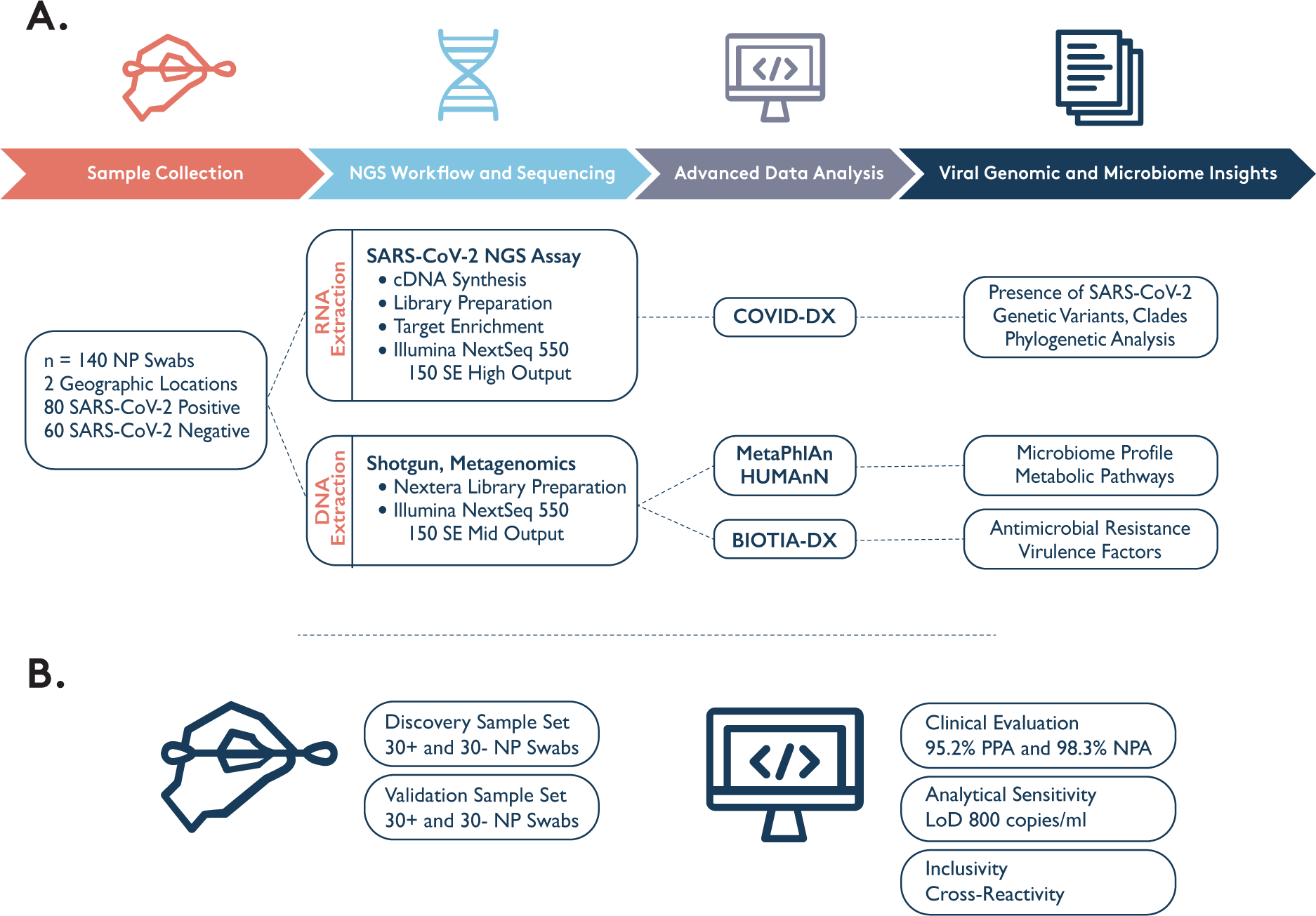
Schematic description of the study workflow (A) and validation (B).

## RESULTS

### Clinical Performance and Analytical Sensitivity

Clinical performance of the SARS-CoV-2 NGS Assay was evaluated by comparing results to an RT-PCR assay approved by the U.S. Food and Drug Administration (FDA) through an Emergency Use Authorization (EUA) authority. A total of 60 clinical NP specimens were tested, including 30 SARS-CoV-2-positive (CVP) and 30 SARS-CoV-2-negative (CVN) specimens. The positive and negative percent agreement (PPA, NPA) was calculated in relation to the RT-PCR comparator method and resulted in a 96.7% PPA and 96.7% NPA. Additional independent clinical validation combined with the original study (120 clinical NP specimens, including 60 CVP and 60 CVN) resulted in a 95.2% PPA and 98.3% NPA (**Table S1 and S2**). Subsampling to 500,000 reads per sample did not significantly change fold enrichment, on-target reads, and presence calling of SARS-CoV-2 viral genome (**Table S2**).

We ran an additional RT-PCR to determine the CT (cycle threshold) values for the CVP specimens using the GenArraytion SARS-CoV-2 RT-PCR assay, which yielded CT values for 44 of these positive samples (validation and independent validation study). The RT-PCR data showed that our samples captured a wide range of viral load, including 22.7% (10/44) at 30.0-35.5 CT, 34% (15/44) at 25.0-30.0 CT, 29.5% (13/44) at 20.0-25.0 CT, and finally 13.6% (6/44) at 15.0-20.0 CT value range (**Table S3**).

The analytical sensitivity established the lowest concentration of SARS-CoV-2 viral genome that can be detected by our assay at least 95% of the time. The limit of detection (LoD) was determined to be 800 copies/ml (0.8 copies/1L) using 30 replicates, which is better than the LoD of several dozen other FDA EUA assays (15). In total, 29/30 (96.67%) known positive samples were successfully identified (**Table S4**). The preliminary 2-fold and 10-fold dilution results using synthetic controls can be found in **Table S4**.

Our hybrid capture technology yielded an average of 43% on-target reads (ranging from 0.005% to 99.5%), and an average fold enrichment of 46,791x (ranging from 5.9x to 108,602x) based on the alignment statistics (HS metrics). The enrichment efficiency was calculated using the number of reads mapping to targets over the total number of reads in the sample. Figure 2 shows the fold enrichment, depth of coverage and and percent coverage of the genome at different depths in relation to the Ct values.

**Fig. 2.**
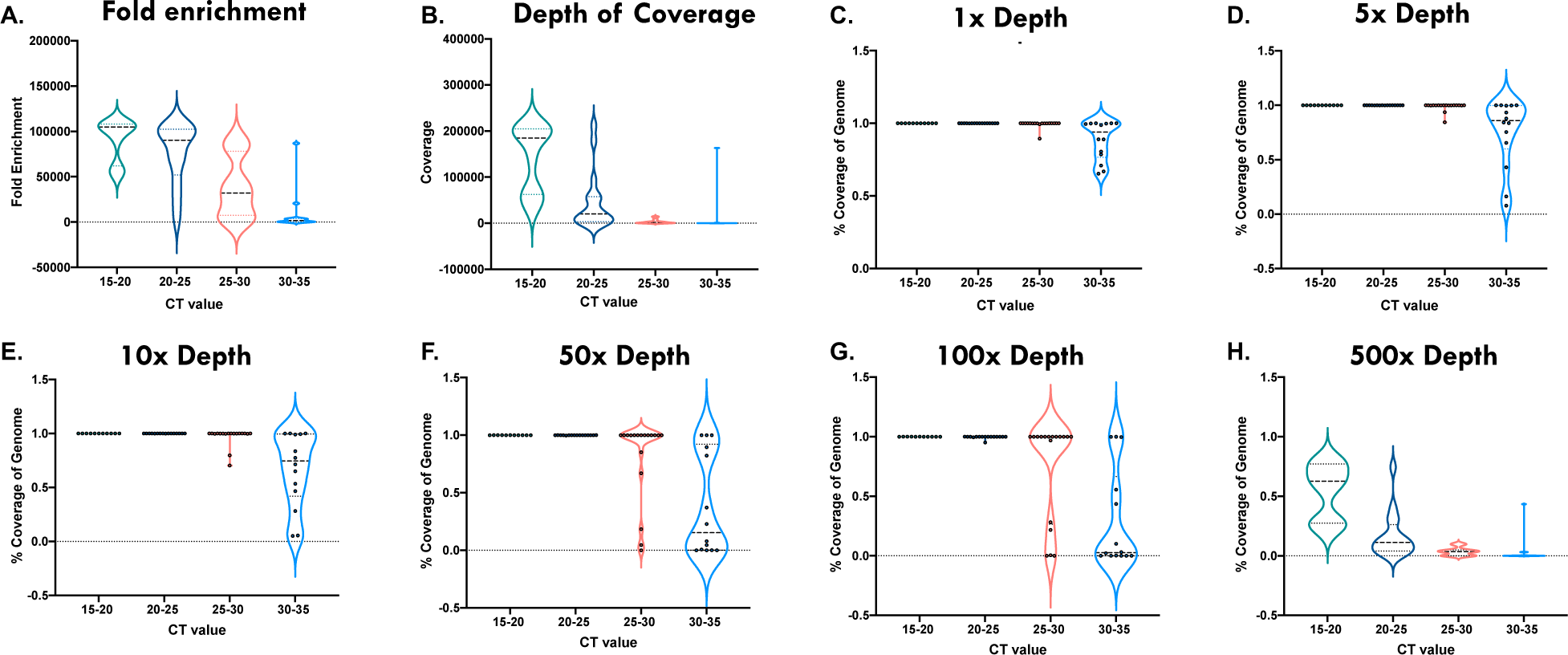
SARS-CoV-2 hybrid capture consistently provides ample fold enrichment, depth of coverage and percent coverage of genome at various depths in samples with Ct values between 15-30. Violin plots depicting the sample distribution of fold enrichment (A), depth of coverage (B), percent coverage of the genome at 1x depth (C), 5x depth (D), 10x depth (E), 50x depth (F), 100x depth (G) and 500x depth (H) in different Ct value groups. Quartiles are depicted in a dotted line pattern (…) and median is shown as dashed line pattern. Ct values of 15-20 are shown in teal (n=11), 20.5-25 in dark blue (n=16), 25.5-30 in salmon (n=18) and 30.5 to 35 in light blue (n=14).

### Inclusivity and Exclusivity Studies

First, our inclusivity analysis, evaluating the assay’s ability to capture different SARS-CoV-2 lineages, was performed on six synthetic control samples. All known variants in the controls were successfully called by the pipeline. We extended our validation for the novel UK B.1.1.7_710528, UK B.1.1.7_601443 and South African EPI_ISL_678597 strain. The known variants have also been successfully called by the pipeline. Then, we performed an extensive *in-silico* inclusivity analysis using the GISAID database. Our analysis identified 16,503 unique sequences in the GISAID database with high identity matches (**Table S5**). Percent identity is defined as the percent of nucleotides in the high scoring pair (the matching section of DNA) that match the query sequence (the probe). We identified 6,200 sequences with 100% mean identity (0 mean mismatches) to our probes, 16,503 sequences with >= 80% mean percent identity to our probes, and 0 sequences with less than 80% mean identity to our probes. Given the high identity for the majority of probes in the majority of sequences, we expect to have a high probability of identifying the presence or absence of most known strains of SARS-CoV-2.

To account for potential cross-reactivity of the probe sequences found in the SARS-CoV-2 Research Panel, we aligned reads to 29 microbial genomes and the human genome along with the SARS-CoV-2 genome (**Table S5**). All pathogens were determined to have no cross-reactivity (no more than 80% homology) with the primers used, except human coronavirus HKU1 with 3 out of 994 probes with 84.6% homology. When we performed *in-silico* exclusivity analysis using approximately 3.6 million viral nucleotide sequences from NCBI Virus, we observed no cross-reactivity (no more than 80% homology, **Table S5**) with the exception of high homology regions in some closely conserved viruses such as bat coronavirus, infectious bronchitis virus, and transmissible gastroenteritis virus strains.

### Novel Genetic Variants Were Detected Using the Hybrid Capture NGS-Based Approach

We used our SARS-CoV-2 NGS Assay to define all genetic variants detected and to match with the globally available database (GISAID, 11-November-2020). After validation of our assay, we defined 71 CVP samples (validation, independent validation, and geographic study samples with positive NGS finding) to undergo NGS analysis to search for genetic mutations. In our 71 CVP samples, we detected 733 mutations (261 different mutations) at 258 different mutation sites (**Table S6**). The most frequently detected variants were the C241T (extragenic), A23403G (D614G), C14408T (P4715L), C3037Y (F924F) and G25563T (Q57H) mutations detected in over 60 samples respectively (Table 1, Figure 3**, Panel A-B**). The majority of the mutations (66%, 173/261) were located in the ORF1ab gene and 14% (37/261) in the S gene (spike protein). Altogether, we found 178 non-synonymous mutations and 14 frameshifts after synonymous mutations; out of those, 58 were radical mutations (based on the Grantham scoring system).

**Fig. 3.**
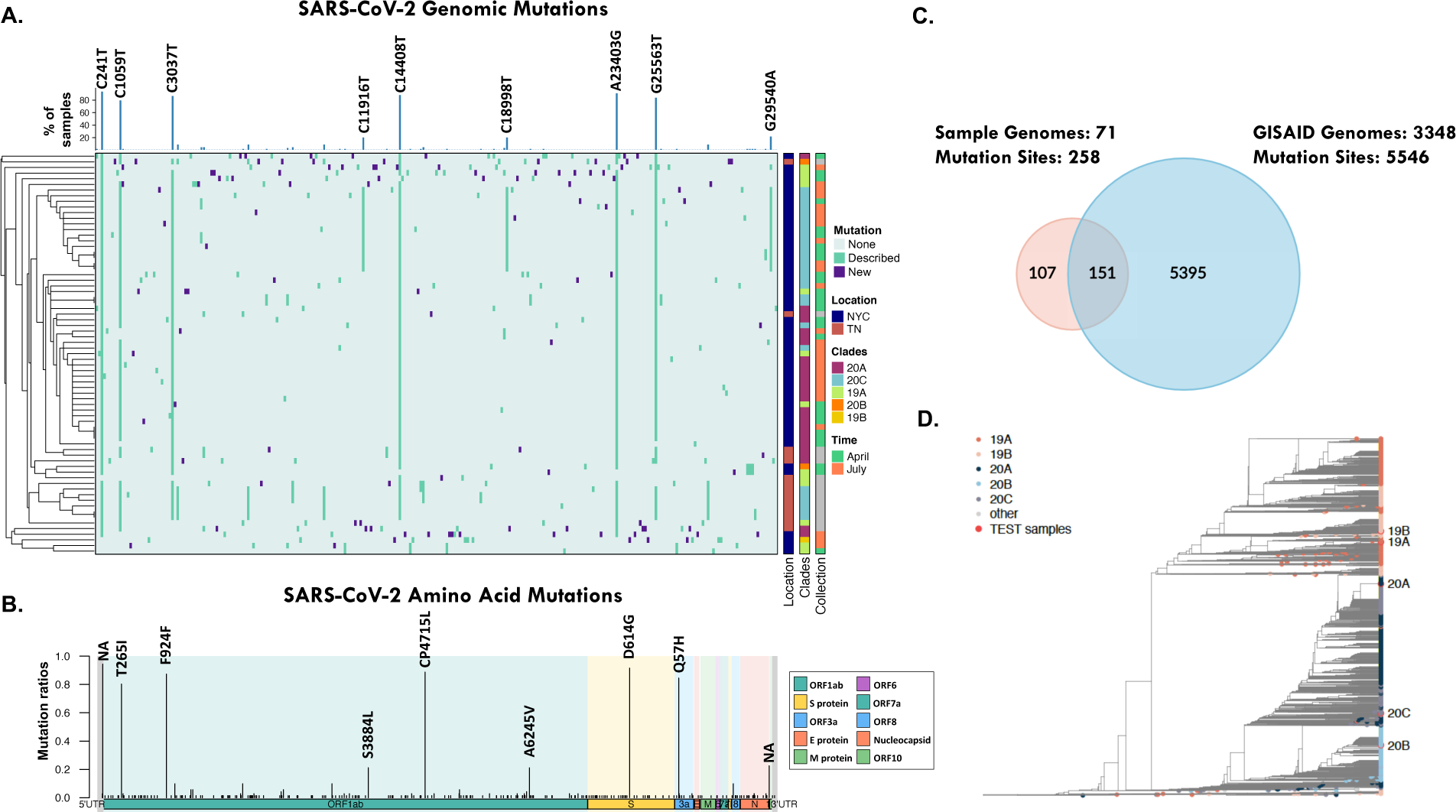
SARS-CoV-2 hybrid capture panel identified novel genetic variants. (A) Heatmap depiction of the nucleotide mutations detected within our SARS-CoV-2 cohort (n = 71). Purple indicates a new mutation, dark green a previously described mutation and light green, no mutation when compared to the SARS-CoV-2 Wuhan strain. Top bar plot annotations indicate the percent of samples containing nucleotide substitutions and those labeled are the most prevalent. Annotations to the right of the heatmap indicate the geographic location, NYC (dark blue), TN (dark orange); phylogenetic clades 20A (purple), 20C (light blue), 19A (lime green), 20B (orange), 19B (yellow); collection date, April 2020 (green), July 2020 (orange). (B) Schematic representation of the amino acid mutation nomenclature of the variants detected in our cohort. (C) Venn diagram displaying the overlap of the mutations detected in our cohort and the mutations reported on GISAID as of 11/11/20. (D) Phylogenetic analysis using SARS-CoV-2 reference genome to determine to which clade in our database test samples are most closely related.

**Table 1.** The list of the most frequent mutations in all samples and those unique for each gepographic locations. (AA: aminoacid, Alt: alteration; Geo: geographic location; NA: not applicable; non-S: non-synonymous; NY: New York; S: synonymous; TN: Tennessee)

| Nucleotide Mutation | AA Mutation | Geo | Frequency in Samples | GISAID Events | Gene | Protein Name | S | AA Alt Properties |
| --- | --- | --- | --- | --- | --- | --- | --- | --- |
| **Most Frequent Mutations in All Samples** |  |  |  |  |  |  |  |  |
| C241T | extragenic | All | 67 | 7 | 5'UTR | NA | NA | NA |
| A23403G | D614G | All | 65 | 6 | S gene | Spike | non-S | radical |
| C14408T | P4715L | All | 63 | 8 | ORF1ab | NSP12/RNApol | non-S | radical |
| C3037T | F924F | All | 62 | 10 | ORF1ab | NSP3 | S | NA |
| G25563T | Q57H | All | 60 | 9 | ORF3a | ORF3a Protein | non-S | conservative |
| C1059T | T265I | All | 57 | 13 | ORF1ab | NSP2 | non-S | conservative |
| G29540A | extragenic | All | 16 | 2 | N/ORF10 Linking Region | NA | NA | NA |
| C11916T | S3884L | All | 15 | 4 | ORF1ab | NSP7/Replicase | non-S | radical |
| C18998T | A6245V | All | 15 | 1 | ORF1ab | NSP14/NSP11 | non-S | conservative |
| **Unique Mutations for Each Geographic Location** |  |  |  |  |  |  |  |  |
| G29540A | extragenic | NY | 16 | 2 | N/ORF10 Linking Region | NA | NA | NA |
| C11916T | S3884L | NY | 15 | 4 | ORF1ab | NSP7/Replicase | non-S | radical |
| C18998T | A6245V | NY | 15 | 1 | ORF1ab | NSP14/NSP11 | non-S | conservative |
| C4113T | A1283V | NY | 4 | 6 | ORF1ab | NSP3 | non-S | conservative |
| C27964T | S24L | TN | 7 | 7 | ORF8 | ORF8 Protein | non-S | radical |
| C3411T | A1049V | TN | 7 | 3 | ORF1ab | NSP3 | non-S | conservative |
| T6394C | D2043D | TN | 7 | 3 | ORF1ab | NSP3 | S | NA |
| C10319T | L3352F | TN | 7 | 4 | ORF1ab | NSP5 | non-S | conservative |
| A4197G | E1311G | TN | 4 | 2 | ORF1ab | NSP3 | non-S | radical |
| G8179A | R2638R | TN | 4 | 4 | ORF1ab | NSP3 | S | NA |
| C15924T | Y5220Y | TN | 4 | 1 | ORF1ab | NSP12/RNApol | S | NA |

We identified 107 new mutations that have not been previously described in GISAID (Figure 3**, Panel C**), with the majority located in the ORF1ab gene (71/107, 66%) and the S gene (24/107, 22%). We detected 81 non-synonymous mutations and 12 frameshifts after synonymous mutations (3 in the spike protein); out of those, 29 were radical mutations (based on the Grantham scoring system).

Samples collected from two geographic locations (NY, TN) shared several similar genetic variants. Unique variants identified in the NY samples included: G29540A (extragenic), C11916T (S3884L), C18998T (A6245V), and C4113T (A1283V) (Table 1). Unique mutations in the TN samples are C27964T (S24L), C3411T (A1049V), T6394C (D2043D), C10319T (L3352F), A4197G (E1311G), G8179A (R2638R) and C15924T (Y5220Y) (Table 1). Most of the unique variants were located in ORF1ab.

To confirm the ability of the capture assay to accurately call novel variants, we validated 6 genetic variants identified in our cohort using PCR amplification followed by Sanger sequencing (**Table S7A**). The genetic regions were PCR amplified using specifically designed primers followed by Sanger sequencing. The validated mutations located in the ORF1ab and S genes contained non-synonymous/synonymous, radical/conservative mutations with various read depth (∼10-400). Additionally, 18 genetic variants we identified as missing from GISAID in October, were later added to GISAID by our November audit, further validating the accuracy of our variant calling approach.

### Phylogenetic and Geographic Origin (Clades) Results

We next performed a phylogenetic analysis to provide additional information on the likely origin and diversity of the SARS-CoV-2 viral genome based on genetic variation in the clinical samples. In our CVP samples, we identified clades 19A, 20A, and 20C as being the most abundant clades (n=13, 27, and 28, respectively, **Table S2**), which matches the results seen from NY cohorts (5). Figure 3**, Panel D** represents our database of 3,365 samples (GISAID, 23-April-2020) and their phylogenetic clustering with 5 clinical test samples representing each clade.

### COVID-19 Status Associated with an Altered Metagenomic Profile

To provide a more in-depth profile of other microbial species in our samples, 106 metagenomic libraries were prepared with Illumina’s Nextera Flex Kit, sequenced, and subsequently processed with MetaPhlAn2. We removed samples with 100% human reads leading to 22 CVN and 26 CVP samples that were further analyzed for their microbiome composition, AMR profiles, and virulence factors.

We detected a total of 66 bacterial genera and 191 bacterial species within our cohort. Overall, the microbiome profiles identified in our cohort reflect a normal, commensal nasopharyngeal microbiome, spanning *Streptococcus, Veillonella, Prevotella, Rothia, Actinomyces, Haemophilus, and Neisseria species* (Figure 4). There was no clustering observed based on COVID-19 status. Additionally, 7 other viruses relevant to human health were detected in the samples, including human herpesvirus 4, influenza A, and human adenovirus D (Figure 4).

**Fig. 4.**
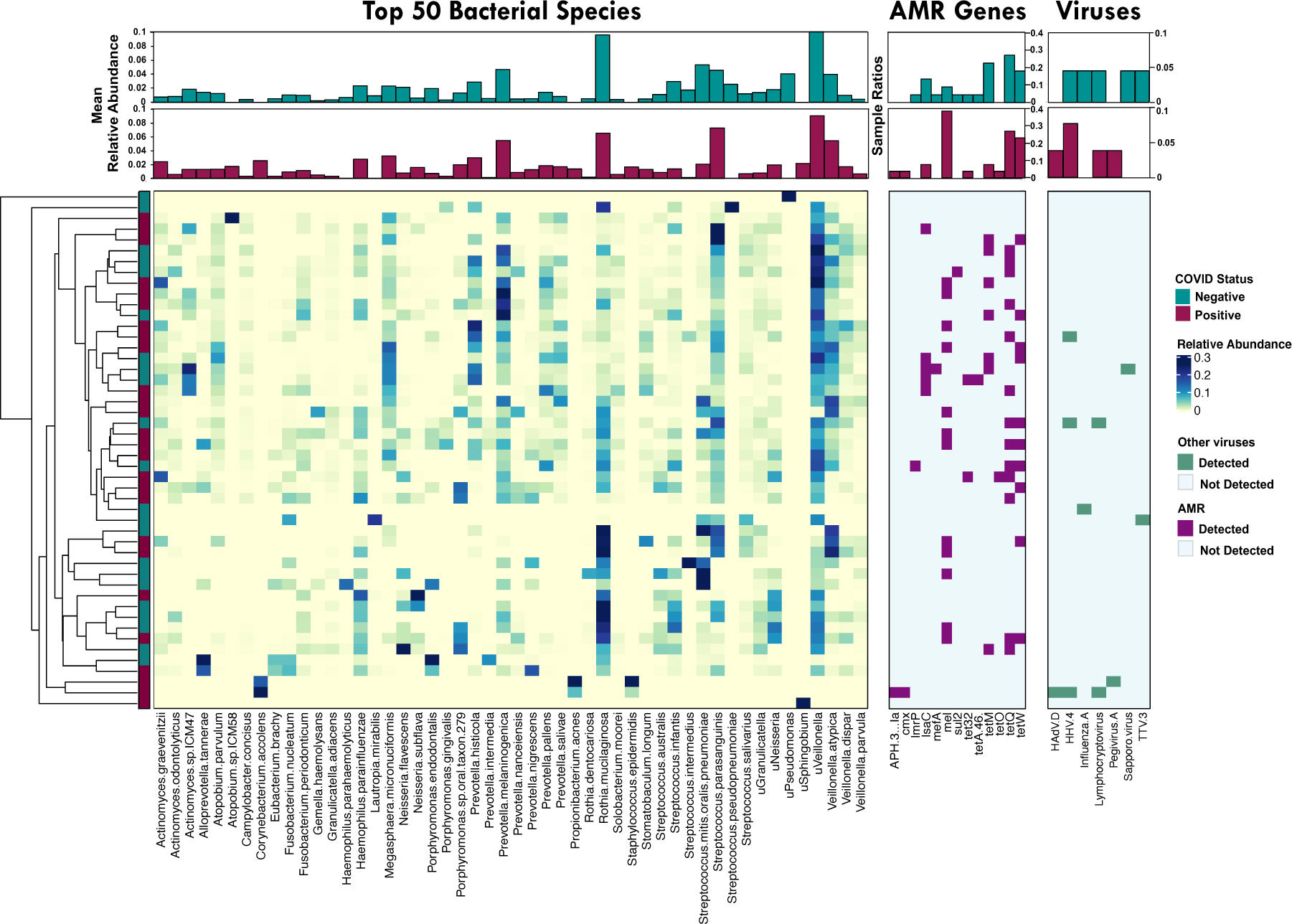
Nasopharyngeal microbiome profiles. Main heatmap depicts the relative abundance of the top 50 most abundant bacterial species found in our cohort. Top bar plot annotations indicate the mean relative abundances per taxa based on COVID-Status, CVP (maroon, n=26), CVN (teal, n=22). Annotation heatmaps display the presence (purple) or absence (light blue) of AMR genes and DNA viruses (green) detected in our samples. Top bar plot annotation show the sample ratios.

We next examined sample richness and clustering or differences between the various species. Alpha diversity metrics showed no significant difference in richness (Observed Species and Shannon Index) or evenness (Pielou’s) between CVP and CVN samples; however, CVN samples exhibited a trend of lower Observed Species and Shannon Index (**Figure S1, Panel A**). Based on the Bray Curtis PCoA analysis (**Figure S1, Panel B**), 4 clusters and 5 outliers were observed, of which cluster A and cluster C are largely dominated by *Streptococcus* (33.1/20.4% relative abundance), *Prevotella* (8.8/12.2% relative abundance), *Veillonella* (7.1/18.5% relative abundance), *Rothia* species (8.3/15.9% relative abundance), while cluster B and cluster D *Prevotella* (19.4/21.1% relative abundance), *Veillonella* (21.7/20.7% relative abundance), *Streptococcus* (15.6/10.8% relative abundance), *Megasphaera* dominant. Cluster C *Streptococcus*/*Veillonella* dominant and cluster D *Prevotella*/*Veillonella* dominant contain 69.2% of the CVP samples within our cohort. Of note, one of the outliers was a CVN sample with 100% of the reads mapping to influenza A virus. Bray Curtis Dissimilarity Index indicated that *Rothia mucilaginosa* is the main driver of clustering, which is significantly differentiated by the relative abundance levels found by LEfSe and Mann-Whitney U-test. Moreover, another driver of clustering is explained by the presence/absence of *Streptococcus parasanguinis* (n=18, 12 CVN samples and 6 CVP samples) and unclassified *Neisseria (*n=26, 8 CVN samples and 18 CVP samples), respectively, also indicated by LEfSe and Mann-Whitney U-test (data not shown).

Although COVID-status did not show clear differences in the nasopharyngeal microbiome communities between CVP and CVN samples, Linear Discriminant Analysis Effect Size (LEfSe) revealed 6 bacterial species and 1 genus (*Actinomyces*) that are significantly increased in CVP samples (**Figure S2, Panel A**), of which 2 species-level taxa, *Actinomyces graevenitzii and Prevotella salivae,* were further confirmed to be significant when comparing their relative abundances using a Mann-Whitney U-test (**Figure S2, Panel B**), while *Megasphaera micronuciformis*, *Veillonella dispar,* and *Streptococcus gordonii* only exhibited a trend of higher relative abundance in CVP samples.

Further, we compared the classification obtained from MetaPhlAn2 with our k-mer based approach pipeline (BIOTIA-DX) and found high concordance in the genus level taxonomy. The top 80% most dominant bacterial genera (11 in 66 total) showed high overlap (8/11) between the two metagenomic classification tools. Two out of the 3 discordant genera were identified on the bottom 20% by each classifier, and only 1 taxa (*Candidatus Saccharibacteria)* was not detected by MetaPhlAn2 (**Figure S3, Panel A**).

Since *Prevotella spp*. were found to be a biomarker for CVP status based on the data obtained by two independent bioinformatic pipelines but were found to conflict at the species level classification, we sought to use 16s rDNA sequencing to aid in confirming the phylogeny of the correct *Prevotella* species. Neighbor-joining phylogenetic analysis of the 16s rDNA sequences obtained from *Prevotella* amplicons confirmed that the biomarker increased in CVP samples is most closely related to *Prevotella salivae* (**Figure S3, Panel B**).

### Distinct Metabolic Pathways Linked to SARS-CoV-2 Positivity

In order to assess the metabolic processes that are represented within our cohort, we used HUMAnN2. A total of 434 metabolic pathways were detected in our dataset, with the most prevalent among bacterial species being those associated with nucleotide, amino acid, lipid, and cell wall biosynthesis. We then compared the differences in metabolic pathways based on COVID-19 status and found 4 metabolic pathways to be overrepresented in CVN samples, *L*-homoserine and *L*-methionine biosynthesis, UDP-N-acetylglucosamine biosynthesis, Seleno amino acid biosynthesis (PWY-6936), and S-adenosyl L-methionine biosynthesis superpathway; while 3 pathways, gluconeogenesis (PWY-66-399), flavin biosynthesis (PWY-6168), and L-histidine degradation (PWY-5030) were increased in CVP samples (**Figure S4, Table S8**). One of the pathways observed in the CVN samples (PWY-6936, seleno-amino acid biosynthesis) was present in *Rothia, Streptococcus,* and *Haemophilus species, while* PWY-5030 that was increased in CVP samples was associated with *Streptococcus gordonii.* Further, we explored the functional profiles associated with the bacterial species increased on CVP samples and observed 24 pathways associated with *Actinomyces graevenitzii*, 45 with *Prevotella salivae*, 30 with *Megasphaera micronuciformis*, 28 with *Veillonella dispar,* and 11 with *Atopobium parvulum* **(Table S8)**.

### Altered Antimicrobial Resistance and Virulence Factor Profile in SARS-CoV-2 Positive Specimens

For assessing the AMR genes and virulence factors present in our samples, the k-mers obtained from BIOTIA-DX software were classified using the CARD database and VFDB, respectively. A total of 13 different AMR genes were detected across all samples (CVP = 19, CVN = 12, all specimens had 55 AMR genes altogether) including genes conferring resistance for several drug classes such as tetracycline (*tet32, tetA*(*46*)*, tetM, tetO, tetQ, tetW*), macrolide (*mel, mefA, lmrP*), aminoglycoside (*APH(3’)-la* and *IsaC*), sulfonamide (*sul2*), and phenicol (*cmx*) (Figure 4). Interestingly, *mel*, which confers macrolide resistance, was significantly overrepresented in CVP samples. In order to understand which bacteria may be carrying these AMR genes, we correlated the AMR profiles with the bacterial taxa and found *mel* to be significantly correlated with *Streptococcus spp.* (*Streptococcus parasanguinis, Streptococcus salivarius, Streptococcus oral taxon 431*).

Furthermore, 7 virulence factors were detected in 10 samples, 6 CVN and 4 CVP samples with the serine-rich repeat protein (SRRP) family (SecA2/SecY2 system) being the most prevalent, found in 3 CVN and 3 CVP samples. Other virulence factors observed include neuraminidase, transferrin-binding protein, direct heme uptake system, major surface protease (MSP), SRRP family, and streptolysin S.

## DISCUSSION

### Novel Variant Discovery and Importance

Using our novel SARS-CoV-2 NGS Assay and COVID-DX pipeline, we found a remarkable number of previously unreported genetic variants, with 107 mutations that were not described previously, including 24 in the functionally important spike protein. We believe this was facilitated by our hybrid capture NGS-based approach, which enabled the amplification of the target of interest so that a greater number of sequence reads are generated from the target instead of the host. In this study, we achieved an average enrichment of the virus of 46,791-fold. The result was an average genome coverage of >93% (at 5X) with >99% coverage in 48/60 CVP samples in the validation set. A subset of the tested, novel variants were validated using an orthogonal sequencing approach (Sanger), and through GISAID audits at multiple time points, indicating that hybrid capture NGS-based approach is a powerful method to detect genetic variants in the SARS-CoV-2 virus; such a method could readily be implemented for other viruses as well.

This test was also found to be sensitive and specific with a positive percent agreement of 95.2% [90%-100% CI] and negative percent agreement of 98.3% [95.2%-100% CI] when compared to an RT-PCR assay approved by the FDA for EUA. These results highlight the utility of a hybrid capture NGS-based approach in the detection and characterization of the SARS-CoV-2 viral genome, especially as we see ongoing evidence of the functional evolution of the virus, warranting widespread and affordable characterization of genetic variants. Our variant findings have been further validated by capturing and sequencing a range of synthetic RNA controls, including the novel strains with larger deletions (6nt) and SNPs of public health interest (*e.g.,* B1.1.7). In our clinical cohort, we detected 12 novel frameshifts after synonymous mutations, out of those 3 deletions were found in the S gene.

Characterization of genetic variants identified in the SARS-CoV-2 viral genome is important for researchers studying the epidemiology (*e.g.,* transmission, spread) and evolution (*e.g.,* substitution rate, adaptation) of the virus over time. Although the virus appears to have a low, random substitution rate with an average rate of 1-2 mutations per month (16), any acquired changes could potentially be selected and alter its transmissibility, infectivity, or pathogenicity (17). Mapping variants across the viral genome can also inform drug therapies and guide vaccine development or updates (18). While the majority of mutations have no effect on the fitness of the virus, there is evidence that some variants may make the virus more transmissible (19). Studies are underway to assess if there are any clinical effects of the existing mutations and if the lineages observed may make the virus better at evading the host immune response (17,19,20). A recent study describes the emerging recurrent deletions in the spike protein that are mapped to antibody epitopes, resulting in resistance to neutralizing antibodies (21).

In our cohort, we have found dominant variants that have been previously reported. For instance, the D614G mutation has become the basis of one of the dominant phylogenetic clades of the virus during the pandemic (17). This mutation may have important implications for the transmission since it is located in the S gene (22). Coronavirus mutations in the functionally important spike protein have the potential to affect virus infectivity, pathogenicity, and susceptibility to neutralizing antibodies (23).

With limited sequencing, low coverage sequencing, and/or direct metagenomics, it is possible that variants are missed and so present in populations long before detection. More recently, a number of reports described strains found in the UK (B.1.1.7 strain) (19, 24), South Africa (B.1.351 strain) (25), and Brazil (P.1 strain) (26). Once described and tested for specifically, it was found that the B.1.1.7 strain was present in many other countries outside the UK, indicating that this strain may have been circulating and spreading in the population for some time (27). These novel genetic variants, such as the UK strain, are of recent concern because these mutations have been shown to decrease the efficiency and performance of amplicon-based assays, with some tests showing a “dropout” in which one of the genetic targets is not reported (28). Currently, as of January 2021, three widely used RT-PCR EUA-approved diagnostic tests in the US are being evaluated due to concerns of a possible increase in the false positivity rates of such assays (28). Furthermore, novel variants have the potential to evade the immune response after exposure or infection, with some strains circulating among the population while infection rates are rampant worldwide. Although more costly, sufficient sequence coverage is a prerequisite for accurate and sensitive variant detection and calling. More viral genomic surveillance studies and trials will be necessary to understand how strain variability impacts vaccine efficacy and long-term immunity.

Genetic variant surveillance is a powerful tool for informing epidemiology and understanding the spread and evolution of the virus among communities in order to inform public health mitigation efforts of variant-driven outbreaks. It is becoming evident that the prevalence of the B.1.1.7 strain is driving the spike of infections reported in the UK, and has contributed to the highest positive rate seen worldwide (16%) in Ireland, following resuming a strict 6-week lockdown at the beginning of December (2). The transmissibility of this genetic variant further stresses the importance of initiatives for actively genotyping a percentage of the population testing positive for SARS-CoV-2. However, more studies would be needed to evaluate current mutation rates and what percentage of the population should be assessed for effective public health monitoring. We have identified and validated the variant T6394C (D2042D) specific to our TN cohort but not found in specimens collected in NY. This finding highlights the utility of our pipeline for identifying variants specific to geographic location.

Overall, in line with previous studies, we have found that while the hybrid capture-based workflow is labor-intensive, includes more steps, and requires higher input amounts in comparison to amplicon-based technologies, this approach is effective at targeting large regions of the genome and discovering novel variants. Hybrid capture can work with millions of targets per panel and dozens to hundreds of overlapping capture probes, leading to higher specificity for recovering entire viral genomes and demonstrating higher uniformity of coverage when compared to amplicon-based technologies (10,29,30). The limitations of amplicon-based sequencing have resulted in discordant single nucleotide variants as well as frequently missed variants (10, 31), and capture-based methods represent an important tool for addressing any missing variation. As the COVID-19 pandemic is still ongoing and new variants are emerging, global implementation of robust SARS-CoV-2 sequencing programs is needed to further characterize the viral genome and to inform public health responses (9).

### Profiling the SARS-CoV-2 Microbiome

Only a few studies evaluating the microbiome associated with COVID-19 patients have been conducted to date. Overall, our microbial profiling overlaps with previous studies using shotgun sequencing and metagenomic analysis on nasopharyngeal bacterial composition (32, 33) with commensal respiratory species detected such as *Streptococcus, Veillonella, Prevotella, Rothia, Actinomyces, Haemophilus, and Neisseria species*. While one study found decreased diversity in SARS-CoV-2 positive specimens, reporting the vast majority of reads mapped to a single organism in each sample, which could be explained by the necessity of the higher coverage of Nanopore sequencing (13), we found no significant difference in richness or evenness between CVP and CVN samples; however, CVN samples exhibited a trend of lower richness. In addition, Mostafa *et al.* reported higher abundance of *Corynebacterium accolens* and decreased abundance of *Propionibacteriaceae* in CVP, which are common skin commensals, respectively (13). Although we have not found clear clustering based on SARS-CoV-2 status, our analysis revealed overrepresentation of 7 bacterial taxa (*Actinomyces* genus, *Actinomyces graevenitzii, Prevotella salivae, Megasphaera micronuciformis*, *Veillonella dispar*, *Atopobium parvulum,* and *Streptococcus gordonii*) with a significantly higher abundance in CVP. *Actinomyces graevenitzii and Prevotella salivae* were further confirmed to be significant when comparing their relative abundances in CVP using a Mann-Whitney U-test.

Previous studies have argued that the nasal microbiome composition and dynamics could modulate host immunity and increase susceptibility to upper respiratory infection, particularly acute viral infection (34–36). Interestingly, *Prevotella*, a taxa enriched in CVP patients, has been found to be increased in patients with acute and chronic respiratory diseases such as asthma and influenza (36–38). The altered microbiome and, specifically, the overrepresentation of *Prevotella spp.* may play a role in modifying the host immunity and consequently the host response to viral infections (14). The observed altered species (*Streptococcus, Veillonella, Prevotella, and Rothia spp.*) driving our clusters have been linked to symptomatic and asymptomatic viral respiratory infections in previous studies (39). In concordance with our results, another study on hospitalized COVID-19 patients has reported high prevalence of *Rothia* across patient samples and built environments (40), raising the importance of microbiome profiling in patients with acute viral respiratory infections.

Further, drug resistance profiling is of importance because the COVID-19 pandemic may pose additional infectious disease threats, including a potential rise in antimicrobial resistance. There has been a dramatic increase in use of antibiotics treating pneumonia associated with the viral respiratory infection. One study showed that over 70% of hospitalized COVID-19 patients have received antibiotics throughout their disease course though only 8% demonstrated bacterial or fungal superinfection (41). Additionally, COVID-19 related long-term hospitalization may lead to increased multidrug resistance (MDR) infections and prolonged antibiotic therapy (42). The overrepresentation of macrolide resistance (*melC*) in SARS-CoV-2 positive patients linked to *Streptococcus spp.* may be associated with the underlying viral infection (43). The metabolic profile associated with nucleotide, amino acid, lipid, and cell wall biosynthesis represented by the overrepresentation of different bacterial species may be associated with an altered bacterial adherence, host mucosal immunity, and secondary co-infections, and can be used as infectious biomarkers for risk stratification (44).

### Prospectives

The SARS-CoV-2 NGS Assay can be used as a method for viral detection by looking at data not only at an individual patient level, but on a population level as well. It also may be used to test the environment and determine how the SARS-CoV-2 virus is spreading in communities by sampling wastewater (45). This may be useful to track viral incidence and inform public health decisions on a community level. Due to shortages in COVID-19 diagnostic tests and testing sites, individuals are not always able to undergo regular testing, especially if they are not showing symptoms. Therefore, sampling wastewater can be used as a secondary option to determine how the virus fluctuates in the community and directs public health measurements to limit the spread of the virus.

Hybrid capture NGS-based methods are also a powerful approach to anticipate and characterize emerging human viruses transmitted from wildlife. NGS enables characterization of genes and pathogenicity important in transmission from animals to humans. Importantly, genetic similarity to known pathogens in PCR-targeted regions of the genome is not an accurate predictor of human pathogenicity (46–48). Coronaviruses, in particular, cannot be accurately confirmed or dismissed as human pathogens based on these PCR fragments because of the propensity for coronavirus genomes to recombine, particularly in the spike gene (49). NGS provides a valuable tool for the detection of emerging viruses in wildlife and generates critical data that is needed to characterize the potential for a virus to be pathogenic in humans.

### Limitations and future work

Our study findings were limited by the cross-sectional nature of testing. While the current work highlights the importance of the characterization of the SARS-CoV-2 viral genome and provides valuable insights into the genetic variants and geographic associations of variants, extension of our work is needed by processing additional samples collected from additional geographic regions and with additional clinical metadata that could drive risk stratification linked to novel variants. Additional longitudinal testing of SARS-CoV-2 positive samples using our technology could enable further screening of variants in the same individual and same geographic location. Further optimization is needed to accommodate different sample types (saliva and other respiratory specimens) and to extend our variant curation to support the continuously growing scientific knowledge around genetic variants and their role in viral fitness. While we observed intra-host diversity through visualizing sequence alignments, our variant caller only reports the most frequent nucleotide or variant at each genomic site. Incorporating calling of multiple variants when present at each genome site to provide intra-host diversity could prove useful.

Although our work contributes to the ongoing elucidation of the role of the microbiome and secondary infections commonly associated with COVID-19, further longitudinal studies linking disease outcome to microbial load and dynamics are required for better understanding the virus, as well as microbe and host interactions during the course of the disease. Expanding the cohort and collecting clinical metadata will also be a key next step since it creates an unprecedented opportunity to map the genetic characterization of the virus and patient history (reason of sampling, travel history, co-morbidities, prior medication) with disease outcome, disease progression, and hospitalization risk.

In summary, genomic innovations are transforming epidemiology to better characterize, respond, and prepare for outbreaks. Novel genome-based approaches enable “precision epidemiological response” and public health interventions on both an individual and population-based scale. Viral metagenomics, and the novel hybrid capture NGS-based assay we describe, can provide important genetic insights into SARS-CoV-2 and other emerging pathogens and improve surveillance and early detection, potentially preventing or mitigating new outbreaks. Better understanding of the continuously evolving SARS-CoV-2 viral genome and the impact of genetic variants may provide individual risk stratification, precision therapeutic options, improved molecular diagnostics, and population-based therapeutic solutions. Future work includes the extension of this hybrid capture NGS-based assay to characterize other viral and bacterial genomes and the collection of clinical metadata to define risk stratification and disease pathogenesis in relation to SARS-CoV-2 genetic variants.

## METHODS

### Experimental Design

We developed and clinically validated the SARS-CoV-2 NGS Assay and COVID-DX software, a highly sensitive hybrid capture NGS-based assay used for targeting and amplifying the viral sequence for detection of the SARS-CoV-2 viral genome and characterization of genetic variants (Figure 1). Additionally, we profiled the nasal microbiome of SARS-CoV-2 positive and negative samples.

### Specimen Collection

Upper respiratory tract specimens (nasopharyngeal [NP] swabs) were collected following the current CDC guidelines (50) and the manufacturer’s instructions for specimen collection. Altogether 120 specimens were analyzed from New York (NY) using the SARS-CoV-2 NGS Assay (validation and independent validation study). An additional 20 SARS-CoV-2 positive specimens were analyzed from Tennessee (TN) to define variant differences in samples collected from two geographic locations. **Table S3** contains the specimen collection date, location, collection device, and transport matrix. For all collections, NP swabs were immediately placed into sterile tubes containing 2-3 ml of viral transport media. Samples were stored for up to 24 hours at room temperature, or up to 72 hours when stored at 2°C to 8°C prior to transportation. After 72 hours, all samples were frozen at –70°C or colder until additional testing was performed.

RT-qPCR technology (Panther Fusion SARS-CoV-2 Assay [Hologic, Marlborough, MA]; and cobas ® SARS-CoV-2 test with the Cobas 6800/8800 System [Roche, Basel, Switzerland]) was used to define the presence of SARS-CoV-2 viral RNA (SARS-CoV-2 positive: CVP and SARS-CoV-2 negative: CVN; **Table S3**). These technologies have been issued EUA approval by the FDA for the clinical diagnostics of COVID-19. The Ct value was defined using GenArraytion COVID-19 duplex RT-qPCR [Rockville, MD]) and cobas ® SARS-CoV-2 test with the Cobas 6800/8800 System (Roche, Basel, Switzerland) (**Table S3**). De-identified samples were collected and processed under the IRB numbered Pro00042824 (Advarra).

### RNA and DNA Extraction

RNA and DNA from NP specimens were isolated and purified by manual extraction using Direct-zol DNA/RNA MiniPrep kit (250 µl input volume, spin column, 100 µg binding capacity, Zymo Research, Irvine, CA). Extracted and purified RNA samples were converted to cDNA through random priming using Random Primer 6, ProtoScript II First Strand cDNA Synthesis kit, and NEBNext Ultra II Non-Directional RNA Second Strand Synthesis kit reagents (NEB, Ipswich, MA).

### Library Preparation, Target Enrichment, and Sequencing

The cDNA samples were converted to Illumina TruSeq-compatible libraries using the Twist Library Preparation kit with Unique Dual Indices (Twist Bioscience, South San Francisco, CA). After library generation, 8 uniquely barcoded libraries were pooled (187.5 ng per library) to create an 8-plex hybridization reaction. Hybridization was performed for 2 hours using Twist Fast Hybridization reagents and SARS-CoV-2 Research Panel, a biotin-bound DNA panel that targets libraries containing the SARS-CoV-2 sequence. These libraries were isolated from human libraries using biotin/streptavidin bead chemistry. Beads were washed several times to reduce the number of off-target libraries in the sequencing pool. All enriched library pools were spiked with 1% PhiX and sequenced on an Illumina NextSeq 550 platform using a NextSeq500/550 High Output kit (Illumina, San Diego, CA) set to 150bp single-end reads, which yielded an average of 15.1M and 1.5M reads from positive and negative samples, respectively. Picard Hs Metrics showed an average of 43% on-target reads, ranging from 0.005% to 99.5%. Additionally, our target enrichment approach yielded an average fold enrichment of 46,791x, ranging from 5.9x to 108,602x. Subsampling to 500,000 reads per sample did not significantly change fold enrichment (mean: 108,598x) or on-target reads (mean: 40.3%).

### Inclusivity and Exclusivity Study Design

To evaluate the inclusivity of the assay to capture different SARS-CoV-2 lineages, we processed six synthetic control samples created by Twist Bioscience with known genetic sequences (Twist Bioscience control 1-6, MT007544.1, MN908947.3, LC528232.1, MT106054.1, MT188340, MT118835), including the original Wuhan coronavirus strain, through laboratory processing and the bioinformatics pipeline. The synthetic controls contain the following variants: control 1: T19065C, T22303G, G26144T, ACGATCGAGTG29749A; control 3: TG11082T; control 4: C9924T; control 5: T514C, C17410T; control 6: C8782T, T18603C, T18975A, A19175C, C27925T, T28144C, C29095T. We extended our validation for the novel UK B.1.1.7_710528 (with mutations including C240T, C912T, C3036T, C3266T, C5387A, C5985T, T6953C, 11287delTCTGGTTTT, C14407T, C14675T, C15278T, C15856T, T16175C, A17614G, 21764delTACATG, 21990delTTA, A23062T, C23270A, A23402G, C23603A, C23708T, T24505G, G24913C, T27884C, C27971T, G28047T, A28110G, 28270delA, G28279C, A28280T, T28281A, G28880A, G28881A, G28882C, and C28976T), the UK B.1.1.7_601443 (with mutations including C240T, C912T, C3036T, C3266T, C5387A, C5985T, T6953C, 11287delTCTGGTTTT, C14407T, C14675T, C15278T, T16175C, 21764delTACATG, 21990delTTA, A23062T, C23270A, A23402G, C23603A, C23708T, T24505G, G24913C, C27971T, G28047T, A28110G, G28279C, A28280T, T28281A, G28880A, G28881A, G28882C and C28976T) and South African EPI_ISL_678597 strain (with mutations including G173T, C240T, C1058T, A2691T, C3036T, G5229T, A10322G, 11287delTCTGGTTTT, C14407T, A21800C, A22205G, 22280delCTTTACTTG, G22812T, G23011A, A23062T, A23402G, C23663T, G25562T, C25903T, C26455T, C28252T, A28253C, C28886T, G29556T).

We also performed the following in-silico inclusivity analysis. First, we downloaded more than 22,544 high-quality SARS-CoV-2 viral genome nucleotide sequences (16,503 unique sequences) from GISAID (2/12/2021) in FASTA format. To enable efficient computational evaluation of these data, we indexed the combined FASTAs and split them into subsets for parallelization. We performed BLASTn alignments using all 994 probes found in the SARS-CoV-2 Research Panel, with the PolyA tail removed, against the parallelized database. We similarly eliminated the PolyA tail from alignment in our pipeline. We chose to present descriptive statistics of the data due to the breadth of high identity matches to the SARS-CoV-2 viral genome.

To account for potential cross-reactivity of probe sequences found in the SARS-CoV-2 Research Panel, we aligned reads to 29 microbial genomes and the human genome along with the SARS-CoV-2 viral genome downloaded from NCBI (**Table S5**). A BLASTn (NCBI) analysis was then performed to quantify the number of primer pairs that had more than 80% homology with each of the genomes in the cohort. Then, we extended our in-silico exclusivity analysis using approximately 3.6 million viral nucleotide sequences from NCBI Virus (2/16/2021; FASTA format), representing the entire available database of viral nucleotide sequences.

### Library Preparation and Sequencing for Metagenomics

Metagenomic libraries were prepared with the Nextera Library Preparation Kit (Illumina) using 1-10 ng of input DNA. After library generation, 12-16 uniquely barcoded libraries were pooled and ran on an Illumina NextSeq 550 platform using a NextSeq500/550 Mid Output kit (Illumina) set to 150bp single-end reads, which yielded an average of 10M and 7.3M reads from positive and negative samples, respectively. After removal of reads mapping to human DNA, an average of 1.7M (positive) and 1.9 M (negative) microbial reads were used for further analysis.

### Quality Control

Quality control steps were performed after nucleic acid extraction, cDNA synthesis, cDNA library generation, target enrichment, metagenomic library, and final sequencing pools. These steps included analyses to determine nucleic acid concentration and fragment size using Qubit RNA High Sensitivity/ dsDNA High Sensitivity/ dsDNA Broad Range Quantitation Assay (Thermo Fisher Scientific, Waltham, MA) and TapeStation D5000/ D1000/ D1000 High Sensitivity platform (Agilent, Santa Clara, CA).

### Internal Controls

Viral transport media (VTM) was used as a negative/no template control (NTC) to eliminate the possibility of sample contamination on the assay run and was used with each extraction batch through sequencing. VERO E6 cells [ATCC CRL-1586] spiked into VTM were used as a negative extraction control (NEC) to monitor for any cross-contamination that occurs during the extraction process, as well as an extraction control to validate extraction reagents and successful RNA extraction. Positive Twist RNA template control - synthetic SARS-CoV-2 RNA Control 2 (MN908947.3) in Gene Expression Universal Reference RNA (Human, Agilent) was used as a positive control (PC) to verify that the assay run was performing as intended. The PC is made of six RNA fragments that are 5000bp in length spanning the entire SARS-CoV-2 viral genome (MN908947.3). All coding and non-coding regions of the viral genome are included in the PC, except for the polyadenylated region. Each RNA fragment is made by *in vitro* transcription using the DNA template as reference. Gene Expression Universal Reference RNA (Human, Agilent) was used as an internal control (IC) to validate the SARS-CoV-2 NGS Assay reagents and successful library generation. Additionally, the number of reads mapped to the human genome was determined and used as internal control in each specimen.

### SARS-CoV-2 NGS Assay Analysis

The data was analyzed using a cloud-based Biotia COVID-DX (v1.0) Software that has been optimized for the SARS-CoV-2 NGS Assay. The pipeline software is contained within Docker images orchestrated by Cromwell / WDL backed by Azure Batch (51). The analysis pipeline included a step to remove low-quality reads, alignment to a reference viral and host genomes, extraction of mapped reads, calculating coverage across the genome, and modeling of coverage to determine presence/absence of the SARS-CoV-2 virus. Furthermore, we detected genomic variants, analyzed phylogenetic and geographic origin, and quantified the SARS-CoV-2 virus.

#### Preprocessing of Sequencing Data

The program cutadapt trimmed adapter sequences using the standard Illumina adapter sequence “AGATCGGAAGAGC” and a minimum read length of 20 bp. Samples with higher than 50,000,000 reads were subsampled to 50,000,000 reads using seqtk before further analysis. FASTQs were split for parallelization and aligned against the genomes of SARS-CoV-2 virus (NC_045512.2), 26 other viral and bacterial respiratory pathogens, and the human genome using Bowtie2 run in local mode. The algorithm to detect SARS-CoV-2 used only reads that map unambiguously to the SARS-CoV-2 reference (NC_045512.2) in order to minimize false positives due to cross-hybridization of the probes or cross-contamination during the Twist NGS hybrid capture. Unambiguously mapped reads were extracted using samtools-view. Duplicates and read statistics were calculated with Picard (2.23.0) (52) and the CollectHSMetrics function. The depth of reads at each site across the SARS-CoV-2 genome is calculated using samtools-depth. Test samples and/or positive controls with fewer than 10,000 total reads are assigned as invalid.

#### Presence/absence calculation and coverage

Detection of the SARS-CoV-2 virus is based on the degree to which the genome was recovered in sequencing. The more genome that is recovered, the more likely it is that the virus is present. A sample-specific integral is computed by calculating the coverage at 1X depth using a sliding window scheme (with a window size of 1000 and step size of 100). This metric is then log transformed. A threshold of 8.6 for samples with less than or equal to 10,000 bases on target or 9.6 for samples with more than 10,000 bases on target determines if SARS-CoV-2 is present or absent. The integral is calculated using the R statistical software package (v4.0.1).

#### Genetic Variants Detection and Phylogenetic Analysis

The BAM file was subsampled to a maximum of 500X coverage at a given site within the SARS-CoV-2 viral genome because coverage can exceed 8,000X, which is computationally prohibitive for variant calling tools. Variants were called in the subsampled BAM file using GATK GenotypeVCF and HaplotypeCaller. The test sample HaplotypeCaller VCF file was merged with a NextStrain VCF file with 3365 samples using BCFtools. A phylogenetic tree was created using R-packages SNPRelate and gdsfmt. The tree was built using hierarchical cluster analysis on Identify by State (IBS) genetic pairwise distances matrix built on GISAID variants reported (version 23-April-2020). Clade geographic data were calculated using GISAID (version 24-June-2020) data to provide the percentage of samples sequenced in each country that are categorized into each clade among all samples of each clade.

The genetic variant annotation provides information on the mutation name, gene location, protein name, amino acid change (NCBI), and synonymy. Additional information included the frequency of genetic variants in our sample cohort and the associated read depth. The Grantham scoring system was used to designate conservative and radical mutations. In this system, the score of 100 and above calls mutations radical.

To validate the variant calling of our assay and software, we selected 3 genetic regions containing mutations identified by our protocol (**Table S7A**). The genetic regions were PCR amplified using specifically designed primers followed by Sanger sequencing. The list of validated genetic variants and the primer sets are described in the supplemental table (**Table S7A**). The primer design considered nearby mutations to the selected variants in order to maximize the validation efficiency of the sequencing reaction.

#### Analytical Sensitivity of SARS-CoV-2 NGS Assay

The analytical sensitivity (Limit of Detection [LoD]) study established the lowest concentration of SARS-CoV-2 viral genome (copies/ml) that can be detected by the SARS-CoV-2 NGS Assay at least 95% of the time. The preliminary LoD was established by testing 10-fold dilutions of SARS-CoV-2 synthetic RNA (MN908947.3, Twist Bioscience, #102024). The preliminary LoD was confirmed by testing triplicates of 2-fold dilutions (2560 copies/ml, 1280 copies/ml, 640 copies/ml, 320 copies/ml, 160 copies/ml, 80 copies/ml, and 40 copies/ml) by spiking the quantified heat-inactivated SARS-CoV-2 (ATCC, VR-1986HK) into negative respiratory clinical matrices (NP swabs -clinical samples previously tested negative for SARS-CoV-2 RNA). Then, the LoD was replicated 30 times.

### Clinical Metagenomics

Metagenomics has been processed through XSEDE and Bridges system at the Pittsburgh Supercomputing Center (PSC) (53, 54). Quality analysis was performed with FastQC (v0.11.3) (55) with default settings to validate the quality of the raw sequencing data. For quality control, adapter trimming and quality filtering were performed using the software fastp (v0.21.0) (56). Four functions of fastp were used: trimming of auto-detected adapter sequences, quality trimming at 5’, quality trimming at 3’, and a screening to detect 512 adapters, including Illumina’s Nextera transposase. In order to remove all human reads, Bowtie2 (v2.3.4.1) (57) and samtools (v1.9) (58) were used to align the quality filtered reads to *Homo sapiens* reference genome (GCF_000001405.39_GRCh38.p13). A quality control with FastQC was performed again to ensure the unmapped reads obtained were cleaned to run metagenomics tools. Processed reads were analyzed utilizing MetaPhlAn2 (59) to identify the relative abundance of microbial species. An additional k-mer based classification tool (BIOTIA-DX) was used as an orthogonal bioinformatic validation tool and to improve sensitivity (60). To explore functional genomic profiling, we used HUMAnN2 (61) to characterize microbial pathways linked to the presented microbial species. Antimicrobial resistance and virulence factors were determined using our k-mer based approach matched to the Comprehensive Antibiotic Resistance Database (CARD) (62) and Virulence Factor Database (VFDB) (62).

#### Metagenomic Validation

Since *Prevotella spp*. were found to be a biomarker for CVP status based on the data obtained by two independent bioinformatic pipelines but were found to conflict at the species level classification, we used 16s rDNA sequencing to aid in confirming the phylogeny of the correct *Prevotella* species. MetaPhlAn2 classified the biomarker as *Prevotella salivae* (LDA score: 3.77) while BIOTIA-DX called it as *Prevotella oral taxon 299* (LDA score: 3.81). For primer design*, Prevotella spp*. type strain 16s rDNA sequences from the ribosomal database project (RDP) (62), including *Prevotella sp. oral taxon* 299, were aligned. We modified Zozaya-Hinchliffe et al. (63) forward primer to add degeneracy and designed a reverse primer that would yield ∼1200bp of the 16s rDNA (**Table S8B**). The primer set was then tested for *in-silico* specificity against the curated 16s rDNA dataset from RDP and was found to be specific to 29/44 *Prevotella spp.,* out of the 13,324 16S rDNA type strain sequences present in the database, including those of interest to this study. The PCR amplification condition and primer set are described in **Table S8B**. PCR amplicons were excised from a modified-TAE agarose gel and purified with Millipore Ultrafree™-DA Centrifugal Filter Device, cloned, and sent for Sanger sequencing with T7/SP6 primers in a reference laboratory. Vector sequences were trimmed from 16s rDNA sequences, then sequences were aligned with all *Prevotella spp*. type strains. Kimura-2-parameter neighbor-joining phylogenetic tree with bootstrap of 500 was constructed with MegaX.

#### Statistical Analysis

Alpha and beta diversity metrics were calculated with the vegan R package using the relative abundance and count outputs obtained from MetaPhlAn2 and BIOTIA-DX. The variant heatmap and the microbiome heatmap visualizations with respective annotations were plotted using the Complex Heatmap R package. All Pearson correlations and correlation plots were calculated and visualized with the Corrplot R package. All other basic statistics and figures were generated with Prism V9.

## Supporting information

Supplemental Figures with Legends

## Data Availability

All data needed to evaluate conclusions are present in the manuscript. Supplemental data and sequencing data were submitted to GISAID.

https://www.gisaid.org/

## ACKNOWLEDGEMENTS

The COVID-DX software pipeline work used CromwellOnAzure, the Microsoft Genomics supported implementation of the Broad Institute’s Cromwell workflow engine on Azure. The metagenomic work used the Extreme Science and Engineering Discovery Environment (XSEDE), which is supported by National Science Foundation grant (ACI-1548562). Specifically, it used the Bridges system, which is supported by NSF award (ACI-1445606), at the Pittsburgh Supercomputing Center (PSC). This work used resources of the COVID-19 HPC Consortium. We thank Nicholas Nystrom, Paola Buitrago, Julian Uran, David O’Neal and collaborators for their assistance with PSC resources access, use, and software installation support. We also thank Zaineb Bello and Caitlin Otto for their laboratory operation support; the Abesse Team, Pierre Davidoff, Shay David and Cory Mason for IT support; Peter Eugster and Bryan Hoglund for logistical support; Shakil Ahmed for regulatory support; and Bradley Connor for his clinical advice and reporting support.

## Author contributions

DNS, MCR, CEM and NBO designed the study. KB, SC designed the SARS-CoV-2 NGS assay; DNS, JB, HLW, MD, CEM, NBO designed and validated the analysis software tool; DNS, MCR, JB, performed the clinical validation; RJB, MKT and CBJ provided the clinical samples and orthogonal analysis; AB performed variant curation; DNS, MCR and MD designed and performed the metagenomic analysis. DNS, MCR, CH and NBO wrote the manuscript with input from all authors.

## Data and materials availability

All data needed to evaulate conclusions are present in the manuscript, supplemental data and sequencing data were submitted to GISAID.

## REFERENCES

1. Centers for Disease Control and Prevention. CDC COVID Data Tracker. Centers Dis Control Prev. Published online 2020.

2. Johns Hopkins. Track Reported Cases of COVID-19 Coronavirus Resource Center. Published online 2020.

3. Kumar R et al. COVID-19 diagnostic approaches: different roads to the same destination. VirusDisease. 2020;31(2):97–105.

4. Artesi M et al. A recurrent mutation at position 26340 of SARS-CoV-2 is associated with failure of the E Gene quantitative reverse transcription-PCR utilized in a commercial dual-target diagnostic assay. J Clin Microbiol. Published online 2020. doi:10.1128/JCM.01598-20

5. Butler D et al. Shotgun Transcriptome and Isothermal Profiling of SARS-CoV-2 Infection Reveals Unique Host Responses, Viral Diversification, and Drug Interactions. bioRxiv Prepr Serv Biol. Published online 2020. doi:10.1101/2020.04.20.048066

6. Gaudin M, Desnues C. Hybrid capture-based next generation sequencing and its application to human infectious diseases. Front Microbiol. 2018;9:2924.

7. Tyson JR et al. Improvements to the ARTIC multiplex PCR method for SARS-CoV-2 genome sequencing using nanopore. bioRxiv. Published online September 4, 2020. doi:10.1101/2020.09.04.283077

8. Twist_Bioscience. NGS Effects of Mismatches on DNA (White Paper). Published 2019. https://www.twistbioscience.com/sites/default/files/resources/2019-05/WhitePaper_NGS_EffectsofMismatchesonDNA_7May19_Rev1_0.pdf

9. Geneva: World Health Organization. Genomic sequencing of SARS-CoV-2: a guide to implementation for maximum impact on public health. Licence CC BY-NC-SA 30 IGO. Published online 2021.

10. Klempt P et al. Performance of targeted library preparation solutions for SARS-CoV-2 whole genome analysis. Diagnostics. 2020;10(10).

11. GISAID. GISAID Initiative. Adv Virus Res. Published online 2020.

12. Centers for Disease Control and Prevention. SARS-CoV-2 Sequencing for Public Health Emergency Response, Epidemiology and Surveillance (SPHERES). Published online 2020. https://www.cdc.gov/coronavirus/2019-ncov/covid-data/spheres.html

13. Mostafa HH et al. Metagenomic Next-Generation Sequencing of Nasopharyngeal Specimens Collected from Confirmed and Suspect COVID-19 Patients. MBio. 2020;11(6):1–13.

14. Khan AA, Khan Z. COVID-2019-associated overexpressed Prevotella proteins mediated host-pathogen interactions and their role in coronavirus outbreak. Bioinformatics. 2020;36(13):4065-4069.

15. MacKay MJ et al. The COVID-19 XPRIZE and the need for scalable, fast, and widespread testing. Nat Biotechnol. 2020;38(9):1021–1024.

16. Duchene S et al. Temporal signal and the phylodynamic threshold of SARS-CoV-2. Virus Evol. 2020;6(2).

17. Korber B et al. Tracking Changes in SARS-CoV-2 Spike: Evidence that D614G Increases Infectivity of the COVID-19 Virus. Cell. 2020;182(4):812–827.e19.

18. Callaway E, Ledford H. How to redesign COVID vaccines so they protect against variants. Nature. 2021;590(7844):15-16.

19. Volz E, et al. Transmission of SARS-CoV-2 Lineage B.1.1.7 in England: Insights from linking epidemiological and genetic data. medRxiv. Published online 2021.

20. Plante JA et al. Spike mutation D614G alters SARS-CoV-2 fitness. Nature. Published online 2020. doi:10.1038/s41586-020-2895-3

21. McCarthy KR et al. Natural deletions in the SARS-CoV-2 spike glycoprotein drive antibody escape. Science (80-). 2021: Online ahead of print. doi:10.1101/2020.11.19.389916

22. Grubaugh ND et al. Making Sense of Mutation: What D614G Means for the COVID-19 Pandemic Remains Unclear. Cell. 2020;182(4):794–795.

23. Walls AC et al. Tectonic conformational changes of a coronavirus spike glycoprotein promote membrane fusion. Proc Natl Acad Sci U S A. 2017;114(42):11157–11162.

24. Davies NG, et al. Estimated transmissibility and severity of novel SARS-CoV-2 Variant of Concern 202012/01 in England. *medRxiv*. Published online 2020.

25. Tegally H et al. Emergence and rapid spread of a new severe acute respiratory syndrome-related coronavirus 2 (SARS-CoV-2) lineage with multiple spike mutations in South Africa. medRxiv. Published online 2020.

26. Rambaut A, et al. Preliminary genomic characterisation of an emergent SARS-CoV-2 lineage in the UK defined by a novel set of spike mutations - SARS-CoV-2 coronavirus / nCoV-2019 Genomic Epidemiology - Virological. virological.org.

27. Oberemok V V. et al. SARS-CoV-2 will continue to circulate in the human population: an opinion from the point of view of the virus-host relationship. Inflamm Res. 2020;69(7):635–640.

28. FDA. Genetic Variants of SARS-CoV-2 May Lead to False Negative Results with Molecular Tests for Detection of SARS-CoV-2 - Letter to Clinical Laboratory Staff and Health Care Providers. Published 2021. https://www.fda.gov/medical-devices/letters-health-care-providers/genetic-variants-sars-cov-2-may-lead-false-negative-results-molecular-tests-detection-sars-cov-2

29. Charre C et al. Evaluation of NGS-based approaches for SARS-CoV-2 whole genome characterisation. Virus Evol. 2020;6(2).

30. Xiao M et al. Multiple approaches for massively parallel sequencing of SARS-CoV-2 genomes directly from clinical samples. Genome Med. 2020;12(1).

31. Samorodnitsky E et al. Evaluation of Hybridization Capture Versus Amplicon-Based Methods for Whole-Exome Sequencing. Hum Mutat. 2015;36(9):903–914.

32. Kumpitsch C et al. The microbiome of the upper respiratory tract in health and disease. BMC Biol. 2019;17(1).

33. Bomar L et al. Bacterial microbiota of the nasal passages across the span of human life. Curr Opin Microbiol. 2018;41:8–14.

34. Edouard S et al. The nasopharyngeal microbiota in patients with viral respiratory tract infections is enriched in bacterial pathogens. Eur J Clin Microbiol Infect Dis. 2018;37(9):1725–1733.

35. Hanada S et al. Respiratory viral infection-induced microbiome alterations and secondary bacterial pneumonia. Front Immunol. 2018;9(NOV).

36. Tsang TK et al. Association between the respiratory microbiome and susceptibility to influenza virus infection. Clin Infect Dis. 2020;71(5):1195–1203.

37. Lee KH et al. The respiratory microbiome and susceptibility to influenza virus infection. PLoS One. 2019;14(1):e0207898.

38. Fazlollahi M et al. The nasal microbiome in asthma. J Allergy Clin Immunol. 2018;142(1):834–843.e2.

39. Demuri GP et al. Dynamics of Bacterial Colonization with Streptococcus pneumoniae, Haemophilus influenzae, and Moraxella catarrhalis during Symptomatic and Asymptomatic Viral Upper Respiratory Tract Infection. Clin Infect Dis. 2018;66(7):1045–1053.

40. Marotz C et al. Microbial context predicts SARS-CoV-2 prevalence in patients and the hospital built environment. medRxiv. Published online 2020. doi:10.1101/2020.11.19.20234229

41. Rawson TM et al. Bacterial and Fungal Coinfection in Individuals With Coronavirus: A Rapid Review To Support COVID-19 Antimicrobial Prescribing. Clin Infect Dis. 2020;71(9):2459–2468.

42. Saleem Z et al. Point prevalence surveys of health-care-associated infections: a systematic review. Pathog Glob Health. 2019;113(4):191–205.

43. Mirzaei R et al. Bacterial co-infections with SARS-CoV-2. IUBMB Life. 2020;72(10):2097–2111.

44. Lijek RS, Weiser JN. Co-infection subverts mucosal immunity in the upper respiratory tract. Curr Opin Immunol. 2012;24(4):417–423.

45. Hart OE, Halden RU. Computational analysis of SARS-CoV-2/COVID-19 surveillance by wastewater-based epidemiology locally and globally: Feasibility, economy, opportunities and challenges. Sci Total Environ. 2020;730.

46. Goldstein T et al. The discovery of Bombali virus adds further support for bats as hosts of ebolaviruses. Nat Microbiol. 2018;3(10):1084–1089.

47. Marsh GA et al. Cedar Virus: A Novel Henipavirus Isolated from Australian Bats. PLoS Pathog. 2012;8(8).

48. Ren W et al. Difference in Receptor Usage between Severe Acute Respiratory Syndrome (SARS) Coronavirus and SARS-Like Coronavirus of Bat Origin. J Virol. 2008;82(4):1899–1907.

49. Wells HL et al. The evolutionary history of ACE2 usage within the coronavirus subgenus Sarbecovirus. bioRxiv. Published online 2020. doi:10.1101/2020.07.07.190546

50. Centers for Disease Control and Prevention (CDC). Interim Guidelines for Clinical Specimens for COVID-19. Centers Dis Control Prev. Published online 2020.

51. Cromwell on Azure (Online GitHub). https://github.com/microsoft/CromwellOnAzure

52. Broad Institute. Picard Tools - By Broad Institute. Github.

53. Nystrom NA et al. Bridges: A uniquely flexible HPC resource for new communities and data analytics. In: ACM International Conference Proceeding Series.; 2015:1-8.

54. Towns J et al. XSEDE: Accelerating scientific discovery. Comput Sci Eng. 2014;16(5):62–74.

55. Andrews S. FASTQC A Quality Control tool for High Throughput Sequence Data. *Babraham Inst*. Published online 2015.

56. Chen S et al. Fastp: An ultra-fast all-in-one FASTQ preprocessor. In: Bioinformatics.; 2018. doi:10.1093/bioinformatics/bty560

57. Langmead B, Salzberg SL. Fast gapped-read alignment with Bowtie 2. Nat Methods. 2012;9(4):357–359.

58. Li H et al. The Sequence Alignment/Map format and SAMtools. Bioinformatics. 2009;25(16):2078–2079.

59. Truong DT et al. MetaPhlAn2 for enhanced metagenomic taxonomic profiling. Nat Methods. 2015;12(10):902–903.

60. O’Hara NB et al. Metagenomic characterization of ambulances across the USA. Microbiome. 2017;5(1):125.

61. Franzosa EA et al. Species-level functional profiling of metagenomes and metatranscriptomes. Nat Methods. 2018;15(11):962–968.

62. McArthur AG et al. The comprehensive antibiotic resistance database. Antimicrob Agents Chemother. 2013;57(7):3348–3357.

63. Zozaya-Hinchliffe M et al. Quantitative PCR assessments of bacterial species in women with and without bacterial vaginosis. J Clin Microbiol. 2010;48(5):1812–1819.

