## Supplemental Figures with Legends for "Targeted Hybridization Capture of SARS-CoV-2 and Metagenomics Enables Genetic Variant Discovery and Nasal Microbiome Insights"

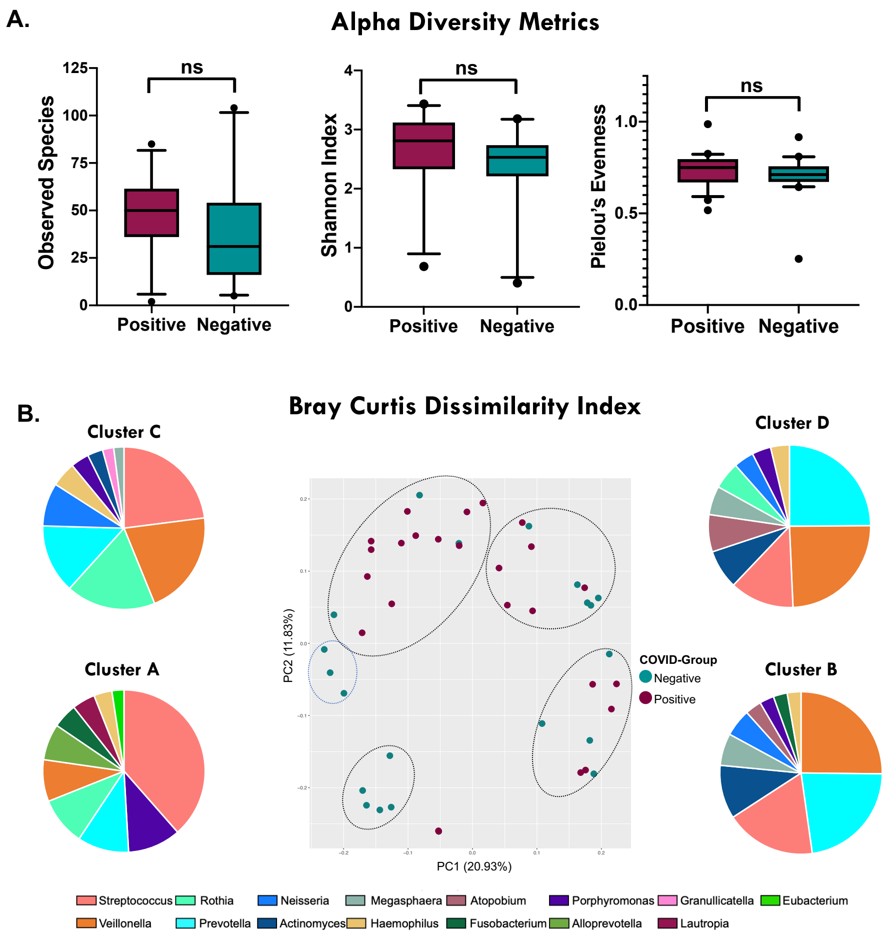


**Figure S1. Nasopharyngeal microbiome profiles are not driven by COVID-status.** (A) Alpha diversity metrics show no difference in microbiome richness or evenness based on COVID status. Observed Species, Shannon Index and Pielou's Evenness were not significant based on a Welch's t-test comparing CVP (n= 26) and CVN (n = 22). (B) Bray Curtis Dissimilarity Index exhibited four clusters (A, B, C, and D) with most of the separation driven by *Rothia mucilaginosa* along PC1 (20.9%) and clustering along. PC2 (11.83%) is explained by the presence/absence of *Streptococcus parasanguinis* and unclassified *Neisseria.* Pie charts represent the top 10 most abundant bacterial genera among samples within each cluster.

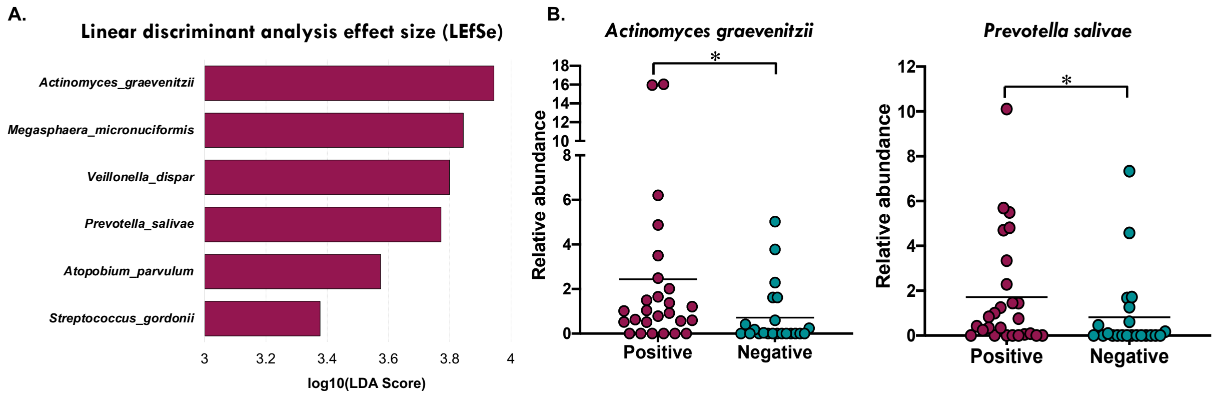


**Figure S2. Nasopharyngeal microbiome revealed the presence of biomarkers associated to CVP samples.** (A) LEfSe analysis showed six bacterial species to be increased in CVP samples. (B) Mann-Whitney-U-test confirmed significant differences in the relative abundance of *Actinomyces graevenitzii* (p = 0.0130) and *Prevotella salivae* (p=0.0394) of CVP samples (n=26) versus CVN samples (n=22).

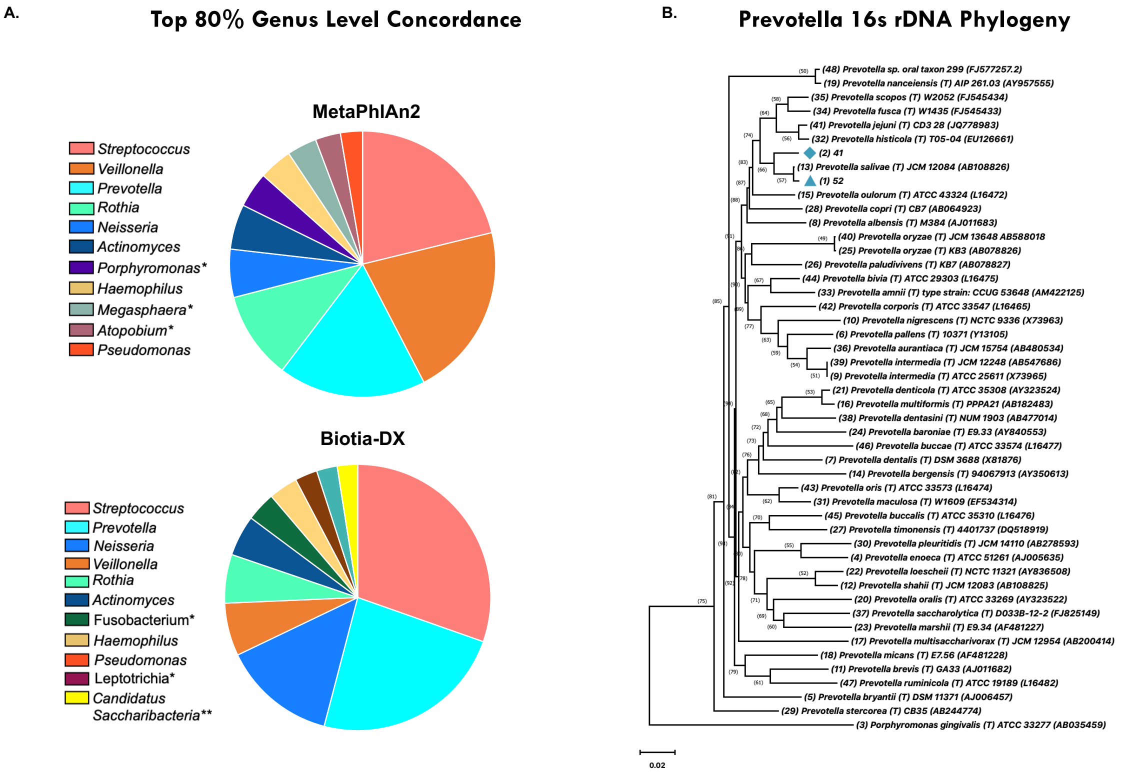


**Figure S3. Comparison bioinformatic pipelines exhibit high concordance in the classification of the nasopharyngeal microbiome.** (A) Pie charts depicting the top 80% relative abundance of bacteria at the genus level for MetaPhlAn2 and BIOTIA-DX classification. (B) Neighbor joining phylogenetic tree of partial *Prevotella spp*.16s rDNA confirms the *Prevotella* species increased in CVP samples is closely related to *Prevotella salivae*. The optimal tree with the sum of branch length (=1.65) is shown. The percentage of replicate trees in which the associated taxa clustered together in the bootstrap test (500 replicates) is shown next to the branches. The tree is drawn to scale, with branch lengths in the same units as those of the evolutionary distances used to infer the phylogenetic tree. The evolutionary distances were computed using the Kimura-2-parameter method and are in the units of the number of base substitutions per site. This analysis involved 48 nucleotide sequences. All ambiguous positions were removed for each sequence pair (pairwise deletion option). There were a total of 997 positions in the final dataset. Evolutionary analyses were conducted in MEGA X.

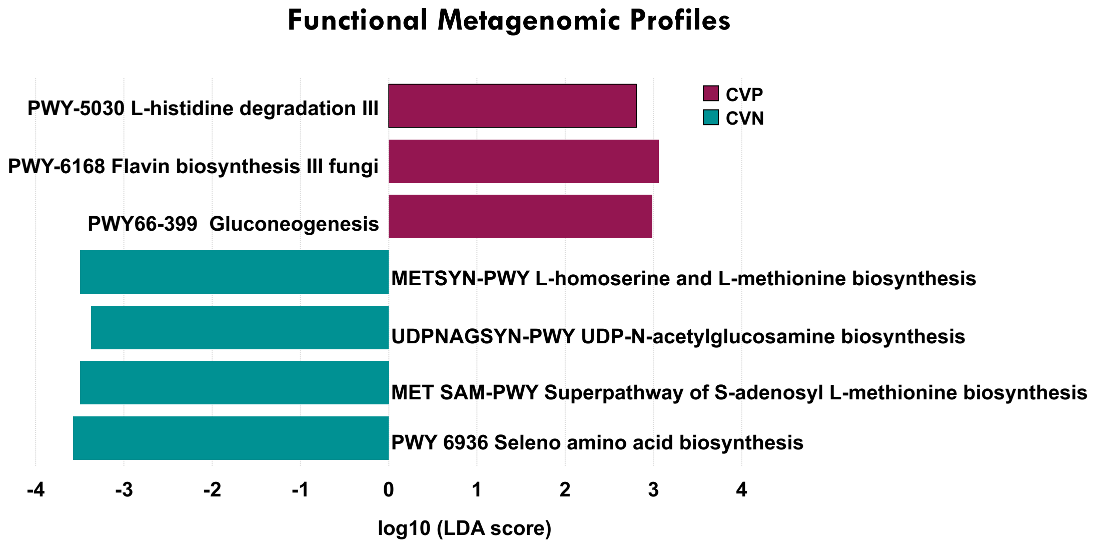


**Figure S4. Functional metagenomic profile in association with the COVID status.** We detected 3 pathways [gluconeogenesis (PWY-66-399), flavin biosynthesis (PWY-6168) and L-histidine degradation (PWY-5030)] that were increased in the CVP samples. 4 metabolic pathways were found to be overrepresented in CVN samples [*L*-homoserine and *L*-methionine biosynthesis, UDP-N-acetylglucosamine biosynthesis, seleno amino acid biosynthesis (PWY-6936) and S-adenosyl L-methionine biosynthesis superpathway].
