## Supplemental Tables for "Targeted Hybridization Capture of SARS-CoV-2 and Metagenomics Enables Genetic Variant Discovery and Nasal Microbiome Insights"

**Supplemental Table 1: Performance of Clinical Evaluation**

**A)** Clinical performance of the SARS-CoV-2 NGS Assay was evaluated by comparing results to an RT-PCR assay authorized by the FDA for use under Emergency User Authorization (EUA RT-PCR). 60 clinical nasopharyngeal swab (NP) specimens were tested, including 30 SARS-CoV-2-positive and 30 SARS-CoV-2-negative specimens. The positive and negative percent agreement was calculated in relation to the EUA RT-PCR comparator method and indicated in the following table.

| | | EUA RT-PCR Comparator Assay | | |
|---|---|---|---|---|
| | | Positive | Negative | % agreement (95% CI) |
| SARS-CoV-2 NGS Assay | Positive | 29 | 0 | PPA: 96.7% (90.6%-100%) |
| | Negative | 1 | 29 | NPA: 96.7% (90.6%-100%) |
| | Invalid | 0 | 1 | |

**One PCR negative sample did not yield sufficient reads using the SARS-CoV-2 NGS Assay to be called negative, and labelled as INVALID.*

**B)** Additional independent validation set combined with previous validation data yielded the following performance characteristics:

| | | EUA RT-PCR Comparator Assay | | |
|---|---|---|---|---|
| | | Positive | Negative | % agreement (95% CI) |
| SARS-CoV-2 NGS Assay | Positive | 57 | 0 | PPA:95.2% (90%-100%) |
| | Negative | 3 | 59 | NPA: 98.3% (95.2%-100%) |
| | Invalid | 0 | 1 | |

**One PCR negative sample did not yield sufficient reads using the SARS-CoV-2 NGS Assay to be called negative, and labelled as INVALID.*

**Supplemental Table 2. The calculation of presence and absence of SARS-CoV-2 viral genome in clinical validation and an independent clinical validation.** We devised a metric to compute the integral under a curve created by calculating the coverage at 1X depth using a sliding window scheme (with a window size of 100 and step size of 10). This metric was then log transformed. We have set a threshold of 8.6 if samples had less than 10,000 bases on target or 9.5 if samples had more than 10,000 bases on target. Metrics were calculated using the R statistical software package (v4.0.1).

Presence: if SARS-CoV-2 was detected. We devised a metric [I1] to compute the integral under a curve created by calculating the coverage at 1X depth using a sliding window scheme (with a window size of 100 and step size of 10). This metric was then log transformed. We have set a threshold of 8.6 if samples had less than 10,000 bases on target or 9.5 if samples had more than 10,000 bases on target. Metrics were calculated using the R statistical software package (v4.0.1). X1-X500: percent coverage at 1X-500X depths, used internally to inform the analysis. E1: evenness, used internally to inform the analysis. I1: integral. The following HS metrics are reported: bait territory, percent of selected bases, target territory, genome size, total reads and on target bases. Red indicates deviant results. Additionally, the number of reads mapped to the human genome will provide an internal control in each individual sample to be called valid.

| Specimen_ID | Specimen_ID | TYPE_PCR | presence | clade | reads | X1 | X5 | X10 | X50 | X100 | X500 | E1 | I1 | validity | BAIT_TERRITO | PCT_SELECTED_B | TARGET_TERR | GENOME_SIZE | TOTAL_READS | ON_TARGET_E | HUMAN_ ON_TARGET_BASES |
|---|---|---|---|---|---|---|---|---|---|---|---|---|---|---|---|---|---|---|---|---|---|
| COVID-NEG-002_S6 | CVN-1 | NEGATIVE | FALSE | NOT_IDENTIFII | 2473492 | 0 | 0 | 0 | 0 | 0 | 0 | 0 | 6 | VALID | 29869 | 0 | 29870 | 3259356417 | 2467880 | 2995 | 226429263 |
| COVID-NEG-003_S7 | CVN-2 | NEGATIVE | FALSE | NOT_IDENTIFII | 3252564 | 0 | 0 | 0 | 0 | 0 | 0 | 0 | 4 | VALID | 29869 | 0 | 29870 | 3259356417 | 3247611 | 11 | 298210227 |
| COVID-Neg-004_S4 | CVN-3 | NEGATIVE | FALSE | NOT_IDENTIFII | 2288625 | 0 | 0 | 0 | 0 | 0 | 0 | 0 | 4 | VALID | 29869 | 0 | 29870 | 3259356417 | 2248079 | 1215 | 121345936 |
| COVID-NEG-005_S9 | CVN-4 | NEGATIVE | FALSE | NOT_IDENTIFII | 10801026 | 0 | 0 | 0 | 0 | 0 | 0 | 0 | 9 | VALID | 29869 | 0 | 29870 | 3259356417 | 10775824 | 11041 | 371203778 |
| COVID-NEG-006_S1 | CVN-5 | NEGATIVE | FALSE | NOT_IDENTIFII | 2061968 | 0 | 0 | 0 | 0 | 0 | 0 | 0 | 7 | VALID | 29869 | 0 | 29870 | 3259356417 | 2053094 | 1761 | 129756751 |
| COVID-NEG-007_S3 | CVN-6 | NEGATIVE | FALSE | NOT_IDENTIFII | 1064413 | 0 | 0 | 0 | 0 | 0 | 0 | 0 | 9 | VALID | 29869 | 0 | 29870 | 3259356417 | 1057055 | 6583 | 70875601 |
| COVID-NEG-008_S3 | CVN-7 | NEGATIVE | FALSE | NOT_IDENTIFII | 2357502 | 0 | 0 | 0 | 0 | 0 | 0 | 0 | 7 | VALID | 29869 | 0 | 29870 | 3259356417 | 2317506 | 1307 | 236724 |
| COVID-NEG-009_S3 | CVN-8 | NEGATIVE | FALSE | NOT_IDENTIFII | 2896238 | 0 | 0 | 0 | 0 | 0 | 0 | 0 | 7 | VALID | 29869 | 0 | 29870 | 3259356417 | 2893354 | 1215 | 121955798 |
| COVID-NEG-010_S3 | CVN-9 | NEGATIVE | FALSE | NOT_IDENTIFII | 4037293 | 0 | 0 | 0 | 0 | 0 | 0 | 0 | 6 | VALID | 29869 | 0 | 29870 | 3259356417 | 4033669 | 2806 | 156046426 |
| COVID-Neg-011_S4 | CVN-10 | NEGATIVE | FALSE | NOT_IDENTIFII | 1730315 | 0 | 0 | 0 | 0 | 0 | 0 | 0 | #NAME? | VALID | 29869 | 0 | 29870 | 3259356417 | 1727355 | 0 | 143663646 |
| COVID-NEG-012_S4 | CVN-11 | NEGATIVE | FALSE | NOT_IDENTIFII | 1452222 | 0 | 0 | 0 | 0 | 0 | 0 | 0 | 6 | VALID | 29869 | 0 | 29870 | 3259356417 | 1449645 | 3491 | 106590485 |
| COVID-NEG-013_S4 | CVN-12 | NEGATIVE | FALSE | NOT_IDENTIFII | 1814277 | 0 | 0 | 0 | 0 | 0 | 0 | 0 | 9 | VALID | 29869 | 0 | 29870 | 3259356417 | 1811892 | 17435 | 142326243 |
| COVID-NEG-014_S4 | CVN-13 | NEGATIVE | FALSE | NOT_IDENTIFII | 49438 | 0 | 0 | 0 | 0 | 0 | 0 | 0 | 7 | VALID | 29869 | 0 | 29870 | 3259356417 | 49344 | 1567 | 3017714 |
| COVID-NEG-015_S4 | CVN-14 | NEGATIVE | FALSE | NOT_IDENTIFII | 75993 | 0 | 0 | 0 | 0 | 0 | 0 | 0 | 8 | VALID | 29869 | 0 | 29870 | 3259356417 | 75797 | 2281 | 4176197 |
| COVID-NEG-016_S4 | CVN-15 | NEGATIVE | FALSE | NOT_IDENTIFII | 62897 | 0 | 0 | 0 | 0 | 0 | 0 | 0 | 7 | VALID | 29869 | 0 | 29870 | 3259356417 | 62444 | 2439 | 4525370 |
| COVID-NEG-017_S4 | CVN-16 | NEGATIVE | FALSE | NOT_IDENTIFII | 68069 | 0 | 0 | 0 | 0 | 0 | 0 | 0 | 7 | VALID | 29869 | 0 | 29870 | 3259356417 | 67913 | 1119 | 5251566 |
| COVID-NEG-018_S4 | CVN-17 | NEGATIVE | FALSE | NOT_IDENTIFII | 1580183 | 0 | 0 | 0 | 0 | 0 | 0 | 0 | 6 | VALID | 29869 | 0 | 29870 | 3259356417 | 1577096 | 4326 | 70789243 |
| COVID-NEG-019_S4 | CVN-18 | NEGATIVE | FALSE | NOT_IDENTIFII | 1110690 | 0 | 0 | 0 | 0 | 0 | 0 | 0 | #NAME? | VALID | 29869 | 0 | 29870 | 3259356417 | 1107581 | 0 | 96166744 |
| COVID-Neg-020_S4 | CVN-19 | NEGATIVE | FALSE | NOT_IDENTIFII | 2915736 | 0 | 0 | 0 | 0 | 0 | 0 | 0 | 6 | VALID | 29869 | 0 | 29870 | 3259356417 | 2913475 | 1183 | 203399293 |
| COVID-Neg-021_S4 | CVN-20 | NEGATIVE | FALSE | NOT_IDENTIFII | 2887705 | 0 | 0 | 0 | 0 | 0 | 0 | 0 | #NAME? | VALID | 29869 | 0 | 29870 | 3259356417 | 2855998 | 0 | 208946591 |
| COVID-NEG-022_S5 | CVN-21 | NEGATIVE | FALSE | NOT_IDENTIFII | 0 | 0 | 0 | 0 | 0 | 0 | 0 | 0 | #NAME? | INVALID | 29869 ? | | 29870 | 3259356417 | 0 | 0 | 0 |
| COVID-NEG-023_S5 | CVN-22 | NEGATIVE | FALSE | NOT_IDENTIFII | 210499 | 0 | 0 | 0 | 0 | 0 | 0 | 0 | 7 | VALID | 29869 | 0 | 29870 | 3259356417 | 209358 | 3504 | 13428920 |
| COVID-NEG-024_S5 | CVN-23 | NEGATIVE | FALSE | NOT_IDENTIFII | 235757 | 0 | 0 | 0 | 0 | 0 | 0 | 0 | #NAME? | VALID | 29869 | 0 | 29870 | 3259356417 | 235196 | 0 | 17213687 |
| COVID-Neg-025_S4 | CVN-24 | NEGATIVE | FALSE | NOT_IDENTIFII | 1098581 | 0 | 0 | 0 | 0 | 0 | 0 | 0 | 8 | VALID | 29869 | 0 | 29870 | 3259356417 | 1090704 | 116783 | 92272166 |
| COVID-NEG-026_S5 | CVN-25 | NEGATIVE | FALSE | NOT_IDENTIFII | 53559 | 0 | 0 | 0 | 0 | 0 | 0 | 0 | 7 | VALID | 29869 | 0 | 29870 | 3259356417 | 52490 | 5627 | 1753667 |
| COVID-NEG-027_S5 | CVN-26 | NEGATIVE | FALSE | NOT_IDENTIFII | 62180 | 0 | 0 | 0 | 0 | 0 | 0 | 0 | #NAME? | VALID | 29869 | 0 | 29870 | 3259356417 | 62023 | 0 | 3964318 |
| COVID-NEG-028_S5 | CVN-27 | NEGATIVE | FALSE | NOT_IDENTIFII | 67953 | 0 | 0 | 0 | 0 | 0 | 0 | 0 | 6 | VALID | 29869 | 0 | 29870 | 3259356417 | 67814 | 289 | 3865381 |
| COVID-NEG-029_S5 | CVN-28 | NEGATIVE | FALSE | NOT_IDENTIFII | 64494 | 0 | 0 | 0 | 0 | 0 | 0 | 0 | 4 | VALID | 29869 | 0 | 29870 | 3259356417 | 64331 | 114 | 3402919 |
| COVID-NEG-030_S5 | CVN-29 | NEGATIVE | FALSE | NOT_IDENTIFII | 131360 | 0 | 0 | 0 | 0 | 0 | 0 | 0 | #NAME? | VALID | 29869 | 0 | 29870 | 3259356417 | 131215 | 0 | 9137168 |
| COVID-NEG-031_S5 | CVN-30 | NEGATIVE | FALSE | NOT_IDENTIFII | 90980 | 0 | 0 | 0 | 0 | 0 | 0 | 0 | 5 | VALID | 29869 | 0 | 29870 | 3259356417 | 86763 | 101 | 4071720 |
| COVID-POS-006_S1 | CVP-1 | POSITIVE | TRUE | 20A | 3716751 | 1 | 1 | 1 | 1 | 1 | 0 | 1 | 10 | VALID | 29869 | 0 | 29870 | 3259356417 | 3713260 | 96984863 | 230534497 |
| COVID-POS-007_S2 | CVP-2 | POSITIVE | TRUE | 20C | 5376358 | 1 | 1 | 1 | 1 | 1 | 0 | 1 | 10 | VALID | 29869 | 0 | 29870 | 3259356417 | 5365099 | 110479051 | 348459749 |
| COVID-POS-008_S3 | CVP-3 | POSITIVE | TRUE | 20A | 5422292 | 1 | 1 | 1 | 1 | 1 | 0 | 1 | 10 | VALID | 29869 | 0 | 29870 | 3259356417 | 5417696 | 193149870 | 251751939 |
| COVID-POS-009_S4 | CVP-4 | POSITIVE | TRUE | 19A | 3465259 | 1 | 1 | 1 | 0 | 0 | 0 | 1 | 10 | VALID | 29869 | 0 | 29870 | 3259356417 | 3459617 | 908084 | 201234095 |
| COVID-POS-010_S5 | CVP-5 | POSITIVE | TRUE | 20A | 1256389 | 1 | 1 | 1 | 0 | 0 | 0 | 1 | 10 | VALID | 29869 | 0 | 29870 | 3259356417 | 1254963 | 919481 | 75421830 |
| COVID-POS-011_S1 | CVP-6 | POSITIVE | TRUE | 20C | 80637571 | 1 | 1 | 1 | 1 | 1 | 1 | 1 | 10 | VALID | 29869 | 1 | 29870 | 3259356417 | 49952761 | 5922268746 | 180963313 |
| COVID-POS-012_S1 | CVP-7 | POSITIVE | FALSE | NOT_IDENTIFII | 730999 | 0 | 0 | 0 | 0 | 0 | 0 | 0 | 7 | VALID | 29869 | 0 | 29870 | 3259356417 | 728300 | 1499 | 62478439 |
| COVID-POS-013_S1 | CVP-8 | POSITIVE | TRUE | 20C | 778532 | 1 | 1 | 1 | 1 | 1 | 0 | 1 | 10 | VALID | 29869 | 0 | 29870 | 3259356417 | 774436 | 15913656 | 48002284 |
| COVID-POS-014_S1 | CVP-9 | POSITIVE | TRUE | 20C | 6660390 | 1 | 1 | 1 | 1 | 1 | 0 | 1 | 10 | VALID | 29869 | 0 | 29870 | 3259356417 | 6655154 | 353142118 | 188374032 |
| COVID-POS-015_S1 | CVP-10 | POSITIVE | TRUE | 20C | 5737861 | 1 | 1 | 1 | 1 | 1 | 0 | 1 | 10 | VALID | 29869 | 0 | 29870 | 3259356417 | 5721633 | 24124077 | 368584192 |
| COVID-POS-016_S1 | CVP-11 | POSITIVE | TRUE | 20A | 6243585 | 1 | 1 | 1 | 1 | 1 | 0 | 1 | 10 | VALID | 29869 | 1 | 29870 | 3259356417 | 6236791 | 369722069 | 94636025 |
| COVID-POS-017_S1 | CVP-12 | POSITIVE | TRUE | 20C | 14308108 | 1 | 1 | 1 | 1 | 1 | 0 | 1 | 10 | VALID | 29869 | 1 | 29870 | 3259356417 | 14298158 | 1503464302 | 81052465 |
| COVID-POS-018_S1 | CVP-13 | POSITIVE | TRUE | 20C | 158979 | 1 | 1 | 1 | 1 | 1 | 0 | 1 | 10 | VALID | 29869 | 1 | 29870 | 3259356417 | 158739 | 11316086 | 4784065 |
| COVID-POS-019_S1 | CVP-14 | POSITIVE | TRUE | NOT_IDENTIFII | 68212 | 0 | 0 | 0 | 0 | 0 | 0 | 0 | 10 | VALID | 29869 | 0 | 29870 | 3259356417 | 68032 | 39061 | 4206422 |
| COVID-POS-020_S2 | CVP-15 | POSITIVE | TRUE | 20A | 60695705 | 1 | 1 | 1 | 1 | 1 | 1 | 1 | 10 | VALID | 29869 | 1 | 29870 | 3259356417 | 49950996 | 6339499218 | 14975922 |
| COVID-POS-021_S2 | CVP-16 | POSITIVE | TRUE | 19A | 244330 | 1 | 1 | 1 | 1 | 1 | 0 | 1 | 10 | VALID | 29869 | 0 | 29870 | 3259356417 | 244087 | 13267047 | 4964282 |
| COVID-POS-022_S2 | CVP-17 | POSITIVE | TRUE | 20C | 1178790 | 1 | 1 | 1 | 1 | 1 | 0 | 1 | 10 | VALID | 29869 | 0 | 29870 | 3259356417 | 1176340 | 3621916 | 98732729 |
| COVID-POS-023_S2 | CVP-18 | POSITIVE | TRUE | 20A | 1239946 | 1 | 1 | 1 | 1 | 1 | 0 | 1 | 10 | VALID | 29869 | 0 | 29870 | 3259356417 | 1219896 | 26468529 | 81436812 |
| COVID-POS-024_S2 | CVP-19 | POSITIVE | TRUE | 20A | 24316832 | 1 | 1 | 1 | 1 | 1 | 0 | 1 | 10 | VALID | 29869 | 1 | 29870 | 3259356417 | 24299551 | 2788286419 | 134469844 |
| COVID-POS-025_S2 | CVP-20 | POSITIVE | TRUE | 20A | 1255877 | 1 | 1 | 1 | 0 | 0 | 0 | 1 | 10 | VALID | 29869 | 0 | 29870 | 3259356417 | 1251199 | 1269979 | 112419865 |
| COVID-POS-026_S2 | CVP-21 | POSITIVE | TRUE | 20A | 239196 | 1 | 1 | 1 | 1 | 0 | 0 | 1 | 10 | VALID | 29869 | 0 | 29870 | 3259356417 | 238715 | 2192286 | 16427173 |
| COVID-POS-027_S2 | CVP-22 | POSITIVE | TRUE | 20A | 33357673 | 1 | 1 | 1 | 1 | 1 | 1 | 1 | 10 | VALID | 29869 | 1 | 29870 | 3259356417 | 33333057 | 4206709822 | 24886026 |
| COVID-POS-028_S6 | CVP-23 | POSITIVE | TRUE | 20C | 63104 | 1 | 1 | 1 | 1 | 1 | 0 | 1 | 10 | VALID | 29869 | 1 | 29870 | 3259356417 | 62936 | 6011953 | 885938 |
| COVID-POS-029_S2 | CVP-24 | POSITIVE | TRUE | 20C | 13271148 | 1 | 1 | 1 | 1 | 1 | 0 | 1 | 10 | VALID | 29869 | 1 | 29870 | 3259356417 | 13200740 | 1638993394 | 22155495 |
| COVID-POS-030_S2 | CVP-25 | POSITIVE | TRUE | 20C | 1738765 | 1 | 1 | 1 | 1 | 1 | 0 | 1 | 10 | VALID | 29869 | 1 | 29870 | 3259356417 | 1737643 | 213201349 | 5561486 |
| COVID-POS-031_S3 | CVP-26 | POSITIVE | TRUE | 19A | 77888 | 1 | 1 | 1 | 0 | 0 | 0 | 1 | 10 | VALID | 29869 | 0 | 29870 | 3259356417 | 77780 | 386475 | 5159470 |
| COVID-POS-032_S3 | CVP-27 | POSITIVE | TRUE | 19A | 117370 | 1 | 1 | 1 | 0 | 0 | 0 | 1 | 10 | VALID | 29869 | 0 | 29870 | 3259356417 | 117124 | 316600 | 6341409 |
| COVID-POS-033_S3 | CVP-28 | POSITIVE | TRUE | 20B | 53540594 | 1 | 1 | 1 | 1 | 1 | 1 | 1 | 10 | VALID | 29869 | 1 | 29870 | 3259356417 | 49959831 | 6206809044 | 29989106 |
| COVID-POS-034_S3 | CVP-29 | POSITIVE | TRUE | 19A | 102921 | 1 | 0 | 0 | 0 | 0 | 0 | 0 | 10 | VALID | 29869 | 0 | 29870 | 3259356417 | 102661 | 71097 | 6979518 |
| COVID-POS-035_S3 | CVP-30 | POSITIVE | TRUE | 19A | 113558 | 1 | 1 | 1 | 0 | 0 | 0 | 1 | 10 | VALID | 29869 | 0 | 29870 | 3259356417 | 113339 | 425426 | 7062458 |
| Ctrl1-1e4_S63_L001 | PC-1: Positive control (10,000 | NA | TRUE | 19A | 503319 | 1 | 1 | 1 | 1 | 1 | 0 | 1 | 10 | VALID | 29869 | 0 | 29870 | 3259356417 | 502730 | 15570678 | 3979669 |
| Ctrl2-1e6-1_S61_L0 | PC-2: Positive control (1,000,0 | NA | TRUE | 19A | 47012721 | 1 | 1 | 1 | 1 | 1 | 1 | 1 | 10 | VALID | 29869 | 1 | 29870 | 3259356417 | 46976306 | 5807449979 | 3285532 |
| Ctrl2-1e6-2_S62_L0 | PC-3: Positive control (1,000,0 | NA | TRUE | 19A | 42266116 | 1 | 1 | 1 | 1 | 1 | 0 | 1 | 10 | VALID | 29869 | 1 | 29870 | 3259356417 | 42210524 | 3640762499 | 191152657 |
| HuRef-1_S64_L001_ | IC-1: Negative internal control | NA | FALSE | NOT_IDENTIFII | 352565 | 0 | 0 | 0 | 0 | 0 | 0 | 0 | 5 | VALID | 29869 | 0 | 29870 | 3259356417 | 351931 | 154 | 2140243 |
| HuRef-2_S65_L001_ | IC-2: Negative internal control | NA | FALSE | NOT_IDENTIFII | 10516002 | 0 | 0 | 0 | 0 | 0 | 0 | 0 | 6 | VALID | 29869 | 0 | 29870 | 3259356417 | 10501466 | 113 | 82856654 |
| NC-1_S66_L001_R1 | NTC-1: Negative template cont | NA | FALSE | NOT_IDENTIFII | 61 | 0 | 0 | 0 | 0 | 0 | 0 | 0 | #NAME? | VALID | 29869 | 0 | 29870 | 3259356417 | 20 | 0 | 544 |
| NC-2_S67_L001_R1 | NTC-2: Negative template cont | NA | FALSE | NOT_IDENTIFII | 1552 | 0 | 0 | 0 | 0 | 0 | 0 | 0 | 5 | VALID | 29869 | 0 | 29870 | 3259356417 | 833 | 3404 | 20909 |

| Specimen_ID | Specimen_ID | TYPE_PCR | presence | clade | reads | X1 | X5 | X10 | X50 | X100 | X500 | E1 | I1 | validity | BAIT_TERRITO | PCT_SELECTED_B | TARGET_TERR | GENOME_SIZE | TOTAL_READS | ON_TARGET_E | HUMAN_ ON_TARGET_BASES |
|---|---|---|---|---|---|---|---|---|---|---|---|---|---|---|---|---|---|---|---|---|---|
| CVN-032_S1_L001_ | CVN-31 | NEGATIVE | FALSE | NOT_IDENTIFII | 1875341 | 0 | 0 | 0 | 0 | 0 | 0 | 0 | 8 | VALID | 29869 | 0 | 29870 | 3259356417 | 1830300 | 5 | 11623695 |
| CVN-033_S2_L001_ | CVN-32 | NEGATIVE | FALSE | NOT_IDENTIFII | 1296720 | 0 | 0 | 0 | 0 | 0 | 0 | 0 | 8 | VALID | 29869 | 0 | 29870 | 3259356417 | 1293993 | 9100 | 8083275 |
| CVN-034_S3_L001_ | CVN-33 | NEGATIVE | FALSE | NOT_IDENTIFII | 911752 | 0 | 0 | 0 | 0 | 0 | 0 | 0 | 5 | VALID | 29869 | 0 | 29870 | 3259356417 | 536590 | 81 | 3228928 |
| CVN-035_S4_L001_ | CVN-34 | NEGATIVE | FALSE | NOT_IDENTIFII | 1113198 | 0 | 0 | 0 | 0 | 0 | 0 | 0 | 6 | VALID | 29869 | 0 | 29870 | 3259356417 | 769442 | 0 | 9434784 |
| CVN-036-1_S31_L0 | CVN-35_1 | NEGATIVE | FALSE | NOT_IDENTIFII | 1007799 | 0 | 0 | 0 | 0 | 0 | 0 | 0 | 4 | VALID | 29869 | 0 | 29870 | 3259356417 | 978437 | 0 | 27663021 |
| CVN-036-2_S5_L00 | CVN-35_2 | NEGATIVE | FALSE | NOT_IDENTIFII | 307574 | 0 | 0 | 0 | 0 | 0 | 0 | 0 | 6 | VALID | 29869 | 0 | 29870 | 3259356417 | 302883 | 0 | 6423988 |
| CVN-037_S6_L001_ | CVN-36 | NEGATIVE | FALSE | NOT_IDENTIFII | 390105 | 0 | 0 | 0 | 0 | 0 | 0 | 0 | 6 | VALID | 29869 | 0 | 29870 | 3259356417 | 325086 | 84 | 4773953 |
| CVN-038_S7_L001_ | CVN-37 | NEGATIVE | FALSE | NOT_IDENTIFII | 317484 | 0 | 0 | 0 | 0 | 0 | 0 | 0 | 5 | VALID | 29869 | 0 | 29870 | 3259356417 | 317040 | 0 | 2836082 |
| CVN-039_S8_L001_ | CVN-38 | NEGATIVE | FALSE | NOT_IDENTIFII | 1263795 | 0 | 0 | 0 | 0 | 0 | 0 | 0 | 6 | VALID | 29869 | 0 | 29870 | 3259356417 | 841854 | 0 | 2982735 |
| CVN-040_S9_L001_ | CVN-39 | NEGATIVE | FALSE | NOT_IDENTIFII | 736275 | 0 | 0 | 0 | 0 | 0 | 0 | 0 | 4 | VALID | 29869 | 0 | 29870 | 3259356417 | 566082 | 0 | 2058619 |
| CVN-041_S10_L001 | CVN-40 | NEGATIVE | FALSE | NOT_IDENTIFII | 3200532 | 0 | 0 | 0 | 0 | 0 | 0 | 0 | #NAME? | VALID | 29869 | 0 | 29870 | 3259356417 | 2918414 | 0 | 26239199 |
| CVN-042_S11_L001 | CVN-41 | NEGATIVE | FALSE | NOT_IDENTIFII | 694610 | 0 | 0 | 0 | 0 | 0 | 0 | 0 | #NAME? | VALID | 29869 | 0 | 29870 | 3259356417 | 626001 | 0 | 8915447 |
| CVN-043_S12_L001 | CVN-42 | NEGATIVE | FALSE | NOT_IDENTIFII | 624484 | 0 | 0 | 0 | 0 | 0 | 0 | 0 | #NAME? | VALID | 29869 | 0 | 29870 | 3259356417 | 578606 | 0 | 1517186 |
| CVN-044_S13_L001 | CVN-43 | NEGATIVE | FALSE | NOT_IDENTIFII | 324529 | 0 | 0 | 0 | 0 | 0 | 0 | 0 | #NAME? | VALID | 29869 | 0 | 29870 | 3259356417 | 266127 | 0 | 657999 |
| CVN-045_S14_L001 | CVN-44 | NEGATIVE | FALSE | NOT_IDENTIFII | 319133 | 0 | 0 | 0 | 0 | 0 | 0 | 0 | #NAME? | VALID | 29869 | 0 | 29870 | 3259356417 | 224207 | 0 | 766335 |
| CVN-046_S15_L001 | CVN-45 | NEGATIVE | FALSE | NOT_IDENTIFII | 291425 | 0 | 0 | 0 | 0 | 0 | 0 | 0 | #NAME? | VALID | 29869 | 0 | 29870 | 3259356417 | 239589 | 0 | 2071713 |
| CVN-047_S16_L001 | CVN-46 | NEGATIVE | FALSE | NOT_IDENTIFII | 3457456 | 0 | 0 | 0 | 0 | 0 | 0 | 0 | 8 | VALID | 29869 | 0 | 29870 | 3259356417 | 3452574 | 223 | 27735865 |
| CVN-048_S17_L001 | CVN-47 | NEGATIVE | FALSE | NOT_IDENTIFII | 1325097 | 0 | 0 | 0 | 0 | 0 | 0 | 0 | 6 | VALID | 29869 | 0 | 29870 | 3259356417 | 943011 | 168 | 4481973 |
| CVN-049_S18_L001 | CVN-48 | NEGATIVE | FALSE | NOT_IDENTIFII | 672352 | 0 | 0 | 0 | 0 | 0 | 0 | 0 | 4 | VALID | 29869 | 0 | 29870 | 3259356417 | 662707 | 0 | 16961962 |
| CVN-050_S19_L001 | CVN-49 | NEGATIVE | FALSE | NOT_IDENTIFII | 1143275 | 0 | 0 | 0 | 0 | 0 | 0 | 0 | #NAME? | VALID | 29869 | 0 | 29870 | 3259356417 | 828594 | 0 | 7241255 |
| CVN-051_S20_L001 | CVN-50 | NEGATIVE | FALSE | NOT_IDENTIFII | 730951 | 0 | 0 | 0 | 0 | 0 | 0 | 0 | 5 | VALID | 29869 | 0 | 29870 | 3259356417 | 465514 | 148 | 3566072 |
| CVN-052_S21_L001 | CVN-51 | NEGATIVE | FALSE | NOT_IDENTIFII | 3033444 | 0 | 0 | 0 | 0 | 0 | 0 | 0 | 5 | VALID | 29869 | 0 | 29870 | 3259356417 | 3030022 | 67 | 20622124 |
| CVN-053_S22_L001 | CVN-52 | NEGATIVE | FALSE | NOT_IDENTIFII | 1129558 | 0 | 0 | 0 | 0 | 0 | 0 | 0 | #NAME? | VALID | 29869 | 0 | 29870 | 3259356417 | 1127982 | 0 | 9535006 |
| CVN-054_S23_L001 | CVN-53 | NEGATIVE | FALSE | NOT_IDENTIFII | 532508 | 0 | 0 | 0 | 0 | 0 | 0 | 0 | #NAME? | VALID | 29869 | 0 | 29870 | 3259356417 | 518792 | 0 | 7713289 |
| CVN-055_S24_L001 | CVN-54 | NEGATIVE | FALSE | NOT_IDENTIFII | 689141 | 0 | 0 | 0 | 0 | 0 | 0 | 0 | 5 | VALID | 29869 | 0 | 29870 | 3259356417 | 551879 | 131 | 7320165 |
| CVN-056_S25_L001 | CVN-55 | NEGATIVE | FALSE | NOT_IDENTIFII | 1412365 | 0 | 0 | 0 | 0 | 0 | 0 | 0 | 7 | VALID | 29869 | 0 | 29870 | 3259356417 | 1084285 | 0 | 34628025 |
| CVN-057_S26_L001 | CVN-56 | NEGATIVE | FALSE | NOT_IDENTIFII | 841108 | 0 | 0 | 0 | 0 | 0 | 0 | 0 | #NAME? | VALID | 29869 | 0 | 29870 | 3259356417 | 792793 | 0 | 17373896 |
| CVN-058_S27_L001 | CVN-57 | NEGATIVE | FALSE | NOT_IDENTIFII | 854878 | 0 | 0 | 0 | 0 | 0 | 0 | 0 | 6 | VALID | 29869 | 0 | 29870 | 3259356417 | 853986 | 3829 | 12467062 |
| CVN-059_S28_L001 | CVN-58 | NEGATIVE | FALSE | NOT_IDENTIFII | 654913 | 0 | 0 | 0 | 0 | 0 | 0 | 0 | #NAME? | VALID | 29869 | 0 | 29870 | 3259356417 | 649394 | 0 | 11765876 |
| CVN-060_S29_L001 | CVN-59 | NEGATIVE | FALSE | NOT_IDENTIFII | 3178590 | 0 | 0 | 0 | 0 | 0 | 0 | 0 | 5 | VALID | 29869 | 0 | 29870 | 3259356417 | 3082478 | 60 | 14321919 |
| CVN-061_S30_L001 | CVN-60 | NEGATIVE | FALSE | NOT_IDENTIFII | 299431 | 0 | 0 | 0 | 0 | 0 | 0 | 0 | #NAME? | VALID | 29869 | 0 | 29870 | 3259356417 | 299140 | 0 | 2052778 |
| CVP-037_S32_L001_ | CVP-31 | POSITIVE | TRUE | 19A | 7934992 | 1 | 1 | 1 | 0 | 0 | 0 | 1 | 10 | VALID | 29869 | 0 | 29870 | 3259356417 | 5253950 | 1385048 | 10696251 |
| CVP-038_S33_L001_ | CVP-32 | POSITIVE | TRUE | 20A | 58496123 | 1 | 1 | 1 | 1 | 1 | 1 | 1 | 10 | VALID | 29869 | 1 | 29870 | 3259356417 | 49911604 | 6198541419 | 14209909 |
| CVP-039_S34_L001_ | CVP-33 | POSITIVE | TRUE | 20C | 1049210 | 1 | 1 | 1 | 1 | 1 | 0 | 1 | 10 | VALID | 29869 | 0 | 29870 | 3259356417 | 1010334 | 42403679 | 34281903 |
| CVP-040_S35_L001_ | CVP-34 | POSITIVE | TRUE | 20C | 800031 | 1 | 1 | 1 | 1 | 1 | 0 | 1 | 10 | VALID | 29869 | 1 | 29870 | 3259356417 | 779667 | 63493015 | 8702634 |
| CVP-041_S36_L001_ | CVP-35 | POSITIVE | TRUE | 20A | 1602710 | 1 | 1 | 1 | 1 | 1 | 0 | 1 | 10 | VALID | 29869 | 0 | 29870 | 3259356417 | 1219753 | 35726666 | 12543853 |
| CVP-042_S37_L001_ | CVP-36 | POSITIVE | TRUE | 20A | 2252087 | 1 | 1 | 1 | 1 | 1 | 0 | 1 | 10 | VALID | 29869 | 0 | 29870 | 3259356417 | 2067438 | 103173299 | 22084349 |
| CVP-043_S38_L001_ | CVP-37 | POSITIVE | TRUE | 20A | 4507123 | 1 | 1 | 1 | 1 | 1 | 0 | 1 | 10 | VALID | 29869 | 1 | 29870 | 3259356417 | 4503306 | 526532631 | 5097123 |
| CVP-044_S39_L001_ | CVP-38 | POSITIVE | TRUE | 20A | 4097802 | 1 | 1 | 1 | 1 | 1 | 0 | 1 | 10 | VALID | 29869 | 1 | 29870 | 3259356417 | 4095284 | 400413816 | 8293434 |
| CVP-045_S40_L001_ | CVP-39 | POSITIVE | TRUE | 20A | 705925 | 1 | 1 | 1 | 1 | 1 | 0 | 1 | 10 | VALID | 29869 | 0 | 29870 | 3259356417 | 691870 | 7082141 | 3813725 |
| CVP-046_S41_L001_ | CVP-40 | POSITIVE | TRUE | 19A | 935100 | 1 | 1 | 1 | 1 | 1 | 0 | 1 | 10 | VALID | 29869 | 1 | 29870 | 3259356417 | 902139 | 81213592 | 4071096 |
| CVP-047_S42_L001_ | CVP-41 | POSITIVE | FALSE | NOT_IDENTIFII | 557045 | 0 | 0 | 0 | 0 | 0 | 0 | 0 | 9 | VALID | 29869 | 0 | 29870 | 3259356417 | 514511 | 114949 | 9038227 |
| CVP-048_S43_L001_ | CVP-42 | POSITIVE | TRUE | 20C | 141438966 | 1 | 1 | 1 | 1 | 1 | 1 | 1 | 10 | VALID | 29869 | 1 | 29870 | 3259356417 | 49912226 | 6117593749 | 5336196 |
| CVP-049_S44_L001_ | CVP-43 | POSITIVE | TRUE | 20C | 116407395 | 1 | 1 | 1 | 1 | 1 | 1 | 1 | 10 | VALID | 29869 | 1 | 29870 | 3259356417 | 49173583 | 5965429667 | 12216323 |
| CVP-050_S45_L001_ | CVP-44 | POSITIVE | TRUE | 20A | 1129068 | 1 | 1 | 0 | 0 | 0 | 0 | 0 | 10 | VALID | 29869 | 0 | 29870 | 3259356417 | 1127536 | 583404 | 11883739 |
| CVP-051_S46_L001_ | CVP-45 | POSITIVE | TRUE | 19B | 726011 | 1 | 1 | 1 | 1 | 0 | 0 | 1 | 10 | VALID | 29869 | 0 | 29870 | 3259356417 | 563641 | 2743930 | 6370849 |
| CVP-052_S47_L001_ | CVP-46 | POSITIVE | TRUE | 20A | 917724 | 1 | 1 | 1 | 1 | 1 | 0 | 1 | 10 | VALID | 29869 | 0 | 29870 | 3259356417 | 701630 | 17158271 | 5157614 |
| CVP-053_S48_L001_ | CVP-47 | POSITIVE | TRUE | 19A | 399041 | 1 | 1 | 1 | 0 | 0 | 0 | 1 | 10 | VALID | 29869 | 0 | 29870 | 3259356417 | 363849 | 874650 | 2036881 |
| CVP-054_S49_L001_ | CVP-48 | POSITIVE | TRUE | 20C | 926795 | 1 | 1 | 1 | 1 | 1 | 0 | 1 | 10 | VALID | 29869 | 1 | 29870 | 3259356417 | 904391 | 72561910 | 5220193 |
| CVP-055_S50_L001_ | CVP-49 | POSITIVE | TRUE | 20C | 43366619 | 1 | 1 | 1 | 1 | 1 | 0 | 1 | 10 | VALID | 29869 | 1 | 29870 | 3259356417 | 43221935 | 5329788105 | 17291019 |
| CVP-056_S51_L001_ | CVP-50 | POSITIVE | TRUE | 20A | 3058620 | 1 | 1 | 1 | 1 | 1 | 0 | 1 | 10 | VALID | 29869 | 1 | 29870 | 3259356417 | 3055094 | 267046187 | 7969640 |
| CVP-057_S52_L001_ | CVP-51 | POSITIVE | TRUE | 20A | 1057279 | 1 | 1 | 1 | 1 | 1 | 0 | 1 | 10 | VALID | 29869 | 0 | 29870 | 3259356417 | 829267 | 7195136 | 5597056 |
| CVP-058_S53_L001_ | CVP-52 | POSITIVE | TRUE | 20A | 787803 | 1 | 1 | 1 | 1 | 1 | 0 | 1 | 10 | VALID | 29869 | 0 | 29870 | 3259356417 | 786327 | 18415321 | 20370353 |
| CVP-059_S54_L001_ | CVP-53 | POSITIVE | TRUE | 20C | 1470681 | 1 | 1 | 1 | 1 | 0 | 0 | 1 | 10 | VALID | 29869 | 0 | 29870 | 3259356417 | 870578 | 2898569 | 7170887 |
| CVP-060_S55_L001_ | CVP-54 | POSITIVE | TRUE | 20C | 8575143 | 1 | 1 | 1 | 1 | 1 | 0 | 1 | 10 | VALID | 29869 | 1 | 29870 | 3259356417 | 8561287 | 1000400795 | 3414156 |
| CVP-061_S56_L001_ | CVP-55 | POSITIVE | TRUE | 20C | 4262833 | 1 | 1 | 1 | 1 | 1 | 0 | 1 | 10 | VALID | 29869 | 1 | 29870 | 3259356417 | 4231828 | 441487651 | 7073977 |
| CVP-062_S57_L001_ | CVP-56 | POSITIVE | FALSE | NOT_IDENTIFII | 1215073 | 0 | 0 | 0 | 0 | 0 | 0 | 0 | 9 | VALID | 29869 | 0 | 29870 | 3259356417 | 1213440 | 87253 | 15206791 |
| CVP-063_S58_L001_ | CVP-57 | POSITIVE | TRUE | 19A | 3340360 | 1 | 0 | 0 | 0 | 0 | 0 | 0 | 10 | VALID | 29869 | 0 | 29870 | 3259356417 | 3337013 | 95419 | 30080017 |
| CVP-064_S59_L001_ | CVP-58 | POSITIVE | TRUE | 20C | 1582760 | 1 | 1 | 1 | 1 | 1 | 0 | 1 | 10 | VALID | 29869 | 1 | 29870 | 3259356417 | 1220553 | 83745222 | 4516113 |
| CVP-065_S60_L001_ | CVP-59 | POSITIVE | TRUE | 20C | 6012828 | 1 | 1 | 1 | 1 | 1 | 0 | 1 | 10 | VALID | 29869 | 1 | 29870 | 3259356417 | 6009518 | 663184278 | 20335132 |
| CVP-066_S61_L001_ | CVP-60 | POSITIVE | TRUE | 20A | 1054976 | 1 | 1 | 1 | 1 | 1 | 0 | 1 | 10 | VALID | 29869 | 0 | 29870 | 3259356417 | 898800 | 38402666 | 6181712 |
| Ctrl1_S68_L001_R1 | PC-1: Positive control | NA | FALSE | NOT_IDENTIFII | 698112 | 0 | 0 | 0 | 0 | 0 | 0 | 0 | 8 | VALID | 29869 | 0 | 29870 | 3259356417 | 697176 | 141673 | 2650189 |
| Ctrl2_S69_L001_R1 | PC-2: Positive control | NA | FALSE | NOT_IDENTIFII | 998649 | 0 | 0 | 0 | 0 | 0 | 0 | 0 | 6 | VALID | 29869 | 0 | 29870 | 3259356417 | 993104 | 33603 | 9175740 |
| HuRef-1_S66_L001_ | IC-1: Negative internal control | NA | FALSE | NOT_IDENTIFII | 698489 | 0 | 0 | 0 | 0 | 0 | 0 | 0 | 4 | VALID | 29869 | 0 | 29870 | 3259356417 | 691182 | 0 | 6368559 |
| HuRef-2_S67_L001_ | IC-2: Negative internal control | NA | FALSE | NOT_IDENTIFII | 517708 | 0 | 0 | 0 | 0 | 0 | 0 | 0 | 5 | VALID | 29869 | 0 | 29870 | 3259356417 | 517243 | 50 | 5200687 |
| VEROE6-1_S64_L00 | NEC-1: Negative extraction cor | NA | FALSE | NOT_IDENTIFII | 743360 | 0 | 0 | 0 | 0 | 0 | 0 | 0 | #NAME? | VALID | 29869 | 0 | 29870 | 3259356417 | 738055 | 0 | 648401 |
| VEROE6-2_S65_L00 | NEC-2: Negative extraction cor | NA | FALSE | NOT_IDENTIFII | 406201 | 0 | 0 | 0 | 0 | 0 | 0 | 0 | #NAME? | VALID | 29869 | 0 | 29870 | 3259356417 | 404166 | 0 | 253776 |
| VTM-1_S62_L001_F | NTC-1: Negative template cont | NA | FALSE | NOT_IDENTIFII | 1848 | 0 | 0 | 0 | 0 | 0 | 0 | 0 | #NAME? | VALID | 29869 | 0 | 29870 | 3259356417 | 669 | 0 | 2701 |
| VTM-2_S63_L001_F | NTC-2: Negative template cont | NA | FALSE | NOT_IDENTIFII | 625 | 0 | 0 | 0 | 0 | 0 | 0 | 0 | 6 | VALID | 29869 | 0 | 29870 | 3259356417 | 210 | 3299 | 2104 |

| Specimen_ID | Specimen_ID | TYPE_PCR | presence | clade | reads | X1 | X5 | X10 | X50 | X100 | X500 | E1 | I1 | validity | BAIT_TERRITO | PCT_SELECTED_B | TARGET_TERR | GENOME_SIZE | TOTAL_READS | ON_TARGET_E | HUMAN_ ON_TARGET_BASES |
|---|---|---|---|---|---|---|---|---|---|---|---|---|---|---|---|---|---|---|---|---|---|
| COVID-Pos-067_S8_ | CVP-61 | POSITIVE | TRUE | 20C | 33571630 | 1 | 1 | 1 | 1 | 1 | 0 | 1 | 10 | VALID | 29869 | 1 | 29870 | 3259356417 | 33432742 | 2150689165 | 1295204804 |
| COVID-Pos-068_S9_ | CVP-62 | POSITIVE | TRUE | 20C | 17780698 | 1 | 1 | 1 | 1 | 1 | 0 | 1 | 10 | VALID | 29869 | 1 | 29870 | 3259356417 | 17744075 | 1734427096 | 237910904 |
| COVID-Pos-069_S1 | CVP-63 | POSITIVE | TRUE | 19A | 1690892 | 1 | 1 | 0 | 0 | 0 | 0 | 0 | 10 | VALID | 29869 | 0 | 29870 | 3259356417 | 1687077 | 418481 | 126079525 |
| COVID-Pos-071_S1 | CVP-65 | POSITIVE | TRUE | 19A | 11036664 | 1 | 0 | 0 | 0 | 0 | 0 | 0 | 10 | VALID | 29869 | 0 | 29870 | 3259356417 | 11000217 | 23215 | 797636227 |
| COVID-Pos-072_S1 | CVP-66 | POSITIVE | TRUE | 20A | 10865511 | 1 | 1 | 1 | 1 | 1 | 0 | 1 | 10 | VALID | 29869 | 0 | 29870 | 3259356417 | 10829129 | 16824752 | 821795041 |
| COVID-Pos-073_S1 | CVP-67 | POSITIVE | TRUE | 20A | 20208866 | 1 | 1 | 1 | 1 | 1 | 0 | 1 | 10 | VALID | 29869 | 1 | 29870 | 3259356417 | 20172611 | 1608000009 | 493782111 |

| | | | | | | | | | | | | | | | | | | | | | |
|---|---|---|---|---|---|---|---|---|---|---|---|---|---|---|---|---|---|---|---|---|---|
| COVID-Pos-074_S14 | CVP-68 | POSITIVE | TRUE | 20A | 19788983 | 1 | 1 | 1 | 1 | 1 | 0 | 1 | 10 | VALID | 29869 | 0 | 29870 | 3259356417 | 19752696 | 753518427 | 1082554910 |
| COVID-Pos-076_S15 | CVP-70 | POSITIVE | TRUE | 19A | 9407581 | 1 | 0 | 0 | 0 | 0 | 0 | 0 | 10 | VALID | 29869 | 0 | 29870 | 3259356417 | 9357807 | 166720 | 739974962 |
| COVID-Pos-077_S16 | CVP-71 | POSITIVE | TRUE | 20A | 2205867 | 1 | 1 | 1 | 0 | 0 | 0 | 1 | 10 | VALID | 29869 | 0 | 29870 | 3259356417 | 2199945 | 566983 | 159934841 |
| COVID-Pos-078_S17 | CVP-72 | POSITIVE | FALSE | NOT_IDENTIFII | 13954504 | 0 | 0 | 0 | 0 | 0 | 0 | 0 | 8 | VALID | 29869 | 0 | 29870 | 3259356417 | 13925298 | 20530 | 1102343219 |
| COVID-Pos-079_S18 | CVP-73 | POSITIVE | TRUE | 20C | 18275102 | 1 | 1 | 1 | 1 | 1 | 0 | 1 | 10 | VALID | 29869 | 0 | 29870 | 3259356417 | 18211692 | 958393797 | 816679479 |
| COVID-Pos-080_S19 | CVP-74 | POSITIVE | TRUE | 20C | 11726362 | 1 | 1 | 1 | 1 | 0 | 0 | 1 | 10 | VALID | 29869 | 0 | 29870 | 3259356417 | 11664157 | 2221094 | 904594331 |
| COVID-Pos-081_S20 | CVP-75 | POSITIVE | TRUE | 20C | 31917334 | 1 | 1 | 1 | 1 | 1 | 0 | 1 | 10 | VALID | 29869 | 0 | 29870 | 3259356417 | 31857965 | 1865250906 | 1247675215 |
| COVID-Pos-083_S21 | CVP-77 | POSITIVE | TRUE | 20C | 69100764 | 1 | 1 | 1 | 1 | 1 | 0 | 1 | 10 | VALID | 29869 | 1 | 29870 | 3259356417 | 49881888 | 4871795517 | 845963785 |
| COVID-Pos-084_S22 | CVP-78 | POSITIVE | TRUE | 20B | 14829910 | 1 | 1 | 1 | 0 | 0 | 0 | 1 | 10 | VALID | 29869 | 0 | 29870 | 3259356417 | 14795608 | 788732 | 1117665326 |
| COVID-Pos-085_S23 | CVP-79 | POSITIVE | TRUE | 20A | 67594318 | 1 | 1 | 1 | 1 | 1 | 1 | 1 | 10 | VALID | 29869 | 1 | 29870 | 3259356417 | 49860131 | 5515803494 | 369420377 |
| COVID-Pos-086_S7_ | CVP-80 | POSITIVE | FALSE | NOT_IDENTIFII | 1654901 | 0 | 0 | 0 | 0 | 0 | 0 | 0 | 8 | VALID | 29869 | 0 | 29870 | 3259356417 | 1651134 | 12638 | 116644448 |
| HuRef-1_S5_L001_F | IC-1: Negative internal control | NA | TRUE | NOT_IDENTIFII | 35512912 | 0 | 0 | 0 | 0 | 0 | 0 | 0 | 9 | VALID | 29869 | 0 | 29870 | 3259356417 | 35394199 | 7094 | 262159671 |
| HuRef-2_S6_L001_F | IC-2: Negative internal control | NA | FALSE | NOT_IDENTIFII | 26535394 | 0 | 0 | 0 | 0 | 0 | 0 | 0 | 7 | VALID | 29869 | 0 | 29870 | 3259356417 | 26450317 | 58 | 296239480 |
| Positive-1_S26_L00 | PC-1: Positive control | NA | TRUE | 19A | 35122364 | 1 | 1 | 1 | 0 | 0 | 0 | 1 | 10 | VALID | 29869 | 0 | 29870 | 3259356417 | 35015304 | 364062 | 266921741 |
| Positive-2_S27_L00 | PC-2: Positive control | NA | TRUE | 19A | 31556953 | 1 | 1 | 1 | 0 | 0 | 0 | 1 | 10 | VALID | 29869 | 0 | 29870 | 3259356417 | 31400028 | 621482 | 196638925 |
| VTM-1_S1_L001_R1 | NTC-1: Negative template cont | NA | FALSE | NOT_IDENTIFII | 189821 | 0 | 0 | 0 | 0 | 0 | 0 | 0 | 8 | VALID | 29869 | 0 | 29870 | 3259356417 | 187676 | 0 | 11720 |
| VTM-2_S2_L001_R1 | NTC-2: Negative template cont | NA | FALSE | NOT_IDENTIFII | 15268 | 0 | 0 | 0 | 0 | 0 | 0 | 0 | 6 | VALID | 29869 | 0 | 29870 | 3259356417 | 14492 | 0 | 4094 |
| VeroE6-1_S3_L001_ | NEC-1: Negative extraction cor | NA | FALSE | NOT_IDENTIFII | 25396952 | 0 | 0 | 0 | 0 | 0 | 0 | 0 | 8 | VALID | 29869 | 0 | 29870 | 3259356417 | 23190789 | 191 | 80498653 |
| VeroE6-2_S4_L001_ | NEC-2: Negative extraction cor | NA | TRUE | NOT_IDENTIFII | 31496180 | 0 | 0 | 0 | 0 | 0 | 0 | 0 | 10 | VALID | 29869 | 0 | 29870 | 3259356417 | 28498542 | 1764 | 98747661 |

**Subsampling data.** We subsampled each of the four FASTQs down to a maximum of 125,000 reads totaling up to 500,000 reads for the full set of four FASTQs per sample. If a FASTQ had fewer than 125,000 reads, it was not

| Specimen_ID | Specimen_ID | TYPE_PCR | presence | clade | reads | X1 | X5 | X10 | X50 | X100 | X500 | E1 | I1 | | validity | BAIT_TERRITO | PCT_SELECTED_B | TARGET_TERR | GENOME_SIZE | TOTAL_READS | HUMAN_ | ON_TARGET_BASES |
|---|---|---|---|---|---|---|---|---|---|---|---|---|---|---|---|---|---|---|---|---|---|---|
| COVID-NEG-002_S6 | CVN-1 | NEGATIVE | FALSE | NOT_IDENTIFII | 2473492 | 0 | 0 | 0 | 0 | 0 | 0 | 0 | 5 | VALID | 29869 | 0 | 29870 | 3259356417 | 498924 | 45782452 | |
| COVID-NEG-003_S7 | CVN-2 | NEGATIVE | FALSE | NOT_IDENTIFII | 3252564 | 0 | 0 | 0 | 0 | 0 | 0 | 0 | #NAME? | VALID | 29869 | 0 | 29870 | 3259356417 | 499282 | 45775822 | |
| COVID-NEG-005_S9 | CVN-4 | NEGATIVE | FALSE | NOT_IDENTIFII | 10801026 | 0 | 0 | 0 | 0 | 0 | 0 | 0 | 7 | VALID | 29869 | 0 | 29870 | 3259356417 | 498903 | 17264995 | |
| COVID-NEG-006_S1 | CVN-5 | NEGATIVE | FALSE | NOT_IDENTIFII | 2061968 | 0 | 0 | 0 | 0 | 0 | 0 | 0 | 6 | VALID | 29869 | 0 | 29870 | 3259356417 | 497977 | 31492807 | |
| COVID-NEG-007_S3 | CVN-6 | NEGATIVE | FALSE | NOT_IDENTIFII | 1064413 | 0 | 0 | 0 | 0 | 0 | 0 | 0 | 8 | VALID | 29869 | 0 | 29870 | 3259356417 | 496552 | 33314243 | |
| COVID-NEG-008_S3 | CVN-7 | NEGATIVE | FALSE | NOT_IDENTIFII | 2357502 | 0 | 0 | 0 | 0 | 0 | 0 | 0 | 6 | VALID | 29869 | 0 | 29870 | 3259356417 | 491566 | 51012 | |
| COVID-NEG-009_S3 | CVN-8 | NEGATIVE | FALSE | NOT_IDENTIFII | 2896238 | 0 | 0 | 0 | 0 | 0 | 0 | 0 | 5 | VALID | 29869 | 0 | 29870 | 3259356417 | 499530 | 21038611 | |
| COVID-NEG-010_S3 | CVN-9 | NEGATIVE | FALSE | NOT_IDENTIFII | 4037293 | 0 | 0 | 0 | 0 | 0 | 0 | 0 | 6 | VALID | 29869 | 0 | 29870 | 3259356417 | 499591 | 19370288 | |
| COVID-NEG-013_S4 | CVN-10 | NEGATIVE | FALSE | NOT_IDENTIFII | 1814277 | 0 | 0 | 0 | 0 | 0 | 0 | 0 | 8 | VALID | 29869 | 0 | 29870 | 3259356417 | 499473 | 39233441 | |
| COVID-NEG-014_S4 | CVN-11 | NEGATIVE | FALSE | NOT_IDENTIFII | 49438 | 0 | 0 | 0 | 0 | 0 | 0 | 0 | 7 | VALID | 29869 | 0 | 29870 | 3259356417 | 49344 | 3017714 | |
| COVID-NEG-015_S4 | CVN-12 | NEGATIVE | FALSE | NOT_IDENTIFII | 75993 | 0 | 0 | 0 | 0 | 0 | 0 | 0 | 8 | VALID | 29869 | 0 | 29870 | 3259356417 | 75797 | 4176197 | |
| COVID-NEG-016_S4 | CVN-13 | NEGATIVE | FALSE | NOT_IDENTIFII | 62897 | 0 | 0 | 0 | 0 | 0 | 0 | 0 | 7 | VALID | 29869 | 0 | 29870 | 3259356417 | 62444 | 4525370 | |
| COVID-NEG-017_S4 | CVN-14 | NEGATIVE | FALSE | NOT_IDENTIFII | 68069 | 0 | 0 | 0 | 0 | 0 | 0 | 0 | 7 | VALID | 29869 | 0 | 29870 | 3259356417 | 67913 | 5251566 | |
| COVID-NEG-018_S4 | CVN-15 | NEGATIVE | FALSE | NOT_IDENTIFII | 1580183 | 0 | 0 | 0 | 0 | 0 | 0 | 0 | 6 | VALID | 29869 | 0 | 29870 | 3259356417 | 499261 | 22421086 | |
| COVID-NEG-019_S4 | CVN-16 | NEGATIVE | FALSE | NOT_IDENTIFII | 1110690 | 0 | 0 | 0 | 0 | 0 | 0 | 0 | #NAME? | VALID | 29869 | 0 | 29870 | 3259356417 | 499158 | 43314118 | |
| COVID-NEG-022_S5 | CVN-17 | NEGATIVE | FALSE | NOT_IDENTIFII | 0 | 0 | 0 | 0 | 0 | 0 | 0 | 0 | #NAME? | INVALID | 29869 | ? | 29870 | 3259356417 | 0 | 0 | |
| COVID-NEG-023_S5 | CVN-18 | NEGATIVE | FALSE | NOT_IDENTIFII | 210499 | 0 | 0 | 0 | 0 | 0 | 0 | 0 | 7 | VALID | 29869 | 0 | 29870 | 3259356417 | 209358 | 13428920 | |
| COVID-NEG-024_S5 | CVN-19 | NEGATIVE | FALSE | NOT_IDENTIFII | 235757 | 0 | 0 | 0 | 0 | 0 | 0 | 0 | #NAME? | VALID | 29869 | 0 | 29870 | 3259356417 | 235196 | 17213687 | |
| COVID-NEG-026_S5 | CVN-20 | NEGATIVE | FALSE | NOT_IDENTIFII | 53559 | 0 | 0 | 0 | 0 | 0 | 0 | 0 | 7 | VALID | 29869 | 0 | 29870 | 3259356417 | 52490 | 1753667 | |
| COVID-NEG-027_S5 | CVN-21 | NEGATIVE | FALSE | NOT_IDENTIFII | 62180 | 0 | 0 | 0 | 0 | 0 | 0 | 0 | #NAME? | VALID | 29869 | 0 | 29870 | 3259356417 | 62023 | 3964318 | |
| COVID-NEG-028_S5 | CVN-22 | NEGATIVE | FALSE | NOT_IDENTIFII | 67953 | 0 | 0 | 0 | 0 | 0 | 0 | 0 | 6 | VALID | 29869 | 0 | 29870 | 3259356417 | 67814 | 3865381 | |
| COVID-NEG-029_S5 | CVN-23 | NEGATIVE | FALSE | NOT_IDENTIFII | 64494 | 0 | 0 | 0 | 0 | 0 | 0 | 0 | 4 | VALID | 29869 | 0 | 29870 | 3259356417 | 64331 | 3402919 | |
| COVID-NEG-030_S5 | CVN-24 | NEGATIVE | FALSE | NOT_IDENTIFII | 131360 | 0 | 0 | 0 | 0 | 0 | 0 | 0 | #NAME? | VALID | 29869 | 0 | 29870 | 3259356417 | 131215 | 9137168 | |
| COVID-NEG-031_S5 | CVN-25 | NEGATIVE | FALSE | NOT_IDENTIFII | 90980 | 0 | 0 | 0 | 0 | 0 | 0 | 0 | 5 | VALID | 29869 | 0 | 29870 | 3259356417 | 86763 | 4071720 | |
| COVID-Neg-004_S4 | CVN-26 | NEGATIVE | FALSE | NOT_IDENTIFII | 2288625 | 0 | 0 | 0 | 0 | 0 | 0 | 0 | 4 | VALID | 29869 | 0 | 29870 | 3259356417 | 492215 | 26581444 | |
| COVID-Neg-011_S4 | CVN-27 | NEGATIVE | FALSE | NOT_IDENTIFII | 1730315 | 0 | 0 | 0 | 0 | 0 | 0 | 0 | #NAME? | VALID | 29869 | 0 | 29870 | 3259356417 | 499665 | 41549069 | |
| COVID-Neg-020_S4 | CVN-28 | NEGATIVE | FALSE | NOT_IDENTIFII | 2915736 | 0 | 0 | 0 | 0 | 0 | 0 | 0 | 5 | VALID | 29869 | 0 | 29870 | 3259356417 | 499707 | 34849949 | |
| COVID-Neg-021_S4 | CVN-29 | NEGATIVE | FALSE | NOT_IDENTIFII | 2887705 | 0 | 0 | 0 | 0 | 0 | 0 | 0 | #NAME? | VALID | 29869 | 0 | 29870 | 3259356417 | 495031 | 36176839 | |
| COVID-Neg-025_S4 | CVN-30 | NEGATIVE | FALSE | NOT_IDENTIFII | 1098581 | 0 | 0 | 0 | 0 | 0 | 0 | 0 | 8 | VALID | 29869 | 0 | 29870 | 3259356417 | 496906 | 42048978 | |
| COVID-POS-006_S1 | CVP-1 | POSITIVE | TRUE | 20A | 3716751 | 1 | 1 | 1 | 1 | 1 | 0 | 1 | 10 | VALID | 29869 | 0 | 29870 | 3259356417 | 499536 | 31036324 | |
| COVID-POS-007_S2 | CVP-2 | POSITIVE | TRUE | 20C | 5376358 | 1 | 1 | 1 | 1 | 1 | 0 | 1 | 10 | VALID | 29869 | 0 | 29870 | 3259356417 | 498995 | 32404111 | |
| COVID-POS-008_S3 | CVP-3 | POSITIVE | TRUE | 20A | 5422292 | 1 | 1 | 1 | 1 | 1 | 0 | 1 | 10 | VALID | 29869 | 0 | 29870 | 3259356417 | 499591 | 23259493 | |
| COVID-POS-009_S4 | CVP-4 | POSITIVE | TRUE | 19A | 3465259 | 1 | 0 | 0 | 0 | 0 | 0 | 1 | 10 | VALID | 29869 | 0 | 29870 | 3259356417 | 499242 | 29062216 | |
| COVID-POS-010_S5 | CVP-5 | POSITIVE | TRUE | 19A | 1256389 | 1 | 1 | 1 | 0 | 0 | 0 | 1 | 10 | VALID | 29869 | 0 | 29870 | 3259356417 | 499472 | 29990092 | |
| COVID-POS-011_S1 | CVP-6 | POSITIVE | TRUE | 20C | 80637571 | 1 | 1 | 1 | 1 | 1 | 0 | 1 | 10 | VALID | 29869 | 1 | 29870 | 3259356417 | 499515 | 1794022 | |
| COVID-POS-012_S1 | CVP-7 | POSITIVE | FALSE | NOT_IDENTIFII | 730999 | 0 | 0 | 0 | 0 | 0 | 0 | 0 | 7 | VALID | 29869 | 0 | 29870 | 3259356417 | 499596 | 42860237 | |
| COVID-POS-013_S1 | CVP-8 | POSITIVE | TRUE | 20C | 778532 | 1 | 1 | 1 | 1 | 1 | 0 | 1 | 10 | VALID | 29869 | 0 | 29870 | 3259356417 | 497402 | 30854625 | |
| COVID-POS-014_S1 | CVP-9 | POSITIVE | TRUE | 20C | 6660390 | 1 | 1 | 1 | 1 | 1 | 0 | 1 | 10 | VALID | 29869 | 0 | 29870 | 3259356417 | 499620 | 14145951 | |
| COVID-POS-015_S1 | CVP-10 | POSITIVE | TRUE | 20C | 5737861 | 1 | 1 | 1 | 1 | 0 | 0 | 1 | 10 | VALID | 29869 | 0 | 29870 | 3259356417 | 498640 | 32129960 | |
| COVID-POS-016_S1 | CVP-11 | POSITIVE | TRUE | 20A | 6243585 | 1 | 1 | 1 | 1 | 1 | 0 | 1 | 10 | VALID | 29869 | 1 | 29870 | 3259356417 | 499470 | 7608737 | |
| COVID-POS-017_S1 | CVP-12 | POSITIVE | TRUE | 20C | 14308108 | 1 | 1 | 1 | 1 | 1 | 0 | 1 | 10 | VALID | 29869 | 1 | 29870 | 3259356417 | 499649 | 2780437 | |
| COVID-POS-018_S1 | CVP-13 | POSITIVE | TRUE | 20C | 158979 | 1 | 1 | 1 | 1 | 1 | 0 | 1 | 10 | VALID | 29869 | 1 | 29870 | 3259356417 | 158739 | 4784065 | |
| COVID-POS-019_S1 | CVP-14 | POSITIVE | TRUE | NOT_IDENTIFII | 68212 | 0 | 0 | 0 | 0 | 0 | 0 | 0 | 10 | VALID | 29869 | 0 | 29870 | 3259356417 | 68032 | 4206422 | |
| COVID-POS-020_S2 | CVP-15 | POSITIVE | TRUE | 20A | 60695705 | 1 | 1 | 1 | 1 | 1 | 0 | 1 | 10 | VALID | 29869 | 1 | 29870 | 3259356417 | 499554 | 148937 | |
| COVID-POS-021_S2 | CVP-16 | POSITIVE | TRUE | 19A | 244330 | 1 | 1 | 1 | 1 | 1 | 0 | 1 | 10 | VALID | 29869 | 0 | 29870 | 3259356417 | 244087 | 4964282 | |
| COVID-POS-022_S2 | CVP-17 | POSITIVE | TRUE | 19A | 1178790 | 1 | 1 | 1 | 0 | 0 | 0 | 1 | 10 | VALID | 29869 | 0 | 29870 | 3259356417 | 499335 | 41931588 | |
| COVID-POS-023_S2 | CVP-18 | POSITIVE | TRUE | 20A | 1239946 | 1 | 1 | 1 | 1 | 1 | 0 | 1 | 10 | VALID | 29869 | 0 | 29870 | 3259356417 | 492472 | 32919126 | |
| COVID-POS-024_S2 | CVP-19 | POSITIVE | TRUE | 20A | 24316832 | 1 | 1 | 1 | 1 | 1 | 0 | 1 | 10 | VALID | 29869 | 1 | 29870 | 3259356417 | 499688 | 2763928 | |
| COVID-POS-025_S2 | CVP-20 | POSITIVE | TRUE | 19A | 1255877 | 1 | 1 | 1 | 0 | 0 | 0 | 1 | 10 | VALID | 29869 | 0 | 29870 | 3259356417 | 499045 | 44827793 | |
| COVID-POS-026_S2 | CVP-21 | POSITIVE | TRUE | 20A | 239196 | 1 | 1 | 1 | 1 | 0 | 0 | 1 | 10 | VALID | 29869 | 0 | 29870 | 3259356417 | 238715 | 16427173 | |
| COVID-POS-027_S2 | CVP-22 | POSITIVE | TRUE | 20A | 33357673 | 1 | 1 | 1 | 1 | 1 | 0 | 1 | 10 | VALID | 29869 | 1 | 29870 | 3259356417 | 499612 | 370017 | |
| COVID-POS-028_S6 | CVP-23 | POSITIVE | TRUE | 20C | 63104 | 1 | 1 | 1 | 1 | 1 | 0 | 1 | 10 | VALID | 29869 | 1 | 29870 | 3259356417 | 62936 | 885938 | |
| COVID-POS-029_S2 | CVP-24 | POSITIVE | TRUE | 20C | 13271148 | 1 | 1 | 1 | 1 | 1 | 0 | 1 | 10 | VALID | 29869 | 1 | 29870 | 3259356417 | 497364 | 834162 | |
| COVID-POS-030_S2 | CVP-25 | POSITIVE | TRUE | 20C | 1738765 | 1 | 1 | 1 | 1 | 1 | 0 | 1 | 10 | VALID | 29869 | 1 | 29870 | 3259356417 | 499707 | 1598091 | |
| COVID-POS-031_S3 | CVP-26 | POSITIVE | TRUE | 19A | 77888 | 1 | 1 | 1 | 0 | 0 | 0 | 1 | 10 | VALID | 29869 | 0 | 29870 | 3259356417 | 77780 | 5159470 | |
| COVID-POS-032_S3 | CVP-27 | POSITIVE | TRUE | 19A | 117370 | 1 | 1 | 1 | 0 | 0 | 0 | 1 | 10 | VALID | 29869 | 0 | 29870 | 3259356417 | 117124 | 6341409 | |
| COVID-POS-033_S3 | CVP-28 | POSITIVE | TRUE | 20B | 53540594 | 1 | 1 | 1 | 1 | 1 | 0 | 1 | 10 | VALID | 29869 | 1 | 29870 | 3259356417 | 499609 | 291486 | |
| COVID-POS-034_S3 | CVP-29 | POSITIVE | TRUE | 19A | 102921 | 1 | 0 | 0 | 0 | 0 | 0 | 0 | 10 | VALID | 29869 | 0 | 29870 | 3259356417 | 102661 | 6979518 | |
| COVID-POS-035_S3 | CVP-30 | POSITIVE | TRUE | 19A | 113558 | 1 | 1 | 1 | 0 | 0 | 0 | 1 | 10 | VALID | 29869 | 0 | 29870 | 3259356417 | 113339 | 7062458 | |
| Ctrl1-1e4_S63_L001 | PC-1: Positive control (10,000 ( | NA | TRUE | 19A | 503319 | 1 | 1 | 1 | 1 | 1 | 0 | 1 | 10 | VALID | 29869 | 0 | 29870 | 3259356417 | 498737 | 3949388 | |
| Ctrl2-1e6-1_S61_L0 | PC-2: Positive control (1,000,0( | NA | TRUE | 19A | 47012721 | 1 | 1 | 1 | 1 | 1 | 0 | 1 | 10 | VALID | 29869 | 1 | 29870 | 3259356417 | 499601 | 36296 | |
| Ctrl2-1e6-2_S62_L0 | PC-3: Positive control (1,000,0( | NA | TRUE | 19A | 42266116 | 1 | 1 | 1 | 1 | 1 | 0 | 1 | 10 | VALID | 29869 | 1 | 29870 | 3259356417 | 499391 | 2262938 | |
| HuRef-1_S64_L001_ | IC-1: Negative internal control | NA | FALSE | NOT_IDENTIFII | 352565 | 0 | 0 | 0 | 0 | 0 | 0 | 0 | 5 | VALID | 29869 | 0 | 29870 | 3259356417 | 351931 | 2140243 | |
| HuRef-2_S65_L001_ | IC-2: Negative internal control | NA | FALSE | NOT_IDENTIFII | 10516002 | 0 | 0 | 0 | 0 | 0 | 0 | 0 | #NAME? | VALID | 29869 | 0 | 29870 | 3259356417 | 499346 | 3896584 | |
| NC-1_S66_L001_R1 | NTC-1: Negative template cont | NA | FALSE | NOT_IDENTIFII | 61 | 0 | 0 | 0 | 0 | 0 | 0 | 0 | #NAME? | VALID | 29869 | 0 | 29870 | 3259356417 | 20 | 544 | |
| NC-2_S67_L001_R1 | NTC-2: Negative template cont | NA | FALSE | NOT_IDENTIFII | 1552 | 0 | 0 | 0 | 0 | 0 | 0 | 0 | 5 | VALID | 29869 | 0 | 29870 | 3259356417 | 833 | 20909 | |

**Supplemental Table 3.** Specimen information of 140 samples used in the validation study, independent validation and geographic study with specification on the collection device, viral transport media, the comparator RT-PCR methodology, results and mean Ct value.

*VTM: viral transport media; NA: we were not able to identify the collection device and matrix due to the label damage during de-identification of samples.*

| Specimen ID | Specimen ID | Collection Device | Matrix (VTM) | Study | Collection Date | Location (State) | Comparator RT-PCR Results | Comparator RT-PCR Methodology | Mean_Ct_value using GenArraytion | Mean_Ct_value using Roche Cobas (ORF1 gene) | Mean_Ct_value using Roche Cobas (E gene) |
|---|---|---|---|---|---|---|---|---|---|---|---|
| COVID-NEG-002 | CVN-1 | BD | UTM | Validation Study | April | New York | NEGATIVE | Panther Fusion SARS-CoV-2 Assay (Hologic) RT-PCR testing | Undetermined | NA | NA |
| COVID-NEG-003 | CVN-2 | PURITAN | UTM | Validation Study | April | New York | NEGATIVE | Panther Fusion SARS-CoV-2 Assay (Hologic) RT-PCR testing | Undetermined | NA | NA |
| COVID-NEG-004 | CVN-3 | STARPLEX | MULTITRANS | Validation Study | April | New York | NEGATIVE | Panther Fusion SARS-CoV-2 Assay (Hologic) RT-PCR testing | Not enough sample | NA | NA |
| COVID-NEG-005 | CVN-4 | PURITAN | UTM | Validation Study | April | New York | NEGATIVE | Panther Fusion SARS-CoV-2 Assay (Hologic) RT-PCR testing | Not enough sample | NA | NA |
| COVID-NEG-006 | CVN-5 | PURITAN | UTM | Validation Study | April | New York | NEGATIVE | Panther Fusion SARS-CoV-2 Assay (Hologic) RT-PCR testing | Undetermined | NA | NA |
| COVID-NEG-007 | CVN-6 | COPAN | UTM | Validation Study | April | New York | NEGATIVE | Panther Fusion SARS-CoV-2 Assay (Hologic) RT-PCR testing | Undetermined | NA | NA |
| COVID-NEG-008 | CVN-7 | COPAN | UTM | Validation Study | April | New York | NEGATIVE | Panther Fusion SARS-CoV-2 Assay (Hologic) RT-PCR testing | Not enough sample | NA | NA |
| COVID-NEG-009 | CVN-8 | STARPLEX | MULTITRANS | Validation Study | April | New York | NEGATIVE | Panther Fusion SARS-CoV-2 Assay (Hologic) RT-PCR testing | Not enough sample | NA | NA |
| COVID-NEG-010 | CVN-9 | PURITAN | UTM | Validation Study | April | New York | NEGATIVE | Panther Fusion SARS-CoV-2 Assay (Hologic) RT-PCR testing | Undetermined | NA | NA |
| COVID-NEG-011 | CVN-10 | BD | UTM | Validation Study | April | New York | NEGATIVE | Panther Fusion SARS-CoV-2 Assay (Hologic) RT-PCR testing | Undetermined | NA | NA |
| COVID-NEG-012 | CVN-11 | PURITAN | UTM | Validation Study | April | New York | NEGATIVE | Panther Fusion SARS-CoV-2 Assay (Hologic) RT-PCR testing | Undetermined | NA | NA |
| COVID-NEG-013 | CVN-12 | BD | UTM | Validation Study | April | New York | NEGATIVE | Panther Fusion SARS-CoV-2 Assay (Hologic) RT-PCR testing | Undetermined | NA | NA |
| COVID-NEG-014 | CVN-13 | NA | NA | Validation Study | April | New York | NEGATIVE | Panther Fusion SARS-CoV-2 Assay (Hologic) RT-PCR testing | Undetermined | NA | NA |
| COVID-NEG-015 | CVN-14 | STARPLEX | MULTITRANS | Validation Study | April | New York | NEGATIVE | Panther Fusion SARS-CoV-2 Assay (Hologic) RT-PCR testing | Undetermined | NA | NA |
| COVID-NEG-016 | CVN-15 | COPAN | UTM | Validation Study | April | New York | NEGATIVE | Panther Fusion SARS-CoV-2 Assay (Hologic) RT-PCR testing | Undetermined | NA | NA |
| COVID-NEG-017 | CVN-16 | COPAN | UTM | Validation Study | April | New York | NEGATIVE | Panther Fusion SARS-CoV-2 Assay (Hologic) RT-PCR testing | Undetermined | NA | NA |
| COVID-NEG-018 | CVN-17 | BD | UTM | Validation Study | April | New York | NEGATIVE | Panther Fusion SARS-CoV-2 Assay (Hologic) RT-PCR testing | Undetermined | NA | NA |
| COVID-NEG-019 | CVN-18 | BD | UTM | Validation Study | April | New York | NEGATIVE | Panther Fusion SARS-CoV-2 Assay (Hologic) RT-PCR testing | Undetermined | NA | NA |
| COVID-NEG-020 | CVN-19 | BD | UTM | Validation Study | April | New York | NEGATIVE | Panther Fusion SARS-CoV-2 Assay (Hologic) RT-PCR testing | Undetermined | NA | NA |
| COVID-NEG-021 | CVN-20 | PURITAN | UTM | Validation Study | April | New York | NEGATIVE | Panther Fusion SARS-CoV-2 Assay (Hologic) RT-PCR testing | Undetermined | NA | NA |
| COVID-NEG-022 | CVN-21 | BD | UTM | Validation Study | April | New York | NEGATIVE | Panther Fusion SARS-CoV-2 Assay (Hologic) RT-PCR testing | Undetermined | NA | NA |
| COVID-NEG-023 | CVN-22 | BD | UTM | Validation Study | April | New York | NEGATIVE | Panther Fusion SARS-CoV-2 Assay (Hologic) RT-PCR testing | Undetermined | NA | NA |
| COVID-NEG-024 | CVN-23 | STARPLEX | MULTITRANS | Validation Study | April | New York | NEGATIVE | Panther Fusion SARS-CoV-2 Assay (Hologic) RT-PCR testing | Undetermined | NA | NA |
| COVID-NEG-025 | CVN-24 | BD | UTM | Validation Study | April | New York | NEGATIVE | Panther Fusion SARS-CoV-2 Assay (Hologic) RT-PCR testing | Undetermined | NA | NA |
| COVID-NEG-026 | CVN-25 | BD | UTM | Validation Study | April | New York | NEGATIVE | Panther Fusion SARS-CoV-2 Assay (Hologic) RT-PCR testing | Undetermined | NA | NA |
| COVID-NEG-027 | CVN-26 | BD | UTM | Validation Study | April | New York | NEGATIVE | Panther Fusion SARS-CoV-2 Assay (Hologic) RT-PCR testing | Undetermined | NA | NA |
| COVID-NEG-028 | CVN-27 | BD | UTM | Validation Study | April | New York | NEGATIVE | Panther Fusion SARS-CoV-2 Assay (Hologic) RT-PCR testing | Undetermined | NA | NA |
| COVID-NEG-029 | CVN-28 | BD | UTM | Validation Study | April | New York | NEGATIVE | Panther Fusion SARS-CoV-2 Assay (Hologic) RT-PCR testing | Undetermined | NA | NA |
| COVID-NEG-030 | CVN-29 | BD | UTM | Validation Study | April | New York | NEGATIVE | Panther Fusion SARS-CoV-2 Assay (Hologic) RT-PCR testing | Undetermined | NA | NA |
| COVID-NEG-031 | CVN-30 | BD | UTM | Validation Study | April | New York | NEGATIVE | Panther Fusion SARS-CoV-2 Assay (Hologic) RT-PCR testing | Undetermined | NA | NA |
| COVID-POS-006 | CVP-1 | REMEL M6 | M6 | Validation Study | April | New York | POSITIVE | Panther Fusion SARS-CoV-2 Assay (Hologic) RT-PCR testing | 27.230 | NA | NA |
| COVID-POS-007 | CVP-2 | COPAN | UTM | Validation Study | April | New York | POSITIVE | Panther Fusion SARS-CoV-2 Assay (Hologic) RT-PCR testing | 24.713 | NA | NA |
| COVID-POS-008 | CVP-3 | REMEL M6 | M6 | Validation Study | April | New York | POSITIVE | Panther Fusion SARS-CoV-2 Assay (Hologic) RT-PCR testing | 26.727 | NA | NA |
| COVID-POS-009 | CVP-4 | REMEL M6 | M6 | Validation Study | April | New York | POSITIVE | Panther Fusion SARS-CoV-2 Assay (Hologic) RT-PCR testing | 31.122 | NA | NA |
| COVID-POS-010 | CVP-5 | BD | UTM | Validation Study | April | New York | POSITIVE | Panther Fusion SARS-CoV-2 Assay (Hologic) RT-PCR testing | 29.452 | NA | NA |
| COVID-POS-011 | CVP-6 | STARPLEX | MULTITRANS | Validation Study | April | New York | POSITIVE | Panther Fusion SARS-CoV-2 Assay (Hologic) RT-PCR testing | 15.660 | NA | NA |
| COVID-POS-012 | CVP-7 | BD | UTM | Validation Study | April | New York | POSITIVE | Panther Fusion SARS-CoV-2 Assay (Hologic) RT-PCR testing | Undetermined | NA | NA |
| COVID-POS-013 | CVP-8 | BD | UTM | Validation Study | April | New York | POSITIVE | Panther Fusion SARS-CoV-2 Assay (Hologic) RT-PCR testing | Undetermined | NA | NA |
| COVID-POS-014 | CVP-9 | BD | UTM | Validation Study | April | New York | POSITIVE | Panther Fusion SARS-CoV-2 Assay (Hologic) RT-PCR testing | 24.863 | NA | NA |
| COVID-POS-015 | CVP-10 | COPAN | UTM | Validation Study | April | New York | POSITIVE | Panther Fusion SARS-CoV-2 Assay (Hologic) RT-PCR testing | 33.331 | NA | NA |
| COVID-POS-016 | CVP-11 | COPAN | UTM | Validation Study | April | New York | POSITIVE | Panther Fusion SARS-CoV-2 Assay (Hologic) RT-PCR testing | Undetermined | NA | NA |
| COVID-POS-017 | CVP-12 | REMEL M6 | M6 | Validation Study | April | New York | POSITIVE | Panther Fusion SARS-CoV-2 Assay (Hologic) RT-PCR testing | 19.900 | NA | NA |
| COVID-POS-018 | CVP-13 | COPAN | UTM | Validation Study | April | New York | POSITIVE | Panther Fusion SARS-CoV-2 Assay (Hologic) RT-PCR testing | Undetermined | NA | NA |
| COVID-POS-019 | CVP-14 | BD | UTM | Validation Study | April | New York | POSITIVE | Panther Fusion SARS-CoV-2 Assay (Hologic) RT-PCR testing | Undetermined | NA | NA |
| COVID-POS-020 | CVP-15 | REMEL M6 | M6 | Validation Study | April | New York | POSITIVE | Panther Fusion SARS-CoV-2 Assay (Hologic) RT-PCR testing | 21.450 | NA | NA |
| COVID-POS-021 | CVP-16 | BD | UTM | Validation Study | April | New York | POSITIVE | Panther Fusion SARS-CoV-2 Assay (Hologic) RT-PCR testing | Undetermined | NA | NA |
| COVID-POS-022 | CVP-17 | BD | UTM | Validation Study | April | New York | POSITIVE | Panther Fusion SARS-CoV-2 Assay (Hologic) RT-PCR testing | 30.931 | NA | NA |
| COVID-POS-023 | CVP-18 | BD | UTM | Validation Study | April | New York | POSITIVE | Panther Fusion SARS-CoV-2 Assay (Hologic) RT-PCR testing | 32.486 | NA | NA |
| COVID-POS-024 | CVP-19 | BD | UTM | Validation Study | April | New York | POSITIVE | Panther Fusion SARS-CoV-2 Assay (Hologic) RT-PCR testing | 24.077 | NA | NA |
| COVID-POS-025 | CVP-20 | BD | UTM | Validation Study | April | New York | POSITIVE | Panther Fusion SARS-CoV-2 Assay (Hologic) RT-PCR testing | 32.812 | NA | NA |
| COVID-POS-026 | CVP-21 | BD | UTM | Validation Study | April | New York | POSITIVE | Panther Fusion SARS-CoV-2 Assay (Hologic) RT-PCR testing | 29.486 | NA | NA |
| COVID-POS-027 | CVP-22 | BD | UTM | Validation Study | April | New York | POSITIVE | Panther Fusion SARS-CoV-2 Assay (Hologic) RT-PCR testing | 20.357 | NA | NA |
| COVID-POS-028 | CVP-23 | NA | NA | Validation Study | April | New York | POSITIVE | Panther Fusion SARS-CoV-2 Assay (Hologic) RT-PCR testing | 22.050 | NA | NA |
| COVID-POS-029 | CVP-24 | COPAN | UTM | Validation Study | April | New York | POSITIVE | Panther Fusion SARS-CoV-2 Assay (Hologic) RT-PCR testing | 24.973 | NA | NA |
| COVID-POS-030 | CVP-25 | COPAN | UTM | Validation Study | April | New York | POSITIVE | Panther Fusion SARS-CoV-2 Assay (Hologic) RT-PCR testing | 22.371 | NA | NA |
| COVID-POS-031 | CVP-26 | BD | UTM | Validation Study | April | New York | POSITIVE | Panther Fusion SARS-CoV-2 Assay (Hologic) RT-PCR testing | 28.781 | NA | NA |
| COVID-POS-032 | CVP-27 | BD | UTM | Validation Study | April | New York | POSITIVE | Panther Fusion SARS-CoV-2 Assay (Hologic) RT-PCR testing | 33.635 | NA | NA |
| COVID-POS-033 | CVP-28 | COPAN | UTM | Validation Study | April | New York | POSITIVE | Panther Fusion SARS-CoV-2 Assay (Hologic) RT-PCR testing | 17.872 | NA | NA |
| COVID-POS-034 | CVP-29 | BD | UTM | Validation Study | April | New York | POSITIVE | Panther Fusion SARS-CoV-2 Assay (Hologic) RT-PCR testing | 33.757 | NA | NA |
| COVID-POS-035 | CVP-30 | COPAN | UTM | Validation Study | April | New York | POSITIVE | Panther Fusion SARS-CoV-2 Assay (Hologic) RT-PCR testing | 31.576 | NA | NA |
| COVID-NEG-032 | CVN-31 | Aptima MultiTest | STM | Independent Validation | July | New York | NEGATIVE | Panther Fusion SARS-CoV-2 Assay (Hologic) RT-PCR testing | Undetermined | NA | NA |
| COVID-NEG-033 | CVN-32 | Aptima MultiTest | STM | Independent Validation | July | New York | NEGATIVE | Panther Fusion SARS-CoV-2 Assay (Hologic) RT-PCR testing | Undetermined | NA | NA |
| COVID-NEG-034 | CVN-33 | Aptima MultiTest | STM | Independent Validation | July | New York | NEGATIVE | Panther Fusion SARS-CoV-2 Assay (Hologic) RT-PCR testing | Undetermined | NA | NA |
| COVID-NEG-035 | CVN-34 | Aptima MultiTest | STM | Independent Validation | July | New York | NEGATIVE | Panther Fusion SARS-CoV-2 Assay (Hologic) RT-PCR testing | Undetermined | NA | NA |
| COVID-NEG-036 | CVN-35 | Aptima MultiTest | STM | Independent Validation | July | New York | NEGATIVE | Panther Fusion SARS-CoV-2 Assay (Hologic) RT-PCR testing | Undetermined | NA | NA |
| COVID-NEG-037 | CVN-36 | Aptima MultiTest | STM | Independent Validation | July | New York | NEGATIVE | Panther Fusion SARS-CoV-2 Assay (Hologic) RT-PCR testing | Undetermined | NA | NA |
| COVID-NEG-038 | CVN-37 | Aptima MultiTest | STM | Independent Validation | July | New York | NEGATIVE | Panther Fusion SARS-CoV-2 Assay (Hologic) RT-PCR testing | Undetermined | NA | NA |
| COVID-NEG-039 | CVN-38 | Aptima MultiTest | STM | Independent Validation | July | New York | NEGATIVE | Panther Fusion SARS-CoV-2 Assay (Hologic) RT-PCR testing | Undetermined | NA | NA |
| COVID-NEG-040 | CVN-39 | Aptima MultiTest | STM | Independent Validation | July | New York | NEGATIVE | Panther Fusion SARS-CoV-2 Assay (Hologic) RT-PCR testing | Undetermined | NA | NA |
| COVID-NEG-041 | CVN-40 | Aptima MultiTest | STM | Independent Validation | July | New York | NEGATIVE | Panther Fusion SARS-CoV-2 Assay (Hologic) RT-PCR testing | Undetermined | NA | NA |
| COVID-NEG-042 | CVN-41 | Aptima MultiTest | STM | Independent Validation | July | New York | NEGATIVE | Panther Fusion SARS-CoV-2 Assay (Hologic) RT-PCR testing | Undetermined | NA | NA |
| COVID-NEG-043 | CVN-42 | Aptima MultiTest | STM | Independent Validation | July | New York | NEGATIVE | Panther Fusion SARS-CoV-2 Assay (Hologic) RT-PCR testing | Undetermined | NA | NA |
| COVID-NEG-044 | CVN-43 | Aptima MultiTest | STM | Independent Validation | July | New York | NEGATIVE | Panther Fusion SARS-CoV-2 Assay (Hologic) RT-PCR testing | Undetermined | NA | NA |
| COVID-NEG-045 | CVN-44 | Aptima MultiTest | STM | Independent Validation | July | New York | NEGATIVE | Panther Fusion SARS-CoV-2 Assay (Hologic) RT-PCR testing | Undetermined | NA | NA |
| COVID-NEG-046 | CVN-45 | Aptima MultiTest | STM | Independent Validation | July | New York | NEGATIVE | Panther Fusion SARS-CoV-2 Assay (Hologic) RT-PCR testing | Undetermined | NA | NA |
| COVID-NEG-047 | CVN-46 | Aptima MultiTest | STM | Independent Validation | July | New York | NEGATIVE | Panther Fusion SARS-CoV-2 Assay (Hologic) RT-PCR testing | Undetermined | NA | NA |
| COVID-NEG-048 | CVN-47 | Aptima MultiTest | STM | Independent Validation | July | New York | NEGATIVE | Panther Fusion SARS-CoV-2 Assay (Hologic) RT-PCR testing | Undetermined | NA | NA |
| COVID-NEG-049 | CVN-48 | Aptima MultiTest | STM | Independent Validation | July | New York | NEGATIVE | Panther Fusion SARS-CoV-2 Assay (Hologic) RT-PCR testing | Undetermined | NA | NA |
| COVID-NEG-050 | CVN-49 | Aptima MultiTest | STM | Independent Validation | July | New York | NEGATIVE | Panther Fusion SARS-CoV-2 Assay (Hologic) RT-PCR testing | Undetermined | NA | NA |
| COVID-NEG-051 | CVN-50 | Aptima MultiTest | STM | Independent Validation | July | New York | NEGATIVE | Panther Fusion SARS-CoV-2 Assay (Hologic) RT-PCR testing | Undetermined | NA | NA |
| COVID-NEG-052 | CVN-51 | Aptima MultiTest | STM | Independent Validation | July | New York | NEGATIVE | Panther Fusion SARS-CoV-2 Assay (Hologic) RT-PCR testing | Undetermined | NA | NA |
| COVID-NEG-053 | CVN-52 | Aptima MultiTest | STM | Independent Validation | July | New York | NEGATIVE | Panther Fusion SARS-CoV-2 Assay (Hologic) RT-PCR testing | Undetermined | NA | NA |
| COVID-NEG-054 | CVN-53 | Aptima MultiTest | STM | Independent Validation | July | New York | NEGATIVE | Panther Fusion SARS-CoV-2 Assay (Hologic) RT-PCR testing | Undetermined | NA | NA |
| COVID-NEG-055 | CVN-54 | Aptima MultiTest | STM | Independent Validation | July | New York | NEGATIVE | Panther Fusion SARS-CoV-2 Assay (Hologic) RT-PCR testing | Undetermined | NA | NA |
| COVID-NEG-056 | CVN-55 | Aptima MultiTest | STM | Independent Validation | July | New York | NEGATIVE | Panther Fusion SARS-CoV-2 Assay (Hologic) RT-PCR testing | Undetermined | NA | NA |
| COVID-NEG-057 | CVN-56 | Aptima MultiTest | STM | Independent Validation | July | New York | NEGATIVE | Panther Fusion SARS-CoV-2 Assay (Hologic) RT-PCR testing | Undetermined | NA | NA |
| COVID-NEG-058 | CVN-57 | Aptima MultiTest | STM | Independent Validation | July | New York | NEGATIVE | Panther Fusion SARS-CoV-2 Assay (Hologic) RT-PCR testing | Undetermined | NA | NA |
| COVID-NEG-059 | CVN-58 | Aptima MultiTest | STM | Independent Validation | July | New York | NEGATIVE | Panther Fusion SARS-CoV-2 Assay (Hologic) RT-PCR testing | Undetermined | NA | NA |
| COVID-NEG-060 | CVN-59 | Aptima MultiTest | STM | Independent Validation | July | New York | NEGATIVE | Panther Fusion SARS-CoV-2 Assay (Hologic) RT-PCR testing | Undetermined | NA | NA |
| COVID-NEG-061 | CVN-60 | Aptima MultiTest | STM | Independent Validation | July | New York | NEGATIVE | Panther Fusion SARS-CoV-2 Assay (Hologic) RT-PCR testing | Undetermined | NA | NA |
| COVID-POS-037 | CVP-31 | Aptima MultiTest | STM | Independent Validation | July | New York | POSITIVE | Panther Fusion SARS-CoV-2 Assay (Hologic) RT-PCR testing | Undetermined | NA | NA |
| COVID-POS-038 | CVP-32 | Aptima MultiTest | STM | Independent Validation | July | New York | POSITIVE | Panther Fusion SARS-CoV-2 Assay (Hologic) RT-PCR testing | 18.761 | NA | NA |
| COVID-POS-039 | CVP-33 | Aptima MultiTest | STM | Independent Validation | July | New York | POSITIVE | Panther Fusion SARS-CoV-2 Assay (Hologic) RT-PCR testing | 29.045 | NA | NA |
| COVID-POS-040 | CVP-34 | Aptima MultiTest | STM | Independent Validation | July | New York | POSITIVE | Panther Fusion SARS-CoV-2 Assay (Hologic) RT-PCR testing | 23.459 | NA | NA |
| COVID-POS-041 | CVP-35 | Aptima MultiTest | STM | Independent Validation | July | New York | POSITIVE | Panther Fusion SARS-CoV-2 Assay (Hologic) RT-PCR testing | 27.451 | NA | NA |
| COVID-POS-042 | CVP-36 | Aptima MultiTest | STM | Independent Validation | July | New York | POSITIVE | Panther Fusion SARS-CoV-2 Assay (Hologic) RT-PCR testing | 30.180 | NA | NA |
| COVID-POS-043 | CVP-37 | Aptima MultiTest | STM | Independent Validation | July | New York | POSITIVE | Panther Fusion SARS-CoV-2 Assay (Hologic) RT-PCR testing | 24.750 | NA | NA |
| COVID-POS-044 | CVP-38 | Aptima MultiTest | STM | Independent Validation | July | New York | POSITIVE | Panther Fusion SARS-CoV-2 Assay (Hologic) RT-PCR testing | 27.416 | NA | NA |
| COVID-POS-045 | CVP-39 | Aptima MultiTest | STM | Independent Validation | July | New York | POSITIVE | Panther Fusion SARS-CoV-2 Assay (Hologic) RT-PCR testing | Undetermined | NA | NA |
| COVID-POS-046 | CVP-40 | Aptima MultiTest | STM | Independent Validation | July | New York | POSITIVE | Panther Fusion SARS-CoV-2 Assay (Hologic) RT-PCR testing | 28.000 | NA | NA |
| COVID-POS-047 | CVP-41 | Aptima MultiTest | STM | Independent Validation | July | New York | POSITIVE | Panther Fusion SARS-CoV-2 Assay (Hologic) RT-PCR testing | Undetermined | NA | NA |
| COVID-POS-048 | CVP-42 | Aptima MultiTest | STM | Independent Validation | July | New York | POSITIVE | Panther Fusion SARS-CoV-2 Assay (Hologic) RT-PCR testing | 17.190 | NA | NA |
| COVID-POS-049 | CVP-43 | Aptima MultiTest | STM | Independent Validation | July | New York | POSITIVE | Panther Fusion SARS-CoV-2 Assay (Hologic) RT-PCR testing | 15.798 | NA | NA |
| COVID-POS-050 | CVP-44 | Aptima MultiTest | STM | Independent Validation | July | New York | POSITIVE | Panther Fusion SARS-CoV-2 Assay (Hologic) RT-PCR testing | Undetermined | NA | NA |
| COVID-POS-051 | CVP-45 | Aptima MultiTest | STM | Independent Validation | July | New York | POSITIVE | Panther Fusion SARS-CoV-2 Assay (Hologic) RT-PCR testing | Undetermined | NA | NA |

| | | | | | | | | | | | |
|---|---|---|---|---|---|---|---|---|---|---|---|
| COVID-POS-052 | CVP-46 | Aptima MultiTest | STM | Independent Validation | July | New York | POSITIVE | Panther Fusion SARS-CoV-2 Assay (Hologic) RT-PCR testing | Undetermined | NA | NA |
| COVID-POS-053 | CVP-47 | Aptima MultiTest | STM | Independent Validation | July | New York | POSITIVE | Panther Fusion SARS-CoV-2 Assay (Hologic) RT-PCR testing | Undetermined | NA | NA |
| COVID-POS-054 | CVP-48 | Aptima MultiTest | STM | Independent Validation | July | New York | POSITIVE | Panther Fusion SARS-CoV-2 Assay (Hologic) RT-PCR testing | 29.809 | NA | NA |
| COVID-POS-055 | CVP-49 | Aptima MultiTest | STM | Independent Validation | July | New York | POSITIVE | Panther Fusion SARS-CoV-2 Assay (Hologic) RT-PCR testing | 22.164 | NA | NA |
| COVID-POS-056 | CVP-50 | Aptima MultiTest | STM | Independent Validation | July | New York | POSITIVE | Panther Fusion SARS-CoV-2 Assay (Hologic) RT-PCR testing | 26.916 | NA | NA |
| COVID-POS-057 | CVP-51 | Aptima MultiTest | STM | Independent Validation | July | New York | POSITIVE | Panther Fusion SARS-CoV-2 Assay (Hologic) RT-PCR testing | 29.363 | NA | NA |
| COVID-POS-058 | CVP-52 | Aptima MultiTest | STM | Independent Validation | July | New York | POSITIVE | Panther Fusion SARS-CoV-2 Assay (Hologic) RT-PCR testing | 28.931 | NA | NA |
| COVID-POS-059 | CVP-53 | Aptima MultiTest | STM | Independent Validation | July | New York | POSITIVE | Panther Fusion SARS-CoV-2 Assay (Hologic) RT-PCR testing | 35.437 | NA | NA |
| COVID-POS-060 | CVP-54 | Aptima MultiTest | STM | Independent Validation | July | New York | POSITIVE | Panther Fusion SARS-CoV-2 Assay (Hologic) RT-PCR testing | Undetermined | NA | NA |
| COVID-POS-061 | CVP-55 | Aptima MultiTest | STM | Independent Validation | July | New York | POSITIVE | Panther Fusion SARS-CoV-2 Assay (Hologic) RT-PCR testing | 25.584 | NA | NA |
| COVID-POS-062 | CVP-56 | Aptima MultiTest | STM | Independent Validation | July | New York | POSITIVE | Panther Fusion SARS-CoV-2 Assay (Hologic) RT-PCR testing | Undetermined | NA | NA |
| COVID-POS-063 | CVP-57 | Aptima MultiTest | STM | Independent Validation | July | New York | POSITIVE | Panther Fusion SARS-CoV-2 Assay (Hologic) RT-PCR testing | Undetermined | NA | NA |
| COVID-POS-064 | CVP-58 | Aptima MultiTest | STM | Independent Validation | July | New York | POSITIVE | Panther Fusion SARS-CoV-2 Assay (Hologic) RT-PCR testing | 22.904 | NA | NA |
| COVID-POS-065 | CVP-59 | Aptima MultiTest | STM | Independent Validation | July | New York | POSITIVE | Panther Fusion SARS-CoV-2 Assay (Hologic) RT-PCR testing | 22.326 | NA | NA |
| COVID-POS-066 | CVP-60 | Aptima MultiTest | STM | Independent Validation | July | New York | POSITIVE | Panther Fusion SARS-CoV-2 Assay (Hologic) RT-PCR testing | 27.392 | NA | NA |
| COVID-Pos-067_S8_ | CVP-61 | NA | NA | Geographic Validation | NA | Tennessee | POSITIVE | Roche Cobas SARS-CoV-2 Assay RT-qPCR testing | NA | 19.2 | 19.82 |
| COVID-Pos-068_S9_ | CVP-62 | NA | NA | Geographic Validation | NA | Tennessee | POSITIVE | Roche Cobas SARS-CoV-2 Assay RT-qPCR testing | NA | 22.14 | 22.78 |
| COVID-Pos-069_S10 | CVP-63 | NA | NA | Geographic Validation | NA | Tennessee | POSITIVE | Roche Cobas SARS-CoV-2 Assay RT-qPCR testing | NA | 31.4 | 33.52 |
| NA | CVP-64 | NA | NA | Geographic Validation | NA | Tennessee | POSITIVE | Roche Cobas SARS-CoV-2 Assay RT-qPCR testing | NA | 24.51 | 25.36 |
| COVID-Pos-071_S11 | CVP-65 | NA | NA | Geographic Validation | NA | Tennessee | POSITIVE | Roche Cobas SARS-CoV-2 Assay RT-qPCR testing | NA | 33.43 | 36.7 |
| COVID-Pos-072_S12 | CVP-66 | NA | NA | Geographic Validation | NA | Tennessee | POSITIVE | Roche Cobas SARS-CoV-2 Assay RT-qPCR testing | NA | 20.7 | 21.54 |
| COVID-Pos-073_S13 | CVP-67 | NA | NA | Geographic Validation | NA | Tennessee | POSITIVE | Roche Cobas SARS-CoV-2 Assay RT-qPCR testing | NA | 25.32 | 26.43 |
| COVID-Pos-074_S14 | CVP-68 | NA | NA | Geographic Validation | NA | Tennessee | POSITIVE | Roche Cobas SARS-CoV-2 Assay RT-qPCR testing | NA | 20.76 | 21.29 |
| NA | CVP-69 | NA | NA | Geographic Validation | NA | Tennessee | POSITIVE | Roche Cobas SARS-CoV-2 Assay RT-qPCR testing | NA | 18.31 | 19.02 |
| COVID-Pos-076_S15 | CVP-70 | NA | NA | Geographic Validation | NA | Tennessee | POSITIVE | Roche Cobas SARS-CoV-2 Assay RT-qPCR testing | NA | 31.2 | 33.16 |
| COVID-Pos-077_S16 | CVP-71 | NA | NA | Geographic Validation | NA | Tennessee | POSITIVE | Roche Cobas SARS-CoV-2 Assay RT-qPCR testing | NA | 30.9 | 33.2 |
| COVID-Pos-078_S17 | CVP-72 | NA | NA | Geographic Validation | NA | Tennessee | POSITIVE | Roche Cobas SARS-CoV-2 Assay RT-qPCR testing | NA | 32.21 | 34.5 |
| COVID-Pos-079_S18 | CVP-73 | NA | NA | Geographic Validation | NA | Tennessee | POSITIVE | Roche Cobas SARS-CoV-2 Assay RT-qPCR testing | NA | 19.81 | 20.61 |
| COVID-Pos-080_S19 | CVP-74 | NA | NA | Geographic Validation | NA | Tennessee | POSITIVE | Roche Cobas SARS-CoV-2 Assay RT-qPCR testing | NA | 29.02 | 30.66 |
| COVID-Pos-081_S20 | CVP-75 | NA | NA | Geographic Validation | NA | Tennessee | POSITIVE | Roche Cobas SARS-CoV-2 Assay RT-qPCR testing | NA | 18 | 18.63 |
| NA | CVP-76 | NA | NA | Geographic Validation | NA | Tennessee | POSITIVE | Roche Cobas SARS-CoV-2 Assay RT-qPCR testing | NA | 17.31 | 17.8 |
| COVID-Pos-083_S21 | CVP-77 | NA | NA | Geographic Validation | NA | Tennessee | POSITIVE | Roche Cobas SARS-CoV-2 Assay RT-qPCR testing | NA | 30.86 | 33.33 |
| COVID-Pos-084_S22 | CVP-78 | NA | NA | Geographic Validation | NA | Tennessee | POSITIVE | Roche Cobas SARS-CoV-2 Assay RT-qPCR testing | NA | 29.83 | 31.21 |
| COVID-Pos-085_S23 | CVP-79 | NA | NA | Geographic Validation | NA | Tennessee | POSITIVE | Roche Cobas SARS-CoV-2 Assay RT-qPCR testing | NA | 19.52 | 20.19 |
| COVID-Pos-086_S7_ | CVP-80 | NA | NA | Geographic Validation | NA | Tennessee | POSITIVE | Roche Cobas SARS-CoV-2 Assay RT-qPCR testing | NA | 31.46 | 34.13 |

**Supplemental Table 4.** The preliminary LoD was established by testing 10-fold dilutions of SARS-CoV-2 synthetic RNA using two different synthetic controls in duplicates. The preliminary LoD was confirmed by testing triplicates of 2-fold dilutions (2560 copies/ml, 1280 copies/ml, 640 copies/ml, 320 copies/ml, 160 copies/ml, 80 copies/ml, and 40 copies/ml) by spiking the quantified heat-inactivated SARS-CoV-2 into negative respiratory clinical matrices. The LOD was determined to be 800 copies/ml. The LOD (800 copies/ml) was replicated 30 times. 29/30 (96.67%) samples were positive.

Presence: if SARS-CoV-2 was detected. We devised a metric [I1] to compute the integral under a curve created by calculating the coverage at 1X depth using a sliding window scheme (with a window size of 100 and step size of 10). This metric was then log transformed. We have set a threshold of 8.6 if samples had less than 10,000 bases on target or 9.5 if samples had more than 10,000 bases on target. Metrics were calculated using the R statistical software package (v4.0.1). X1-X500: percent coverage at 1X-500X depths, used internally to inform the analysis. E1: evenness, used internally to inform the analysis. I1: integral. The following HS metrics are reported: bait territory, percent of selected bases, target territory, genome size, total reads and on target bases. Red indicates deviant results. Additionally, the number of reads mapped to the human genome will provide an internal control in each individual sample to be called valid.

| Specimen_ID | Copy_number | presence | clade | reads | X1 | X5 | X10 | X50 | X100 | X500 | E1 | I1 | validity | BAIT_TERRITO | PCT_SELECTED | TARGET_TERRI | GENOME_SIZE | TOTAL_READS | HUMAN_ ON_TARGET_BASES |
|---|---|---|---|---|---|---|---|---|---|---|---|---|---|---|---|---|---|---|---|
| Ctrl1-1_S8_L001_R1_00 | 1,000,000 copi | TRUE | 19A | 29242555 | 0.999832608 | 0.999832608 | 0.999832608 | 0.999832608 | 0.999832608 | 0.49390693 | 0.985538337 | 10.3118109 | VALID | 29869 | 0.961489 | 29870 | 3259356417 | 29196007 | 14388940 |
| Ctrl1-1e1-1_S24_L001_ | 10 copies | FALSE | NOT_IDENTIFIE | 10419736 | 0.076163375 | 0.01469702 | 0.008938735 | 0 | 0 | 0 | 0 | 7.739785667 | VALID | 29869 | 6.00E-06 | 29870 | 3259356417 | 10386287 | 121167542 |
| Ctrl1-1e1-2_S31_L001_ | 10 copies | FALSE | NOT_IDENTIFIE | 5404930 | 0.253398058 | 0.114094409 | 0.052058922 | 0 | 0 | 0 | 0.153766321 | 8.941866573 | VALID | 29869 | 6.40E-05 | 29870 | 3259356417 | 5388306 | 39509304 |
| Ctrl1-1e2-1_S23_L001_ | 100 copies | TRUE | 19A | 12682484 | 0.883662538 | 0.716538333 | 0.487345162 | 0.002142618 | 0 | 0 | 0.609015735 | 10.18794516 | VALID | 29869 | 0.000215 | 29870 | 3259356417 | 12638565 | 137696159 |
| Ctrl1-1e2-2_S30_L001_ | 100 copies | TRUE | 19A | 7914059 | 0.628054905 | 0.59306997 | 0.532909274 | 0.140308001 | 0.025677938 | 0 | 0.492902578 | 9.846468767 | VALID | 29869 | 0.000613 | 29870 | 3259356417 | 7894599 | 55935164 |
| Ctrl1-1e3-1_S22_L001_ | 1000 copies | TRUE | 19A | 9361244 | 0.999029126 | 0.994844325 | 0.97268162 | 0.525477067 | 0.07248075 | 0 | 0.783089615 | 10.3110048 | VALID | 29869 | 0.001467 | 29870 | 3259356417 | 9329773 | 193556872 |
| Ctrl1-1e3-2_S29_L001_ | 1000 copies | TRUE | 19A | 8974574 | 0.999497824 | 0.999296953 | 0.997288249 | 0.927552728 | 0.631235353 | 0 | 0.800771058 | 10.31147511 | VALID | 29869 | 0.003328 | 29870 | 3259356417 | 8953657 | 107550804 |
| Ctrl1-1e4-1_S21_L001_ | 10,000 copies | TRUE | 19A | 11424774 | 0.999698694 | 0.999698694 | 0.999531302 | 0.998091731 | 0.992835621 | 0.018312688 | 0.952392198 | 10.3116766 | VALID | 29869 | 0.016043 | 29870 | 3259356417 | 11334486 | 250479871 |
| Ctrl1-1e4-2_S28_L001_ | 10,000 copies | TRUE | 19A | 10661256 | 0.999832608 | 0.999832608 | 0.999832608 | 0.999296953 | 0.997957817 | 0.027150988 | 0.97135418 | 10.3118109 | VALID | 29869 | 0.026395 | 29870 | 3259356417 | 10629908 | 116229177 |
| Ctrl1-1e5-1_S20_L001_ | 100,000 copies | TRUE | 19A | 8260389 | 0.999832608 | 0.999832608 | 0.999832608 | 0.999832608 | 0.999263475 | 0.044994978 | 0.98540159 | 10.3118109 | VALID | 29869 | 0.126148 | 29870 | 3259356417 | 8233645 | 140795110 |
| Ctrl1-1e5-2_S27_L001_ | 100,000 copies | TRUE | 19A | 13357543 | 0.999832608 | 0.999832608 | 0.999832608 | 0.999832608 | 0.999665216 | 0.072079009 | 0.987786576 | 10.3118109 | VALID | 29869 | 0.262216 | 29870 | 3259356417 | 13320088 | 113717738 |
| Ctrl1-1e6-1_S19_L001_ | 1,000,000 copi | TRUE | 19A | 20139533 | 0.999832608 | 0.999832608 | 0.999832608 | 0.999832608 | 0.999665216 | 0.190525611 | 0.988589305 | 10.3118109 | VALID | 29869 | 0.630525 | 29870 | 3259356417 | 20099108 | 134874935 |
| Ctrl1-1e6-2_S26_L001_ | 1,000,000 copi | TRUE | 19A | 21207543 | 0.999832608 | 0.999832608 | 0.999832608 | 0.999832608 | 0.999665216 | 0.161031135 | 0.989106499 | 10.3118109 | VALID | 29869 | 0.513411 | 29870 | 3259356417 | 21161007 | 157192338 |
| Ctrl1-2_S17_L001_R1_0 | 1,000,000 copi | TRUE | 19A | 36430747 | 0.999832608 | 0.999832608 | 0.999832608 | 0.999832608 | 0.999665216 | 0.479879478 | 0.986268274 | 10.3118109 | VALID | 29869 | 0.869207 | 29870 | 3259356417 | 36349956 | 63869908 |
| Ctrl2-1e1-1_S38_L001_ | 10 copies | FALSE | NOT_IDENTIFIE | 10061630 | 0.130632742 | 0.083495146 | 0.046534985 | 0 | 0 | 0 | 0.09916304 | 8.279294857 | VALID | 29869 | 2.40E-05 | 29870 | 3259356417 | 10034833 | 54260476 |
| Ctrl2-1e1-2_S45_L001_ | 10 copies | FALSE | NOT_IDENTIFIE | 10320262 | 0.065048544 | 0.016839638 | 0 | 0 | 0 | 0 | 0 | 7.582038785 | VALID | 29869 | 5.00E-06 | 29870 | 3259356417 | 10295989 | 136974435 |
| Ctrl2-1e2-1_S37_L001_ | 100 copies | TRUE | 19A | 10734891 | 0.598627385 | 0.390927352 | 0.199899565 | 0 | 0 | 0 | 0.416002678 | 9.800402784 | VALID | 29869 | 0.000105 | 29870 | 3259356417 | 10704358 | 67888592 |
| Ctrl2-1e2-2_S44_L001_ | 100 copies | TRUE | 19A | 14469329 | 0.689019083 | 0.277770338 | 0.051456311 | 0 | 0 | 0 | 0.401606964 | 9.94188531 | VALID | 29869 | 4.70E-05 | 29870 | 3259356417 | 14434586 | 203914099 |
| Ctrl2-1e3-1_S36_L001_ | 1000 copies | TRUE | 19A | 10553630 | 0.985838634 | 0.922765316 | 0.757415467 | 0.027150988 | 0 | 0 | 0.715049528 | 10.29798401 | VALID | 29869 | 0.000415 | 29870 | 3259356417 | 10522237 | 64406976 |
| Ctrl2-1e3-2_S43_L001_ | 1000 copies | TRUE | 19A | 15517306 | 0.999096083 | 0.98844995 | 0.949079344 | 0.168463341 | 0.006427854 | 0 | 0.777965809 | 10.31128365 | VALID | 29869 | 0.000484 | 29870 | 3259356417 | 15476960 | 196558644 |
| Ctrl2-1e4-1_S35_L001_ | 10,000 copies | TRUE | 19A | 9463750 | 1 | 1 | 1 | 0.99916304 | 0.996149983 | 0.021861399 | 0.964455602 | 10.31197876 | VALID | 29869 | 0.022316 | 29870 | 3259356417 | 9439784 | 84515603 |
| Ctrl2-1e4-2_S42_L001_ | 10,000 copies | TRUE | 19A | 10707048 | 1 | 1 | 0.999866086 | 0.984198192 | 0.798292601 | 0 | 0.839881617 | 10.31197876 | VALID | 29869 | 0.003186 | 29870 | 3259356417 | 10676412 | 64493503 |
| Ctrl2-1e5-1_S34_L001_ | 100,000 copies | TRUE | 19A | 6097557 | 1 | 1 | 1 | 1 | 0.999933043 | 0.051623703 | 0.987300292 | 10.31197876 | VALID | 29869 | 0.313331 | 29870 | 3259356417 | 6083804 | 47758321 |
| Ctrl2-1e5-2_S41_L001_ | 100,000 copies | TRUE | 19A | 11434148 | 1 | 1 | 1 | 1 | 0.999899565 | 0.035888852 | 0.982747744 | 10.31197876 | VALID | 29869 | 0.045606 | 29870 | 3259356417 | 11401573 | 64417387 |
| Ctrl2-1e6-1_S33_L001_ | 1,000,000 copi | TRUE | 19A | 35898577 | 1 | 1 | 1 | 1 | 1 | 0.305122196 | 0.987777456 | 10.31197876 | VALID | 29869 | 0.768918 | 29870 | 3259356417 | 35829039 | 142123437 |
| Ctrl2-1e6-2_S40_L001_ | 1,000,000 copi | TRUE | 19A | 13566990 | 1 | 1 | 1 | 1 | 1 | 0.101606964 | 0.988473079 | 10.31197876 | VALID | 29869 | 0.411275 | 29870 | 3259356417 | 13537196 | 64086847 |
| HuRef-1_S9_L001_R1_0 | IC-1: Negative i | FALSE | NOT_IDENTIFIE | 924528 | 0.109708738 | 0.049849347 | 0.005925678 | 0 | 0 | 0 | 0 | 8.097874995 | VALID | 29869 | 0.000123 | 29870 | 3259356417 | 921945 | 4243363 |
| HuRef-2_S18_L001_R1_ | IC-2: Negative i | FALSE | NOT_IDENTIFIE | 2889969 | 0.119584868 | 0.020689655 | 0 | 0 | 0 | 0 | 0 | 8.190931278 | VALID | 29869 | 2.70E-05 | 29870 | 3259356417 | 2880457 | 18141077 |
| NC-1_S25_L001_R1_001 | NTC-1: Negativ | FALSE | NOT_IDENTIFIE | 10776962 | 0.032373619 | 0 | 0 | 0 | 0 | 0 | 0 | 6.884248831 | VALID | 29869 | 1.00E-06 | 29870 | 3259356417 | 10749411 | 160928072 |
| NC-2_S32_L001_R1_001 | NTC-2: Negativ | FALSE | NOT_IDENTIFIE | 2847967 | 0.004385671 | 0 | 0 | 0 | 0 | 0 | 0 | 4.885247659 | VALID | 29869 | 0 | 29870 | 3259356417 | 2839939 | 23969249 |
| NC-3_S39_L001_R1_001 | NTC-3: Negativ | FALSE | NOT_IDENTIFIE | 11229155 | 0.031904921 | 0 | 0 | 0 | 0 | 0 | 0 | 6.86966524 | VALID | 29869 | 1.00E-06 | 29870 | 3259356417 | 11198299 | 68087022 |
| NC-4_S46_L001_R1_001 | NTC-4: Negativ | FALSE | NOT_IDENTIFIE | 17662430 | 0.004318714 | 0 | 0 | 0 | 0 | 0 | 0 | 4.86986274 | VALID | 29869 | 0 | 29870 | 3259356417 | 17608283 | 255817281 |
| Negative-Control-1_S7_ | NTC-5: Negativ | FALSE | NOT_IDENTIFIE | 59151 | 0.151791095 | 0.036424506 | 0.033478406 | 0.007833947 | 0 | 0 | 0.041613659 | 8.429410167 | VALID | 29869 | 0.012513 | 29870 | 3259356417 | 58914 | 652 |
| Negative-Control-2_S16 | NTC-6: Negativ | FALSE | NOT_IDENTIFIE | 3722 | 0.014830934 | 0.014830934 | 0.014830934 | 0.002644794 | 0 | 0 | 0 | 6.103620106 | VALID | 29869 | 0.384921 | 29870 | 3259356417 | 392 | 10607 |

| Specimen_ID | Copy_number | presence | clade | reads | X1 | X5 | X10 | X50 | X100 | X500 | E1 | I1 | validity | BAIT_TERRITO | PCT_SELECTED | TARGET_TERRI | GENOME_SIZE | TOTAL_READS | HUMAN_ ON_TARGET_BASES |
|---|---|---|---|---|---|---|---|---|---|---|---|---|---|---|---|---|---|---|---|
| 10-copies-1_S27_L001_ | 40_copies_per | TRUE | other | 3635714 | 0.999096083 | 0.991965182 | 0.920053565 | 0.447639772 | 0.112688316 | 0 | 0.68126849 | 10.31182765 | VALID | 29869 | 0.003504 | 29870 | 3259356417 | 3530179 | 192878728 |
| 10-copies-2_S28_L001_ | 40_copies_per | TRUE | 19A | 5210775 | 0.500903917 | 0.478908604 | 0.4579846 | 0.199397389 | 0.07037161 | 0 | 0.422484613 | 9.620548423 | VALID | 29869 | 0.001211 | 29870 | 3259356417 | 5204956 | 306570686 |
| 10-copies-3_S29_L001_ | 40_copies_per | TRUE | 19B | 3228494 | 0.976297288 | 0.707532641 | 0.40629394 | 0.000334784 | 0 | 0 | 0.615203381 | 10.28872586 | VALID | 29869 | 0.000687 | 29870 | 3259356417 | 3226232 | 301819726 |
| 20-copies-1_S30_L001_ | 80_copies_per | TRUE | 19B | 1558234 | 0.78339471 | 0.60629394 | 0.548878473 | 0.218212253 | 0.053632407 | 0 | 0.491863507 | 10.06977817 | VALID | 29869 | 0.004621 | 29870 | 3259356417 | 1522751 | 69894943 |
| 20-copies-2_S31_L001_ | 80_copies_per | TRUE | 19A | 1697104 | 0.590023435 | 0.569199866 | 0.503481754 | 0.119450954 | 0.01238701 | 0 | 0.476665551 | 9.787067224 | VALID | 29869 | 0.002621 | 29870 | 3259356417 | 1694199 | 94566358 |
| 20-copies-3_S32_L001_ | 80_copies_per | TRUE | 19A | 2871430 | 0.726247071 | 0.53150318 | 0.307499163 | 0.003816538 | 0 | 0 | 0.502025444 | 9.994795243 | VALID | 29869 | 0.000574 | 29870 | 3259356417 | 2869068 | 241551634 |
| 40-copies-1_S49_L001_ | 160_copies_pe | TRUE | other | 1781187 | 0.568831604 | 0.517140944 | 0.404954804 | 0.048342819 | 0.006226984 | 0 | 0.444584868 | 9.748881316 | VALID | 29869 | 0.001835 | 29870 | 3259356417 | 1707976 | 80694315 |
| 40-copies-2_S50_L001_ | 160_copies_pe | FALSE | NOT_IDENTIFIE | 5101840 | 0.009273519 | 0.009273519 | 0.004318714 | 0 | 0 | 0 | 0 | 5.634067842 | VALID | 29869 | 7.00E-06 | 29870 | 3259356417 | 5041740 | 933784 |
| 40-copies-3_S51_L001_ | 160_copies_pe | TRUE | 19B | 4084246 | 0.970539002 | 0.964847673 | 0.962202879 | 0.928456645 | 0.84137931 | 0.023568798 | 0.770097961 | 10.28380192 | VALID | 29869 | 0.016891 | 29870 | 3259356417 | 4079331 | 316512116 |
| 80-copies-1_S52_L001_ | 320_copies_pe | TRUE | 19A | 3388994 | 0.745329762 | 0.285771677 | 0.06722464 | 0 | 0 | 0 | 0.41575159 | 10.01908689 | VALID | 29869 | 0.00023 | 29870 | 3259356417 | 3305524 | 239416545 |
| 80-copies-2_S53_L001_ | 320_copies_pe | FALSE | NOT_IDENTIFIE | 7571989 | 0.01975226 | 0.008938735 | 0 | 0 | 0 | 0 | 0 | 6.390172873 | VALID | 29869 | 3.00E-06 | 29870 | 3259356417 | 6925864 | 2255704 |
| 80-copies-3_S54_L001_ | 320_copies_pe | TRUE | 19B | 4468662 | 0.993538668 | 0.993170405 | 0.985135588 | 0.908369602 | 0.733578842 | 0.000234349 | 0.774570701 | 10.3066229 | VALID | 29869 | 0.008243 | 29870 | 3259356417 | 4463267 | 320677316 |
| 160-copies-1_S55_L001 | 640_copies_pe | TRUE | 19B | 2126034 | 0.914060931 | 0.907432206 | 0.863475059 | 0.419450954 | 0.097991296 | 0 | 0.678340896 | 10.22311388 | VALID | 29869 | 0.005902 | 29870 | 3259356417 | 2030354 | 80080662 |
| 160-copies-2_S56_L001 | 640_copies_pe | FALSE | NOT_IDENTIFIE | 11356658 | 0.032842317 | 0.032842317 | 0.032842317 | 0 | 0 | 0 | 0.032842317 | 6.898622795 | VALID | 29869 | 1.90E-05 | 29870 | 3259356417 | 11175869 | 1930825 |
| 160-copies-3_S57_L001 | 640_copies_pe | TRUE | 19B | 6818268 | 0.999029126 | 0.999029126 | 0.998995648 | 0.996083026 | 0.9840308 | 0.009106127 | 0.855548912 | 10.31181422 | VALID | 29869 | 0.011152 | 29870 | 3259356417 | 6810495 | 515686369 |
| 320-copies-1_S58_L001 | 1280_copies_p | TRUE | 19B | 1462814 | 0.988550385 | 0.98530298 | 0.97268162 | 0.653799799 | 0.268396384 | 0 | 0.721695295 | 10.30223378 | VALID | 29869 | 0.013312 | 29870 | 3259356417 | 1420671 | 77021657 |
| 320-copies-2_S59_L001 | 1280_copies_p | TRUE | 19B | 1988161 | 0.999229997 | 0.999196518 | 0.999196518 | 0.998727821 | 0.997589555 | 0.025644459 | 0.920015595 | 10.31184783 | VALID | 29869 | 0.073709 | 29870 | 3259356417 | 1957593 | 86974382 |
| 320-copies-3_S60_L001 | 1280_copies_p | TRUE | 19B | 3951638 | 0.997857382 | 0.997857382 | 0.997857382 | 0.968831604 | 0.884332106 | 0.00820221 | 0.795475555 | 10.31051747 | VALID | 29869 | 0.01646 | 29870 | 3259356417 | 3948697 | 242485573 |
| 640-copies-1_S61_L001 | 2560_copies_p | TRUE | 19A | 3037201 | 0.959223301 | 0.639805825 | 0.222865752 | 0 | 0 | 0 | 0.639825912 | 10.27168115 | VALID | 29869 | 0.000502 | 29870 | 3259356417 | 2988926 | 249004313 |
| 640-copies-2_S62_L001 | 2560_copies_p | TRUE | 19B | 9968231 | 0.99936391 | 0.99936391 | 0.99936391 | 0.999196518 | 0.998292601 | 0.01365919 | 0.932017419 | 10.31188476 | VALID | 29869 | 0.012978 | 29870 | 3259356417 | 9593974 | 496932225 |
| 640-copies-3_S63_L001 | 2560_copies_p | TRUE | 19B | 5407242 | 0.999464345 | 0.999029126 | 0.996752595 | 0.514462672 | 0.036826247 | 0 | 0.830057171 | 10.31190491 | VALID | 29869 | 0.002273 | 29870 | 3259356417 | 5403938 | 339433386 |
| COVID-Neg-003_S39_L0 | CLINICAL_MAT | FALSE | NOT_IDENTIFIE | 4133163 | 0 | 0 | 0 | 0 | 0 | 0 | 0 | #NAME? | VALID | 29869 | 0 | 29870 | 3259356417 | 4050110 | 315197180 |
| COVID-Neg-006_S40_L0 | CLINICAL_MAT | FALSE | NOT_IDENTIFIE | 7347273 | 0.023635755 | 0.004017409 | 0 | 0 | 0 | 0 | 0 | 6.569665573 | VALID | 29869 | 2.00E-06 | 29870 | 3259356417 | 7103177 | 459058602 |
| COVID-Neg-008_S41_L0 | CLINICAL_MAT | FALSE | NOT_IDENTIFIE | 6041431 | 0.001774356 | 0.001774356 | 0.001774356 | 0 | 0 | 0 | 0 | 3.980342249 | VALID | 29869 | 2.00E-06 | 29870 | 3259356417 | 5920322 | 1564807 |
| COVID-Neg-009_S42_L0 | CLINICAL_MAT | FALSE | NOT_IDENTIFIE | 5030196 | 0 | 0 | 0 | 0 | 0 | 0 | 0 | #NAME? | VALID | 29869 | 0 | 29870 | 3259356417 | 5019325 | 292523032 |
| COVID-Neg-014_S43_L0 | CLINICAL_MAT | FALSE | NOT_IDENTIFIE | 1613606 | 0.020087044 | 0.01975226 | 0.018613994 | 0.004184801 | 0 | 0 | 0.020087044 | 6.406979991 | VALID | 29869 | 8.10E-05 | 29870 | 3259356417 | 1609830 | 122253956 |
| Negative-Control-1_S25 | NTC-1: Negativ | FALSE | NOT_IDENTIFIE | 6646 | 0.01134918 | 0.002142618 | 0.002142618 | 0.002142618 | 0.002142618 | 0 | 0 | 5.836050443 | VALID | 29869 | 0.033064 | 29870 | 3259356417 | 2484 | 51190 |
| Negative-Control-2_S26 | NTC-2: Negativ | FALSE | NOT_IDENTIFIE | 8108 | 0.014362236 | 0.010244392 | 0.010244392 | 0.007231336 | 0.007231336 | 0.002075661 | 0.010244392 | 6.071507255 | VALID | 29869 | 0.310643 | 29870 | 3259356417 | 3539 | 49276 |
| VeroE6-1_S36_L001_R1 | NEC-1: Negativ | FALSE | NOT_IDENTIFIE | 12259584 | 0 | 0 | 0 | 0 | 0 | 0 | 0 | #NAME? | VALID | 29869 | 0 | 29870 | 3259356417 | 12052879 | 27387434 |
| VeroE6-2_S37_L001_R1 | NEC-2: Negativ | FALSE | NOT_IDENTIFIE | 14591058 | 0 | 0 | 0 | 0 | 0 | 0 | 0 | #NAME? | VALID | 29869 | 0 | 29870 | 3259356417 | 14388229 | 24278506 |
| VeroE6-3_S38_L001_R1 | NEC-3: Negativ | FALSE | NOT_IDENTIFIE | 15735310 | 0.008938735 | 0 | 0 | 0 | 0 | 0 | 0 | 5.597298994 | VALID | 29869 | 0 | 29870 | 3259356417 | 15423512 | 53266016 |
| VTM-1-Negative_S33_L | NTC-3: Negativ | TRUE | 20B | 16824 | 0.999765651 | 0.991965182 | 0.971108135 | 0.035152327 | 0 | 0 | 0.831348542 | 10.31195526 | VALID | 29869 | 0.821609 | 29870 | 3259356417 | 6499 | 45698 |
| VTM-2-Negative_S34_L | NTC-4: Negativ | FALSE | NOT_IDENTIFIE | 7856 | 0.003280884 | 0.003280884 | 0.003280884 | 0 | 0 | 0 | 0 | 4.595017815 | VALID | 29869 | 0.001854 | 29870 | 3259356417 | 5396 | 74396 |
| VTM-3-Negative_S35_L | NTC-5: Negativ | FALSE | NOT_IDENTIFIE | 38570 | 0 | 0 | 0 | 0 | 0 | 0 | 0 | #NAME? | VALID | 29869 | 0 | 29870 | 3259356417 | 29062 | 15525 |

| Specimen_ID | Copy_number | presence | clade | reads | X1 | X5 | X10 | X50 | X100 | X500 | E1 | I1 | validity | BAIT_TERRITO | PCT_SELECTED | TARGET_TERRI | GENOME_SIZE | TOTAL_READS | HUMAN_ ON_TARGET_BASES |
|---|---|---|---|---|---|---|---|---|---|---|---|---|---|---|---|---|---|---|---|
| 200-10_S21_L001_R1_0 | 800_copies_pe | TRUE | 19B | 19567791 | 0.732407097 | 0.55125544 | 0.38781386 | 0.013157014 | 0 | 0 | 0.511671409 | 10.00267584 | VALID | 29869 | 0.000246 | 29870 | 3259356417 | 11132676 | 122940840 |
| 200-11_S22_L001_R1_0 | 800_copies_pe | TRUE | 19B | 1321419 | 0.898225644 | 0.816672246 | 0.701171744 | 0.070304653 | 0.001841312 | 0 | 0.632462337 | 10.20635062 | VALID | 29869 | 0.008107 | 29870 | 3259356417 | 776735 | 8384372 |
| 200-12_S23_L001_R1_0 | 800_copies_pe | TRUE | 19B | 1179898 | 0.820120522 | 0.718848343 | 0.596417811 | 0.040073652 | 0 | 0 | 0.596484767 | 10.11401084 | VALID | 29869 | 0.005934 | 29870 | 3259356417 | 763835 | 16902585 |
| 200-13_S24_L001_R1_0 | 800_copies_pe | TRUE | NOT_IDENTIFIE | 45998887 | 0.565952461 | 0.017241379 | 0 | 0 | 0 | 0 | 0 | 9.74433414 | VALID | 29869 | 7.00E-06 | 29870 | 3259356417 | 37378484 | 881833606 |
| 200-14_S25_L001_R1_0 | 800_copies_pe | TRUE | 19A | 10620613 | 0.895882156 | 0.440207566 | 0.085805156 | 0 | 0 | 0 | 0.556957929 | 10.20361064 | VALID | 29869 | 0.000128 | 29870 | 3259356417 | 10244131 | 98843845 |
| 200-15_S26_L001_R1_0 | 800_copies_pe | TRUE | 19B | 32055937 | 0.997120857 | 0.996551724 | 0.995647807 | 0.947639772 | 0.737562772 | 0.000100435 | 0.774707368 | 10.30978657 | VALID | 29869 | 0.001398 | 29870 | 3259356417 | 30833828 | 118540231 |
| 200-16_S27_L001_R1_0 | 800_copies_pe | TRUE | 19B | 1203350 | 0.992735186 | 0.97562772 | 0.916136592 | 0.234315367 | 0.009206562 | 0 | 0.749891688 | 10.30502615 | VALID | 29869 | 0.007983 | 29870 | 3259356417 | 1170632 | 4507215 |
| 200-17_S28_L001_R1_0 | 800_copies_pe | TRUE | 19A | 12491956 | 0.958151992 | 0.629293606 | 0.281352528 | 0 | 0 | 0 | 0.630010044 | 10.27089697 | VALID | 29869 | 0.000165 | 29870 | 3259356417 | 11999480 | 107464020 |
| 200-18_S29_L001_R1_0 | 800_copies_pe | TRUE | 19A | 11610093 | 0.960060261 | 0.599933043 | 0.221057918 | 0 | 0 | 0 | 0.653381319 | 10.27172658 | VALID | 29869 | 0.000158 | 29870 | 3259356417 | 11225387 | 96433434 |
| 200-19_S30_L001_R1_0 | 800_copies_pe | TRUE | 19A | 15005488 | 0.982055574 | 0.851724138 | 0.633143622 | 0 | 0 | 0 | 0.716626594 | 10.29503167 | VALID | 29869 | 0.000231 | 29870 | 3259356417 | 14475151 | 171534027 |
| 200-1_S12_L001_R1_00 | 800_copies_pe | TRUE | 19A | 17638397 | 0.822932708 | 0.662939404 | 0.436826247 | 0.008637429 | 0 | 0 | 0.571662389 | 10.11959632 | VALID | 29869 | 0.000338 | 29870 | 3259356417 | 9636299 | 67754989 |
| 200-20_S31_L001_R1_0 | 800_copies_pe | TRUE | 19A | 15092369 | 0.918245732 | 0.630498828 | 0.295279545 | 0 | 0 | 0 | 0.599760071 | 10.22757937 | VALID | 29869 | 0.000143 | 29870 | 3259356417 | 14573562 | 157934041 |
| 200-21_S32_L001_R1_0 | 800_copies_pe | TRUE | 19A | 15481079 | 0.871543355 | 0.727017074 | 0.512052226 | 0.000870439 | 0 | 0 | 0.618968865 | 10.17622902 | VALID | 29869 | 0.000287 | 29870 | 3259356417 | 10663574 | 112019872 |
| 200-22_S33_L001_R1_0 | 800_copies_pe | FALSE | NOT_IDENTIFIE | 8328941 | 0.099531302 | 0.024338801 | 0.000870439 | 0 | 0 | 0 | 0 | 7.996611816 | VALID | 29869 | 1.10E-05 | 29870 | 3259356417 | 7920723 | 580114419 |
| 200-23_S34_L001_R1_0 | 800_copies_pe | TRUE | NOT_IDENTIFIE | 28285717 | 0.18928691 | 0 | 0 | 0 | 0 | 0 | 0 | 8.64621916 | VALID | 29869 | 3.00E-06 | 29870 | 3259356417 | 20517187 | 543105407 |
| 200-24_S35_L001_R1_0 | 800_copies_pe | TRUE | 19B | 1390054 | 0.992902578 | 0.969601607 | 0.891998661 | 0.067124205 | 0.000502176 | 0 | 0.761192947 | 10.3061275 | VALID | 29869 | 0.007254 | 29870 | 3259356417 | 992758 | 8708540 |
| 200-25_S36_L001_R1_0 | 800_copies_pe | TRUE | 19B | 11118947 | 0.994442585 | 0.972380315 | 0.933980583 | 0.16324071 | 0.00063609 | 0 | 0.771242049 | 10.30715344 | VALID | 29869 | 0.00104 | 29870 | 3259356417 | 8499394 | 54170658 |
| 200-26_S37_L001_R1_0 | 800_copies_pe | TRUE | 19B | 1161004 | 0.958151992 | 0.90756612 | 0.80860395 | 0.057248075 | 0.001272179 | 0 | 0.721742551 | 10.26980614 | VALID | 29869 | 0.007307 | 29870 | 3259356417 | 829972 | 8319691 |
| 200-27_S38_L001_R1_0 | 800_copies_pe | TRUE | 19A | 17007769 | 0.860964178 | 0.289320388 | 0.038634081 | 0 | 0 | 0 | 0.468011383 | 10.16475324 | VALID | 29869 | 8.20E-05 | 29870 | 3259356417 | 12352019 | 282490493 |
| 200-28_S39_L001_R1_0 | 800_copies_pe | TRUE | NOT_IDENTIFIE | 21146650 | 0.259758955 | 0 | 0 | 0 | 0 | 0 | 0 | 8.966659075 | VALID | 29869 | 6.00E-06 | 29870 | 3259356417 | 15495553 | 445292801 |
| 200-29_S40_L001_R1_0 | 800_copies_pe | TRUE | 19B | 10058214 | 0.98570472 | 0.98339471 | 0.981854704 | 0.964445932 | 0.926715768 | 0.027753599 | 0.827016962 | 10.29772526 | VALID | 29869 | 0.009916 | 29870 | 3259356417 | 9810839 | 259157734 |
| 200-2_S13_L001_R1_00 | 800_copies_pe | TRUE | 19A | 12677581 | 0.548041513 | 0.396819551 | 0.223870104 | 0 | 0 | 0 | 0.421367593 | 9.711886712 | VALID | 29869 | 0.00018 | 29870 | 3259356417 | 8016402 | 281079062 |
| 200-30_S41_L001_R1_0 | 800_copies_pe | TRUE | 19B | 14551071 | 0.986709073 | 0.969601607 | 0.914797456 | 0.129394041 | 0 | 0 | 0.763404515 | 10.29925975 | VALID | 29869 | 0.000576 | 29870 | 3259356417 | 14305969 | 790497424 |
| 200-3_S14_L001_R1_00 | 800_copies_pe | TRUE | NOT_IDENTIFIE | 28464812 | 0.248242384 | 0 | 0 | 0 | 0 | 0 | 0 | 8.92131059 | VALID | 29869 | 3.00E-06 | 29870 | 3259356417 | 28292444 | 645863434 |
| 200-4_S15_L001_R1_00 | 800_copies_pe | TRUE | 19A | 18027697 | 0.821694007 | 0.64978239 | 0.412621359 | 0.004753934 | 0 | 0 | 0.567785403 | 10.1164547 | VALID | 29869 | 0.000133 | 29870 | 3259356417 | 17927625 | 225429392 |
| 200-5_S16_L001_R1_00 | 800_copies_pe | TRUE | NOT_IDENTIFIE | 13619534 | 0.58948778 | 0.145363241 | 0.01155005 | 0 | 0 | 0 | 0.311416137 | 9.783694646 | VALID | 29869 | 4.00E-05 | 29870 | 3259356417 | 13512783 | 268724949 |
| 200-6_S17_L001_R1_00 | 800_copies_pe | TRUE | NOT_IDENTIFIE | 18334099 | 0.20860395 | 0.000167392 | 0 | 0 | 0 | 0 | 0 | 8.747342448 | VALID | 29869 | 4.00E-06 | 29870 | 3259356417 | 18234520 | 415781789 |
| 200-7_S18_L001_R1_00 | 800_copies_pe | TRUE | NOT_IDENTIFIE | 17140943 | 0.656913291 | 0.111817877 | 0.0058922 | 0 | 0 | 0 | 0.284365584 | 9.892913859 | VALID | 29869 | 3.10E-05 | 29870 | 3259356417 | 17011770 | 248862820 |
| 200-8_S19_L001_R1_00 | 800_copies_pe | TRUE | 19A | 15823930 | 0.763207231 | 0.336625377 | 0.092869099 | 0 | 0 | 0 | 0.478724473 | 10.04440834 | VALID | 29869 | 6.40E-05 | 29870 | 3259356417 | 15727502 | 300419232 |
| 200-9_S20_L001_R1_00 | 800_copies_pe | TRUE | 19A | 1015377 | 0.652025444 | 0.56344158 | 0.460528959 | 0.02962839 | 0 | 0 | 0.505350458 | 9.885474396 | VALID | 29869 | 0.00591 | 29870 | 3259356417 | 629856 | 8206781 |
| COVID-NEG-032_S7_L0 | CLINICAL_MAT | FALSE | NOT_IDENTIFIE | 24082540 | 0.004921326 | 0 | 0 | 0 | 0 | 0 | 0 | 5.000482923 | VALID | 29869 | 0 | 29870 | 3259356417 | 23172209 | 215139876 |
| COVID-NEG-033_S8_L0 | CLINICAL_MAT | FALSE | NOT_IDENTIFIE | 18917582 | 0.145329762 | 0.095915634 | 0.040274523 | 0 | 0 | 0 | 0.126615333 | 8.385910351 | VALID | 29869 | 1.30E-05 | 29870 | 3259356417 | 18805063 | 251941381 |
| COVID-NEG-034_S9_L0 | CLINICAL_MAT | FALSE | NOT_IDENTIFIE | 12325773 | 0 | 0 | 0 | 0 | 0 | 0 | 0 | #NAME? | VALID | 29869 | 0 | 29870 | 3259356417 | 6924478 | 67630669 |
| COVID-NEG-035_S10_L | CLINICAL_MAT | FALSE | NOT_IDENTIFIE | 14378979 | 0 | 0 | 0 | 0 | 0 | 0 | 0 | #NAME? | VALID | 29869 | 0 | 29870 | 3259356417 | 9823676 | 77295770 |
| COVID-NEG-036_S11_L | CLINICAL_MAT | FALSE | NOT_IDENTIFIE | 10888882 | 0 | 0 | 0 | 0 | 0 | 0 | 0 | #NAME? | VALID | 29869 | 0 | 29870 | 3259356417 | 10588610 | 316362634 |
| VTM-1_S1_L001_R1_00 | NTC-1: Negativ | FALSE | NOT_IDENTIFIE | 232 | 0 | 0 | 0 | 0 | 0 | 0 | 0 | #NAME? | VALID | 29869 | 0 | 29870 | 3259356417 | 171 | 4411 |
| VTM-2_S2_L001_R1_00 | NTC-2: Negativ | FALSE | NOT_IDENTIFIE | 1841 | 0 | 0 | 0 | 0 | 0 | 0 | 0 | #NAME? | VALID | 29869 | 0 | 29870 | 3259356417 | 1412 | 22687 |
| VTM-3_S3_L001_R1_00 | NTC-3: Negativ | FALSE | NOT_IDENTIFIE | 341 | 0 | 0 | 0 | 0 | 0 | 0 | 0 | #NAME? | VALID | 29869 | 0 | 29870 | 3259356417 | 170 | 4806 |
| VeroE6-1_S4_L001_R1_ | NEC-1: Negativ | FALSE | NOT_IDENTIFIE | 23958030 | 0 | 0 | 0 | 0 | 0 | 0 | 0 | #NAME? | VALID | 29869 | 0 | 29870 | 3259356417 | 23621777 | 45537443 |
| VeroE6-2_S5_L001_R1_ | NEC-2: Negativ | FALSE | NOT_IDENTIFIE | 10102253 | 0 | 0 | 0 | 0 | 0 | 0 | 0 | #NAME? | VALID | 29869 | 0 | 29870 | 3259356417 | 9994035 | 5395262 |
| VeroE6-3_S6_L001_R1_ | NEC-3: Negativ | FALSE | NOT_IDENTIFIE | 18004012 | 0 | 0 | 0 | 0 | 0 | 0 | 0 | #NAME? | VALID | 29869 | 0 | 29870 | 3259356417 | 17627828 | 36860317 |

**Supplemental Table 5. In-silico studies inclusivity and exclusivity study included GISAID, NCBI Viral and a combined genome dataset.** The total number of sequences with different percent identity is shown in the table. To account for potential cross-reactivity of the SARS-CoV-2 NGS hybridization primers, we aligned reads to 29 microbial genomes and the human genome along with the SARS-CoV-2 genome. All pathogens were determined to have no cross-reactivity (no more than 80% homology) with the primers used, except human coronavirus HKU1 with 3 out of 994 probes with 84.6% homology.

| **DATABASE** | **GISAID** | **NCBI Virus** | **Combined Genome** |
|---|---|---|---|
| Date | **2/12/21** | **2/16/21** | **2/16/21** |
| Total Sequences Reviewed | 22,544 | 3,603,771 | 31 |
| Unique Sequences Reviewed | 16,503 | 3,603,771 | 31 |
| Sequences with 100% Mean Percent Identity | 6,200 | 50,200 | 1 |
| Sequences with >= 80% Mean Percent Identity | 16,503 | 65,747 | 2 |
| Sequences with < 80% Mean Percent Identity | 0 | 3,538,024 | 29 |
| Sequences with 0% Mean Percent Identity | 0 | 3,538,023 | 29 |
| **Pathogens in Combined Genome** | **Accession** | **Homology Above 80%** | |
| Adenovirus (e.g. C1 Ad. 71) | NC_001405.1 | 0 | |
| Human Metapneumovirus (hMPV) | NC_039199.1 | 0 | |
| Parainfluenza virus 1 | JQ901971.1 | 0 | |
| Parainfluenza virus 2 | NC_003443.1 | 0 | |
| Parainfluenza virus 3 | NC_001796.2 | 0 | |
| Parainfluenza virus 4 | KF483663.1 | 0 | |
| Influenza A | AB284320.1 | 0 | |
| Influenza B | NC_002208.1 | 0 | |
| Enterovirus (e.g. EV68) | AY426531.1 | 0 | |
| Respiratory syncytial virus | NC_038235.1 | 0 | |
| Rhinovirus | ENA\|L24917\|L24917.1 | 0 | |
| Haemophilus influenzae | Haemophilus_influenzae_ATCC_51907 | 0 | |
| Legionella pneumophila | Legionella_pneumophila_subsp_pneumophila_ATCC_33152 | 0 | |
| Mycobacterium tuberculosis | NC_000962.3 | 0 | |
| Streptococcus pneumoniae | Streptococcus_pneumoniae_ATCC_700669 | 0 | |
| Streptococcus pyogenes | Streptococcus_pyogenes_ATCC_12344 | 0 | |
| Mycoplasma pneumoniae | NZ_CP010546.1 | 0 | |
| Candida albicans | 1,CP017628.1,CP017629.1,CP017630.1 | 0 | |
| Pseudomonas aeruginosa | Pseudomonas_aeruginosa_ATCC_9027 | 0 | |
| Staphylococcus epidermidis | Staphylococcus_epidermidis_ATCC_12228 | 0 | |
| Staphylococcus salivarius | Streptococcus_salivarius_ATCC_9759 | 0 | |
| Human coronavirus 229E | NC_002645.1 | 0 | |
| Human coronavirus OC43 | NC_006213.1 | 0 | |
| Human coronavirus HKU1 | NC_006577.2 | 3 | |
| Human coronavirus NL63 | DQ445911.1 | 0 | |
| MERS-coronavirus | KT006149.2 | 0 | |
| Chlamydia pneumonia | NC_005043.1 | 0 | |
| Pneumocystis jirovecii (PCP) | GCA_001477535.1 | 0 | |
| Bordetella pertussis | CP011448.1 | 0 | |
| Human Genome | GrC38 | 0 | |

**Supplemental Table 6. NGS Variant Analysis and Annotation of 71 COVID Positive Samples.** We detected 733 mutations (261 different mutations) at 258 different mutation sites. Out of these mutations, 153 were previosuly reported and we identified 107 new mutations. The annotation provides information on the mutation name, gene location, protein name, amino acid change (NCBI) and synonymy. We provided information on the frequency of genetic variants in our sample cohort andthe associated read depth. We used the Grantham scoring system to designate conservative and radical mutations. In this system the score of 100 and above calls mutations radical.

| Mutation_Name | Sample_Frequencies | Read_Depth | Gene | Protein_Name | AA_Change_NCBI | Synonymy | Grantham Score | Radical >100 | Reported in GISAID (11-11-2020) | Notes |
|---|---|---|---|---|---|---|---|---|---|---|
| C222T | 2 | 31, 43 | 5'UTR | NA | extragenic | NA | NA | NA | Reported | |
| C228T | 1 | 30 | 5'UTR | NA | extragenic | NA | NA | NA | Reported | |
| C241T | 67 | 3-483 | 5'UTR | NA | extragenic | NA | NA | NA | Reported | |
| C335T | 1 | 235 | Orf1ab | NSP1 | R24C | non-synonymous | 180 | radical | Reported | |
| C478T | 1 | 310 | Orf1ab | NSP1 | I71I | synonymous | NA | NA | Reported | |
| C601T | 1 | 430 | Orf1ab | NSP1 | G112G | synonymous | NA | NA | Reported | |
| C683T | 1 | 438 | Orf1ab | NSP1 | L140L | synonymous | NA | NA | Reported | |
| T908C | 1 | 11 | Orf1ab | NSP2 | L215L | synonymous | NA | NA | New | |
| C936T | 1 | 6 | Orf1ab | NSP2 | T224I | non-synonymous | 89 | conservative | Reported | |
| C1059T | 57 | 3-484 | Orf1ab | NSP2 | T265I | non-synonymous | 89 | conservative | Reported | |
| A1133T | 2 | 5, 8 | Orf1ab | NSP2 | K290_stop | non-synonymous | NA | NA | New | |
| C1170T | 1 | 417 | Orf1ab | NSP2 | S302F | non-synonymous | 155 | radical | Reported | |
| C1392T | 1 | 114 | Orf1ab | NSP2 | S376L | non-synonymous | 145 | radical | Reported | |
| G1542T | 1 | 457 | Orf1ab | NSP2 | R426L | non-synonymous | 102 | radical | New | |
| A1631G | 1 | 14 | Orf1ab | NSP2 | K456E | non-synonymous | 56 | conservative | New | |
| A1669T | 1 | 40 | Orf1ab | NSP2 | K468N | non-synonymous | 94 | conservative | New | |
| T1671A | 1 | 8 | Orf1ab | NSP2 | L469H | non-synonymous | 99 | conservative | New | |
| G1820A | 1 | 81 | Orf1ab | NSP2 | G519S | non-synonymous | 56 | conservative | Reported | |
| A1900G | 1 | 456 | Orf1ab | NSP2 | R545R | synonymous | NA | NA | New | |
| C1917T | 1 | 453 | Orf1ab | NSP2 | T551I | non-synonymous | 89 | conservative | Reported | |
| T1927C | 1 | 109 | Orf1ab | NSP2 | T554T | synonymous | NA | NA | Reported | |
| G1942T | 1 | 455 | Orf1ab | NSP2 | V559V | synonymous | NA | NA | Reported | |
| C2048T | 1 | 493 | Orf1ab | NSP2 | L595L | synonymous | NA | NA | Reported | |
| C2106T | 1 | 445 | Orf1ab | NSP2 | T614I | non-synonymous | 89 | conservative | Reported | |
| G2204A | 1 | 473 | Orf1ab | NSP2 | E647K | non-synonymous | 56 | conservative | New | |
| C2363T | 1 | 480 | Orf1ab | NSP2 | L700F | non-synonymous | 22 | conservative | Reported | |
| G2374A | 1 | 492 | Orf1ab | NSP2 | L703L | synonymous | NA | NA | New | |
| C2508T | 1 | 466 | Orf1ab | NSP2 | P748L | non-synonymous | 98 | conservative | Reported | |
| G2632T | 1 | 492 | Orf1ab | NSP2 | M789I | non-synonymous | 10 | conservative | Reported | |
| C3037T | 62 | 2-726 | Orf1ab | NSP3 | F924F | synonymous | NA | NA | Reported | |
| G3248A | 1 | 15 | Orf1ab | NSP3 | E995K | non-synonymous | 56 | conservative | New | |
| C3411T | 7 | 16-482 | Orf1ab | NSP3 | A1049V | non-synonymous | 64 | conservative | Reported | |
| A3427C | 1 | 282 | Orf1ab | NSP3 | P1054P | synonymous | NA | NA | New | |
| C3486T | 1 | 434 | Orf1ab | NSP3 | A1074V | non-synonymous | 64 | conservative | New | |
| T3657A | 1 | 9 | Orf1ab | NSP3 | L1131H | non-synonymous | 99 | conservative | New | |
| T3713C | 1 | 7 | Orf1ab | NSP3 | S1150P | non-synonymous | 74 | conservative | New | |
| C3737T | 1 | 51 | Orf1ab | NSP3 | P1158S | non-synonymous | 74 | conservative | Reported | |
| T3741C | 1 | 9 | Orf1ab | NSP3 | I1159T | non-synonymous | 89 | conservative | Reported | |
| G3871A | 1 | 5 | Orf1ab | NSP3 | K1202K | synonymous | NA | NA | Reported | |
| A4083G | 1 | 3 | Orf1ab | NSP3 | D1273G | non-synonymous | 94 | conservative | New | |
| C4113T | 4 | 423-459 | Orf1ab | NSP3 | A1283V | non-synonymous | 64 | conservative | Reported | |
| A4197G | 4 | 14-499 | Orf1ab | NSP3 | E1311G | non-synonymous | 98 | conservative | Reported | |
| T4342C | 1 | 274 | Orf1ab | NSP3 | I1359I | synonymous | NA | NA | New | |
| C4540T | 1 | 2 | Orf1ab | NSP3 | Y1423Y | synonymous | NA | NA | Reported | |
| T4619A | 1 | 3 | Orf1ab | NSP3 | Y1452N | non-synonymous | 143 | radical | New | |
| A4620T | 1 | 3 | Orf1ab | NSP3 | Y1452F | non-synonymous | 22 | conservative | New | |
| A5114G | 1 | 18 | Orf1ab | NSP3 | T1617A | non-synonymous | 58 | conservative | New | |
| T5471del (1) | 1 | 44 | Orf1ab | NSP3 | F1736F | Frameshift after synonymous | NA | radical | New | (1) GT5470G or T5471-deletion causes |
| C5497T | 1 | 471 | Orf1ab | NSP3 | C1744C | synonymous | NA | NA | Reported | |
| C5512T | 1 | 19 | Orf1ab | NSP3 | N1749N | synonymous | NA | NA | Reported | |
| A5533G | 1 | 57 | Orf1ab | NSP3 | G1756G | synonymous | NA | NA | Reported | |
| C5736T | 1 | 8 | Orf1ab | NSP3 | A1824V | non-synonymous | 64 | conservative | Reported | |
| C5777T | 1 | 10 | Orf1ab | NSP3 | H1838Y | non-synonymous | 83 | conservative | New | |
| G5830A | 1 | 443 | Orf1ab | NSP3 | K1855K | synonymous | NA | NA | Reported | |
| C6070T | 1 | 479 | Orf1ab | NSP3 | I1935I | synonymous | NA | NA | Reported | |
| A6131G | 1 | 36 | Orf1ab | NSP3 | K1956E | non-synonymous | 56 | conservative | New | |
| A6295G | 2 | 19-468 | Orf1ab | NSP3 | I2010M | non-synonymous | 10 | conservative | Reported | |
| A6324del (2) | 1 | 39 | Orf1ab | NSP3 | E2020E | frameshift after synonymous | Frameshift after NA | radical | New | (2) GA6323G or T6324-deletion causes |
| T6394C | 7 | 20-473 | Orf1ab | NSP3 | D2043D | synonymous | NA | NA | Reported | |
| C6629T | 1 | 4 | Orf1ab | NSP3 | L2122F | non-synonymous | 22 | conservative | Reported | |
| C6706T | 1 | 465 | Orf1ab | NSP3 | N2147N | synonymous | NA | NA | Reported | |
| C6720T | 1 | 53 | Orf1ab | NSP3 | T2152I | non-synonymous | 89 | conservative | New | |
| T6763C | 1 | 13 | Orf1ab | NSP3 | T2166T | synonymous | NA | NA | New | |
| A6785G | 1 | 5 | Orf1ab | NSP3 | T2174A | non-synonymous | 58 | conservative | New | |
| A6832T | 1 | 8 | Orf1ab | NSP3 | R2189S | non-synonymous | 110 | radical | Reported | |
| G6884A | 2 | 244, 473 | Orf1ab | NSP3 | G2207S | non-synonymous | 56 | conservative | Reported | |
| G6885T | 1 | 464 | Orf1ab | NSP3 | G2207V | non-synonymous | 109 | radical | New | |
| T6949A | 1 | 14 | Orf1ab | NSP3 | N2228K | non-synonymous | 94 | conservative | Reported | |
| C7005T | 1 | 86 | Orf1ab | NSP3 | T2247I | non-synonymous | 89 | conservative | New | |
| C7113T | 1 | 465 | Orf1ab | NSP3 | T2283I | non-synonymous | 89 | conservative | Reported | |
| C7119del (3) | 1 | 3 | Orf1ab | NSP3 | S2285L | non-synonymous | 145 | radical | New | (3) TC7118T or C7119-deletion causes |
| A7309T | 1 | 17 | Orf1ab | NSP3 | Q2348H | non-synonymous | 24 | conservative | New | |
| C8025T | 1 | 475 | Orf1ab | NSP3 | A2587V | non-synonymous | 64 | conservative | Reported | |
| C8078T | 3 | 206-457 | Orf1ab | NSP3 | P2605S | non-synonymous | 74 | conservative | Reported | |
| G8111A | 1 | 3 | Orf1ab | NSP3 | A2616T | non-synonymous | 58 | conservative | New | |
| G8179A | 4 | 7-485 | Orf1ab | NSP3 | R2638R | synonymous | NA | NA | Reported | |
| C8389T | 1 | 313 | Orf1ab | NSP3 | N2708N | synonymous | NA | NA | Reported | |
| A8446T | 1 | 15 | Orf1ab | NSP3 | K2727N | non-synonymous | 94 | conservative | Reported | |
| C8782T | 1 | 37 | Orf1ab | NSP4 | S2338S | synonymous | NA | NA | Reported | |
| C8818T | 1 | 28 | Orf1ab | NSP4 | C2851C | synonymous | NA | NA | Reported | |
| A9409T | 1 | 483 | Orf1ab | NSP4 | V3048V | synonymous | NA | NA | Reported | |
| A9409G | 1 | 14 | Orf1ab | NSP4 | V3048V | synonymous | NA | NA | Reported | |
| T9444G | 1 | 17 | Orf1ab | NSP4 | L3060R | non-synonymous | 102 | radical | New | |
| G9460T | 1 | 75 | Orf1ab | NSP4 | M3065I | non-synonymous | 10 | conservative | New | |
| C9893T | 1 | 105 | Orf1ab | NSP4 | L3210F | non-synonymous | 22 | conservative | Reported | |
| C9969T | 1 | 458 | Orf1ab | NSP4 | A3235V | non-synonymous | 64 | conservative | Reported | |
| C10039T | 1 | 468 | Orf1ab | NSP4 | T3258T | synonymous | NA | NA | Reported | |
| C10319T | 7 | 79-487 | Orf1ab | NSP5 | L3352F | non-synonymous | 22 | conservative | Reported | |
| C10340T | 1 | 61 | Orf1ab | NSP5 | P3359S | non-synonymous | 74 | conservative | Reported | |
| G10481A | 1 | 11 | Orf1ab | NSP5 | G3406S | non-synonymous | 56 | conservative | New | |
| C10647T | 1 | 35 | Orf1ab | NSP5 | T3461I | non-synonymous | 89 | conservative | Reported | |
| G10754A | 1 | 408 | Orf1ab | NSP5 | A3497T | non-synonymous | 58 | conservative | New | |
| C10755T | 1 | 4 | Orf1ab | NSP5 | A3497V | non-synonymous | 64 | conservative | Reported | |
| C10851T | 2 | 51, 361 | Orf1ab | NSP5 | A3529V | non-synonymous | 64 | conservative | Reported | |
| C10956T | 1 | 2 | Orf1ab | NSP5 | S3564L | non-synonymous | 145 | radical | New | |
| G11083T | 2 | 215, 369 | Orf1ab | NSP5 | L3606F | non-synonymous | 22 | conservative | Reported | |
| C11224T | 1 | 160 | Orf1ab | NSP5 | V3653V | synonymous | NA | NA | Reported | |
| G11335T | 1 | 501 | Orf1ab | NSP5 | V3690V | synonymous | NA | NA | Reported | |
| G11382A | 1 | 43 | Orf1ab | NSP5 | R3706K | non-synonymous | 26 | conservative | New | |
| T11459A | 1 | 15 | Orf1ab | NSP5 | S3732T | non-synonymous | 58 | conservative | New | |
| C11511G | 1 | 5 | Orf1ab | NSP6 | T3749R | non-synonymous | 71 | conservative | Reported | |
| A11792T (4) | 1 | 10 | Orf1ab | NSP6 | K3843-STOP | non-synonymous | STOP codon | radical | New | (4) The stop codon resulting in a truncated |
| C11916T | 15 | 164-512 | Orf1ab | NSP7/Replicase | S3884L | non-synonymous | 145 | radical | Reported | |
| G12806A | 1 | 13 | Orf1ab | NSP9/Replicase | V4181I | non-synonymous | 29 | conservative | New | |
| T12879C | 1 | 61 | Orf1ab | NSP9/Replicase | I4205T | non-synonymous | 89 | conservative | New | |
| T12953C | 1 | 24 | Orf1ab | NSP9/Replicase | F4230L | non-synonymous | 22 | conservative | New | |
| T13000del (5) | 1 | 6 | Orf1ab | NSP9/Replicase | S4245S | synonymous | NA | radical | New | (5) GT12999G or T13000-deletion causes |
| C13368T | 1 | 3 | Orf1ab | NSP10/RNA synthase | T4368I | non-synonymous | 89 | conservative | New | |
| C13541T | 1 | 34 | Orf1ab | NSP12/RNAdepRNApol | A4426V | non-synonymous | 64 | conservative | New | |
| C13568A | 1 | 8 | Orf1ab | NSP12/RNAdepRNApol | A4435D | non-synonymous | 126 | radical | Reported | |
| C13643T | 1 | 23 | Orf1ab | NSP12/RNAdepRNApol | S4460F | non-synonymous | 155 | radical | New | |
| C13821T | 2 | 468, 473 | Orf1ab | NSP12/RNAdepRNApol | L4519L | synonymous | NA | NA | Reported | |
| CT13829C | 1 | 3 | Orf1ab | NSP12/RNAdepRNApol | A4522V | non-synonymous | 64 | conservative | New | |
| T13914C | 1 | 462 | Orf1ab | NSP12/RNAdepRNApol | N4550N | synonymous | NA | NA | Reported | |
| T14190A | 1 | 47 | Orf1ab | NSP12/RNAdepRNApol | A4642A | synonymous | NA | NA | New | |
| T14232C | 1 | 3 | Orf1ab | NSP12/RNAdepRNApol | P4656P | synonymous | NA | NA | New | |
| C14408T | 63 | 5-498 | Orf1ab | NSP12/RNAdepRNApol | P4715L | non-synonymous | 98 | conservative | Reported | |
| C14422A | 1 | 18 | Orf1ab | NSP12/RNAdepRNApol | P4720T | non-synonymous | 38 | conservative | Reported | |
| A14533C | 1 | 41 | Orf1ab | NSP12/RNAdepRNApol | R4757R | synonymous | NA | NA | New | |
| C14708T | 1 | 6 | Orf1ab | NSP12/RNAdepRNApol | A4815V | non-synonymous | 64 | conservative | Reported | |
| C14805T | 2 | 273, 432 | Orf1ab | NSP12/RNAdepRNApol | Y4847Y | synonymous | NA | NA | Reported | |
| C15277T | 1 | 332 | Orf1ab | NSP12/RNAdepRNApol | H5005Y | non-synonymous | 83 | conservative | Reported | |
| G15444T | 1 | 466 | Orf1ab | NSP12/RNAdepRNApol | M5060I | non-synonymous | 10 | conservative | Reported | |
| T15539C | 1 | 23 | Orf1ab | NSP12/RNAdepRNApol | V5092A | non-synonymous | 64 | conservative | New | |
| G15906T | 2 | 11, 447 | Orf1ab | NSP12/RNAdepRNApol | Q5214H | non-synonymous | 24 | conservative | Reported | |
| C15924T | 4 | 11-481 | Orf1ab | NSP12/RNAdepRNApol | Y5220Y | synonymous | NA | NA | Reported | |
| A15992C | 1 | 65 | Orf1ab | NSP12/RNAdepRNApol | D5243A | non-synonymous | 126 | radical | New | |
| A16104C | 1 | 19 | Orf1ab | NSP12/RNAdepRNApol | I5280I | synonymous | NA | NA | New | |

| | | | | | | | | | | |
|---|---|---|---|---|---|---|---|---|---|---|
| **C16208T** | 1 | 472 | Orf1ab | NSP12/RNAdepRNApol | A5315V | non-synonymous | 64 | conservative | New | |
| **C16260T** | 1 | 452 | Orf1ab | NSP13/Helicase | C5332C | synonymous | NA | NA | Reported | |
| **C16393T** | 1 | 12 | Orf1ab | NSP13/Helicase | P5377S | non-synonymous | 74 | conservative | Reported | |
| **C16420T (6)** | 1 | 9 | Orf1ab | NSP13/Helicase | Q5386-STOP | non-synonymous | NA | radical | New | (6) The stop codon creates a truncated |
| **C16498A** | 1 | 8 | Orf1ab | NSP13/Helicase | Q5412K | non-synonymous | 53 | conservative | New | |
| **G16945T** | 1 | 5 | Orf1ab | NSP13/Helicase | A5561S | non-synonymous | 99 | conservative | Reported | |
| **T17247C** | 2 | 429, 479 | Orf1ab | NSP13/Helicase | R5661R | synonymous | NA | NA | Reported | |
| **T17543A** | 1 | 35 | Orf1ab | NSP13/Helicase | M5760K | non-synonymous | 95 | conservative | New | |
| **T17549G** | 1 | 34 | Orf1ab | NSP13/Helicase | L5762R | non-synonymous | 102 | radical | New | |
| **C17550A** | 1 | 34 | Orf1ab | NSP13/Helicase | L5762L | synonymous | NA | NA | Reported | |
| **C17550T** | 1 | 32 | Orf1ab | NSP13/Helicase | L5762L | synonymous | NA | NA | Reported | |
| **T17556A** | 1 | 34 | Orf1ab | NSP13/Helicase | T5764T | synonymous | NA | NA | New | |
| **C17746T** | 1 | 10 | Orf1ab | NSP13/Helicase | P5828S | non-synonymous | 74 | conservative | Reported | |
| **C17747T** | 1 | 262 | Orf1ab | NSP13/Helicase | P5828L | non-synonymous | 98 | conservative | Reported | |
| **A17858G** | 1 | 106 | Orf1ab | NSP13/Helicase | Y5865C | non-synonymous | 194 | radical | Reported | |
| **G18025T** | 1 | 5 | Orf1ab | NSP13/Helicase | V5921L | non-synonymous | 32 | conservative | Reported | |
| **C18060T** | 1 | 276 | Orf1ab | NSP14/NSP11 | L5932L | synonymous | NA | NA | Reported | |
| **G18197A** | 1 | 4 | Orf1ab | NSP14/NSP11 | R5978K | non-synonymous | 26 | conservative | New | |
| **G18412T** | 1 | 557 | Orf1ab | NSP14/NSP11 | V6050F | non-synonymous | 50 | conservative | Reported | |
| **G18462A** | 2 | 53, 491 | Orf1ab | NSP14/NSP11 | P6066P | synonymous | NA | NA | Reported | |
| **T18476C** | 1 | 43 | Orf1ab | NSP14/NSP11 | F6071S | non-synonymous | 155 | radical | New | |
| **C18486T** | 1 | 29 | Orf1ab | NSP14/NSP11 | L6074L | synonymous | NA | NA | Reported | |
| **C18687T** | 1 | 19 | Orf1ab | NSP14/NSP11 | C6141C | synonymous | NA | NA | Reported | |
| **C18705T** | 1 | 496 | Orf1ab | NSP14/NSP11 | D6147D | synonymous | NA | NA | Reported | |
| **A18774G** | 2 | 42, 73 | Orf1ab | NSP14/NSP11 | Q6170Q | synonymous | NA | NA | Reported | |
| **G18782T** | 1 | 3 | Orf1ab | NSP14/NSP11 | G6173V | non-synonymous | 109 | radical | Reported | |
| **T18783del (7)** | 1 | 4 | Orf1ab | NSP14/NSP11 | G6173G | synonymous | NA | radical | Reported | (7) GT18782G or T18783-deletion causes |
| **C18807T** | 1 | 455 | Orf1ab | NSP14/NSP11 | N6181N | synonymous | NA | NA | Reported | |
| **C18877T** | 3 | 7-470 | Orf1ab | NSP14/NSP11 | L6205L | synonymous | NA | NA | Reported | |
| **C18998T** | 15 | 66-508 | Orf1ab | NSP14/NSP11 | A6245V | non-synonymous | 64 | conservative | Reported | |
| **T19025A** | 1 | 26 | Orf1ab | NSP14/NSP11 | L6254H | non-synonymous | 99 | conservative | New | |
| **A19073G** | 1 | 464 | Orf1ab | NSP14/NSP11 | D6270G | non-synonymous | 94 | conservative | Reported | |
| **T19147C** | 1 | 445 | Orf1ab | NSP14/NSP11 | Y6295H | non-synonymous | 83 | conservative | New | |
| **T19181A** | 1 | 10 | Orf1ab | NSP14/NSP11 | V6306E | non-synonymous | 121 | radical | New | |
| **C19698T** | 1 | 469 | Orf1ab | NSP15/NSP11 | I6478I | synonymous | NA | NA | New | |
| **C19763T** | 1 | 476 | Orf1ab | NSP15/NSP11 | T6500I | non-synonymous | 89 | conservative | Reported | |
| **T19985C** | 1 | 38 | Orf1ab | NSP15/EndoRNAse | F6574S | non-synonymous | 155 | radical | New | |
| **G20005A** | 1 | 493 | Orf1ab | NSP15/EndoRNAse | G6581S | non-synonymous | 56 | conservative | Reported | |
| **A20755C** | 2 | 278, 467 | Orf1ab | NSP16/NSP13 | S6831R | non-synonymous | 110 | radical | Reported | |
| **C20843T** | 1 | 11 | Orf1ab | NSP16/NSP13 | P6860L | non-synonymous | 98 | conservative | Reported | |
| **T20852C** | 1 | 2 | Orf1ab | NSP16/NSP13 | M6863T | non-synonymous | 81 | conservative | New | |
| **A20992T** | 1 | 21 | Orf1ab | NSP16/NSP13 | I6910F | non-synonymous | 21 | conservative | New | |
| **C21017T** | 1 | 469 | Orf1ab | NSP16/NSP13 | T6918I | non-synonymous | 89 | conservative | Reported | |
| **A21137T** | 2 | 4, 38 | Orf1ab | NSP16/NSP13 | K6958M | non-synonymous | 95 | conservative | Reported | |
| **G21305A** | 1 | 46 | Orf1ab | NSP16/NSP13 | R7014H | non-synonymous | 29 | conservative | Reported | |
| **T21368C *** | 1 | 4 | Orf1ab | NSP16/NSP13 | I7035T | non-synonymous | 89 | conservative | New | |
| **G21485T** | 1 | 434 | Orf1ab | NSP16/NSP13 | S7074I | non-synonymous | 142 | conservative | New | |
| **T21524del (8)** | 1 | 72 | Orf1ab | NSP16/NSP13 | V7087V | synonymous | NA | radical | New | (8) GT21523G or T21524-deletion causes |
| **C21621T** | 1 | 135 | S gene | Spike | T20I | non-synonymous | 89 | conservative | Reported | |
| **C21716G** | 1 | 3 | S gene | Spike | Q52E | non-synonymous | 29 | conservative | New | |
| **C21789T** | 1 | 484 | S gene | Spike | T76I | non-synonymous | 89 | conservative | Reported | |
| **G21800A** | 1 | 180 | S gene | Spike | D80N | non-synonymous | 23 | conservative | Reported | |
| **C21805del (9)** | 1 | 22 | S gene | Spike | N81N | synonymous | NA | radical | New | (9) AC21804A or C21805-deletion causes |
| **C21885T** | 1 | 6 | S gene | Spike | T108I | non-synonymous | 89 | conservative | New | |
| **C21930T** | 1 | 11 | S gene | Spike | A123V | non-synonymous | 64 | conservative | Reported | |
| **A21949C** | 1 | 3 | S gene | Spike | K129N | non-synonymous | 94 | conservative | New | |
| **TTTTGGGTGTTTA21981T * (10)** | 1 | 42 | S gene | Spike | F140F | synonymous | NA | radical | New | (10) Deletion of the TTTTGGGTGTTA |
| **T22089C** | 1 | 385 | S gene | Spike | L176P | non-synonymous | 98 | conservative | New | |
| **T22191C** | 1 | 475 | S gene | Spike | I210T | non-synonymous | 89 | conservative | Reported | |
| **C22281T** | 1 | 490 | S gene | Spike | T240I | non-synonymous | 89 | conservative | Reported | |
| **G22344C** | 1 | 18 | S gene | Spike | G261A | non-synonymous | 60 | conservative | Reported | |
| **C22419T** | 1 | 4 | S gene | Spike | T286I | non-synonymous | 89 | conservative | New | |
| **G22599A** | 1 | 449 | S gene | Spike/RBD | R356K | non-synonymous | 26 | conservative | Reported | |
| **T22721C** | 1 | 10 | S gene | Spike/RBD | L387L | synonymous | NA | NA | New | |
| **G22918T** | 1 | 447 | S gene | Spike/RBD/ACE2 | L452L | synonymous | NA | NA | Reported | |
| **T23020C** | 1 | 11 | S gene | Spike/RBD/ACE2 | F486F | synonymous | NA | NA | New | |
| **T23094A** | 1 | 5 | S gene | Spike/RBD | V511E | non-synonymous | 121 | radical | New | |
| **T23119C** | 1 | 434 | S gene | Spike/RBD | H519H | synonymous | NA | NA | New | |
| **T23197del (11)** | 1 | 4 | S gene | Spike/RBD | G545G | synonymous | na | radical | New | (11) GT23196G or T23197-deletion causes |
| **T23227C** | 1 | 494 | S gene | Spike/RBD | S555S | synonymous | na | NA | New | |
| **C23286T** | 1 | 14 | S gene | Spike/RBD | A575V | non-synonymous | 64 | conservative | New | |
| **A23403G** | 65 | 8-478 | S gene | Spike | D614G | non-synonymous | 94 | conservative | Reported | |
| **G23608A** | 1 | 13 | S gene | Spike | R682R | synonymous | NA | NA | Reported | |
| **A23723T** | 1 | 3 | S gene | Spike/S2 | S721C | non-synonymous | 112 | radical | New | |
| **G24004T** | 1 | 12 | S gene | Spike/S2 | K814N | non-synonymous | 94 | conservative | New | |
| **G24087A** | 1 | 9 | S gene | Spike/S2/S2'/FP2 | G842D | non-synonymous | 94 | conservative | New | |
| **G24200C** | 1 | 48 | S gene | Spike/S2/S2' | G880R | non-synonymous | 125 | radical | New | |
| **C24213T** | 1 | 4 | S gene | Spike/S2/S2' | S884F | non-synonymous | 155 | radical | Reported | |
| **C24270T** | 1 | 16 | S gene | Spike/S2/S2' | A903V | non-synonymous | 64 | conservative | New | |
| **T24283A** | 1 | 9 | S gene | Spike/S2/S2' | N907K | non-synonymous | 94 | conservative | New | |
| **T24552C** | 1 | 4 | S gene | Spike/S2/S2' | I997T | non-synonymous | 89 | conservative | New | |
| **T24873A** | 1 | 14 | S gene | Spike/S2/S2' | V1104E | non-synonymous | 121 | radical | New | |
| **C24878A** | 1 | 14 | S gene | Spike/S2/S2' | Q1106K | non-synonymous | 53 | conservative | New | |
| **A25141G** | 1 | 22 | S gene | Spike/S2/S2' | L1193L | synonymous | NA | NA | New | |
| **G25354T** | 1 | 5 | S gene | Spike/S2/S2' | V1264V | synonymous | NA | NA | Reported | |
| **G25471T** | 1 | 270 | Orf3a | Orf3a Protein | D27Y | non-synonymous | 160 | radical | Reported | |
| **G25563T** | 60 | 6-469 | Orf3a | Orf3a Protein | Q57H | non-synonymous | 24 | conservative | Reported | |
| **C25571T** | 1 | 12 | Orf3a | Orf3a Protein | S60F | non-synonymous | 155 | radical | Reported | |
| **C25587T** | 2 | 456, 466 | Orf3a | Orf3a Protein | L65L | synonymous | NA | NA | Reported | |
| **C25613T** | 1 | 374 | Orf3a | Orf3a Protein | S74F | non-synonymous | 155 | radical | Reported | |
| **G25690T** | 1 | 481 | Orf3a | Orf3a Protein | G100C | non-synonymous | 159 | radical | Reported | |
| **C25693T** | 1 | 7 | Orf3a | Orf3a Protein | L101F | non-synonymous | 22 | conservative | Reported | |
| **G25793A** | 1 | 219 | Orf3a | Orf3a Protein | R134H | non-synonymous | 29 | conservative | Reported | |
| **A26057C** | 1 | 8 | Orf3a | Orf3a Protein | D222A | non-synonymous | 126 | radical | Reported | |
| **G26144T** | 2 | 414, 476 | Orf3a | Orf3a Protein | G251V | non-synonymous | 109 | radical | Reported | |
| **G26188T (12)** | 1 | 452 | Orf3a | Orf3a Protein | E266-STOP | non-synonymous | NA | radical | New | (12) The codon creates a truncated |
| **C26425T** | 1 | 30 | E gene | Envelope Protein | R61C | non-synonymous | 180 | radical | Reported | |
| **C26509T** | 1 | 246 | E/M Linking | NA | NA | NA | NA | NA | New | |
| **G26581C** | 1 | 19 | M gene | Membrane/Matrix Protein | W20S | non-synonymous | 177 | radical | New | |
| **G26992A** | 1 | 7 | M gene | Membrane/Matrix Protein | G157E | non-synonymous | 98 | conservative | New | |
| **G27238T (13)** | 1 | 44 | Orf6 | Orf6 Protein | E13-STOP | non-synonymous | NA | radical | Reported | (13) The stop codon creates a truncated |
| **G27238A** | 1 | 30 | Orf6 | Orf6 Protein | E13K | non-synonymous | 98 | conservative | Reported | |
| **G27260T** | 1 | 7 | Orf6 | Orf6 Protein | R20M | non-synonymous | 91 | conservative | New | |
| **G27506T** | 1 | 417 | Orf7a | Orf7a Protein | G38V | non-synonymous | 109 | radical | Reported | |
| **A27669T** | 1 | 13 | Orf7a | Orf7a Protein | E92D | non-synonymous | 45 | conservative | Reported | |
| **G27915A** | 1 | 286 | Orf8 | Orf8 Protein | G8R | non-synonymous | 125 | radical | Reported | |
| **C27964T** | 7 | 11-472 | Orf8 | Orf8 Protein | S24L | non-synonymous | 145 | radical | Reported | |
| **A28058T** | 1 | 31 | Orf8 | Orf8 Protein | A55A | synonymous | NA | NA | New | |
| **G28079T** | 1 | 453 | Orf8 | Orf8 Protein | V62V | synonymous | NA | NA | Reported | |
| **C28093T** | 1 | 187 | Orf8 | Orf8 Protein | S67F | non-synonymous | 155 | radical | Reported | |
| **T28144C** | 1 | 86 | Orf8 | Orf8 Protein | L84S | non-synonymous | 145 | radical | Reported | |
| **C28253T** | 1 | 140 | Orf8 | Orf8 Protein | F120F | synonymous | NA | NA | Reported | |
| **G28326T** | 1 | 440 | N gene | Nucleocapsid | G18V | non-synonymous | 109 | radical | Reported | |
| **C28435A** | 1 | 433 | N gene | Nucleocapsid | T54T | synonymous | NA | NA | Reported | |
| **G28559del (14)** | 1 | 21 | N gene | Nucleocapsid | G96V | non-synonymous | 109 | radical | New | (14) The TG28558Tor G28559-deletion |
| **G28560C** | 1 | 22 | N gene | Nucleocapsid | G96A | non-synonymous | 60 | conservative | New | |
| **G28614A** | 1 | 9 | N gene | Nucleocapsid | G114E | non-synonymous | 98 | conservative | New | |
| **C28657T** | 1 | 477 | N gene | Nucleocapsid | D128D | synonymous | NA | NA | Reported | |
| **C28821A** | 1 | 465 | N gene | Nucleocapsid | S183Y | non-synonymous | 144 | radical | Reported | |
| **G28851T** | 1 | 465 | N gene | Nucleocapsid | S193I | non-synonymous | 142 | radical | Reported | |
| **A28860G** | 1 | 492 | N gene | Nucleocapsid | N196S | non-synonymous | 46 | conservative | Reported | |
| **G28881A** | 2 | 8, 500 | N gene | Nucleocapsid | R203K | non-synonymous | 26 | conservative | Reported | |
| **G28882A** | 2 | 8,497 | N gene | Nucleocapsid | R203R | synonymous | NA | NA | Reported | Others reported it as GGG28881AAC |
| **G28883C** | 2 | 8, 497 | N gene | Nucleocapsid | G204R | non-synonymous | 125 | radical | Reported | Others reported it as GGG28881AAC |
| **C28887T** | 2 | 466, 482 | N gene | Nucleocapsid | T205I | non-synonymous | 89 | conservative | Reported | Others reported it as GGG28881AAC |
| **G28914T** | 1 | 25 | N gene | Nucleocapsid | G214V | non-synonymous | 109 | radical | New | |
| **C29028T** | 1 | 419 | N gene | Nucleocapsid | A252V | non-synonymous | 64 | conservative | New | |
| **C29280T** | 1 | 473 | N gene | Nucleocapsid | A336V | non-synonymous | 64 | conservative | New | |
| **G29402T** | 2 | 410, 469 | N gene | Nucleocapsid | D377Y | non-synonymous | 160 | radical | Reported | |
| **C29445T** | 1 | 467 | N gene | Nucleocapsid | T391I | non-synonymous | 89 | conservative | Reported | |
| **G29540A** | 16 | 8-459 | N/Orf10 Linking | NA | extragenic | NA | NA | NA | Reported | |
| **C29686T** | 1 | 437 | 3'UTR | NA | extragenic | NA | NA | NA | Reported | |
| **G29779T** | 1 | 13 | 3'UTR | NA | extragenic | NA | NA | NA | Reported | |

**Supplemental Table 7. Validation data of SARS-CoV-2 genetic variants, *Prevotella spp.* and the primer sets.** (A) To validate the variant calling of our assay and software, we selected 3 genetic regions containing mutations identified by our protocol. The genetic regions were PCR amplified using specifically designed primers followed by Sanger sequencing. The mutations for validation were selected using the following considerations: read depth, location, synonymy, Grantham score and AA (aminoacid replacement). Most new mutations (not yet reported by GISAID by November 11, 2020) were observed in the ORF1ab and S gene regions. One of the specific TN mutations (T6394C) was not reported by GISAID, and also included in the selection from the ORF1ab region. The primer design considered nearby mutations to the selected variants in order to maximize the validation efficiency of the sequencing reaction. The mutations that were primarily selected are highlighted in bold. (B) Metagenomic validation of *Prevotella spp.*, PCR amplification conditions and primer set.

| **A. Validation of SARS-CoV-2 genetic variants and primer sets.** | | | | | | | | |
|---|---|---|---|---|---|---|---|---|
| Mutations | Read Depth | Location | Protein | Synonymy | Grantham | AA change | Forward Primer | Reverse Primer |
| A6295G | 19, 482 | ORF1ab | NSP3 | non-synonymous | 10 | conservative | 5'-ATG GTG ATG TGG TGG CTA TTG A-3' | 5'-GAT CTG TGT GGC CAA CCT CT-3' |
| **T6394C** | 20-473 | ORF1ab | NSP3 | synonymous | NA | NA | | |
| **G15906T** | 11, 447 | ORF1ab | NSP12/RNAde | non-synonymous | 24 | conservative | 5'-GTC AAG CTG TCA CGG CCA AT-3' | 5'-AAC CTG GAG CAT TGC AAA CA-3' |
| C15924T | 11-481 | ORF1ab | NSP12/RNAde [illegible] | synonymous | NA | NA | | |
| **G21485T** | 434 | ORF1ab | NSP16/NSP13 | non-synonymous | 142 | radical | 5'-TAT CTT GGC AAA CCA CGC GA-3' | 5'-CCC TGT TTT CCT TCA AGG TCC-3' |
| C21621T | 135 | S gene | Spike Protein | non-synonymous | 89 | conservative | | |
| **B. Prevotella 16s rDNA primer set.** | | | | | | | | |
| *Prevotella spp.* | PCR amplification was performed with 1x KAPA-HiFi polymerase Ready Mix, 250nM of each primer, 1 ng of DNA input with the following cycling conditions: 95°C for 3 minutes, 35 cycles of 98°C for 20 seconds, 58°C for 15 seconds, 72°C for 1 minute; 72°C for 5 minutes. | | | | | | 5'-GGG ATG CGT CYG ATT AGB YWG YH-3' | 5'-SCY TAG GYC GHY CCT YSC GGT-3' |

**Supplemental Table 8. Relevant Functional Metagenomic Profiles Obtained from HUMAnN2 Analysis.** We detected a total of 434 functional pathways in our data set, of which 4 were found to be increased in CVN samples and 3 in CVP samples. Additionally we have listed the functional pathways associated to those bacterial species significantly increased in CVP samples with LEfSE analysis.

| Pathway ID MetaCyc | Pathway name | Taxa | COVID-Status |
|---|---|---|---|
| PWY-6168 | flavin biosynthesis III (fungi) | Bacillus subtilis | CVP |
| PWY-6168 | flavin biosynthesis III (fungi) | Dialister micraerophilus | CVP |
| PWY-6168 | flavin biosynthesis III (fungi) | Escherichia coli | CVP |
| PWY-6168 | flavin biosynthesis III (fungi) | Kingella denitrificans | CVP |
| PWY-6168 | flavin biosynthesis III (fungi) | Saccharomyces cerevisiae | CVP |
| PWY-6168 | flavin biosynthesis III (fungi) | unclassified | CVP |
| PWY-6936 | seleno-amino acid biosynthesis | Actinomyces sp HPA0247 | CVP |
| PWY-6936 | seleno-amino acid biosynthesis | Actinomyces sp ICM39 | CVP |
| PWY-6936 | seleno-amino acid biosynthesis | Actinomyces sp ICM47 | CVP |
| PWY-6936 | seleno-amino acid biosynthesis | Bacillus subtilis | CVP |
| PWY-6936 | seleno-amino acid biosynthesis | Campylobacter showae | CVP |
| PWY-6936 | seleno-amino acid biosynthesis | Corynebacterium propinquum | CVP |
| PWY-6936 | seleno-amino acid biosynthesis | Escherichia coli | CVP |
| PWY-6936 | seleno-amino acid biosynthesis | Granulicatella adiacens | CVP |
| PWY-6936 | seleno-amino acid biosynthesis | Haemophilus haemolyticus | CVP |
| PWY-6936 | seleno-amino acid biosynthesis | Haemophilus parahaemolyticus | CVP |
| PWY-6936 | seleno-amino acid biosynthesis | Haemophilus parainfluenzae | CVP |
| PWY-6936 | seleno-amino acid biosynthesis | Haemophilus pittmaniae | CVP |
| PWY-6936 | seleno-amino acid biosynthesis | Haemophilus sputorum | CVP |
| PWY-6936 | seleno-amino acid biosynthesis | Kingella denitrificans | CVP |
| PWY-6936 | seleno-amino acid biosynthesis | Lachnospiraceae bacterium ICM 7 | CVP |
| PWY-6936 | seleno-amino acid biosynthesis | Lautropia mirabilis | CVP |
| PWY-6936 | seleno-amino acid biosynthesis | Neisseria flavescens | CVP |
| PWY-6936 | seleno-amino acid biosynthesis | Neisseria macacae | CVP |
| PWY-6936 | seleno-amino acid biosynthesis | Neisseria sicca | CVP |
| PWY-6936 | seleno-amino acid biosynthesis | Neisseria sp oral taxon 14 | CVP |
| PWY-6936 | seleno-amino acid biosynthesis | Prevotella multiformis | CVP |
| PWY-6936 | seleno-amino acid biosynthesis | Pseudomonas synxantha | CVP |
| PWY-6936 | seleno-amino acid biosynthesis | Rothia dentocariosa | CVP |
| PWY-6936 | seleno-amino acid biosynthesis | Rothia mucilaginosa | CVP |
| PWY-6936 | seleno-amino acid biosynthesis | Saccharomyces cerevisiae | CVP |
| PWY-6936 | seleno-amino acid biosynthesis | Salmonella enterica | CVP |
| PWY-6936 | seleno-amino acid biosynthesis | Selenomonas flueggei | CVP |
| PWY-6936 | seleno-amino acid biosynthesis | Selenomonas noxia | CVP |
| PWY-6936 | seleno-amino acid biosynthesis | Selenomonas sputigena | CVP |
| PWY-6936 | seleno-amino acid biosynthesis | Staphylococcus aureus | CVP |
| PWY-6936 | seleno-amino acid biosynthesis | Staphylococcus epidermidis | CVP |
| PWY-6936 | seleno-amino acid biosynthesis | Streptococcus infantis | CVP |
| PWY-6936 | seleno-amino acid biosynthesis | Streptococcus mitis oralis pneumoniae | CVP |
| PWY-6936 | seleno-amino acid biosynthesis | Streptococcus pyogenes | CVP |
| PWY-6936 | seleno-amino acid biosynthesis | Streptococcus sanguinis | CVP |
| PWY-6936 | seleno-amino acid biosynthesis | unclassified | CVP |
| PWY66-399 | gluconeogenesis | unclassified | CVP |
| UDPNAGSYN-PWY | UDP-N-acetyl-D-glucosamine biosynthesis | Bifidobacterium dentium | CVN |
| UDPNAGSYN-PWY | UDP-N-acetyl-D-glucosamine biosynthesis | Bifidobacterium longum | CVN |
| UDPNAGSYN-PWY | UDP-N-acetyl-D-glucosamine biosynthesis | Campylobacter concisus | CVN |
| UDPNAGSYN-PWY | UDP-N-acetyl-D-glucosamine biosynthesis | Capnocytophaga granulosa | CVN |
| UDPNAGSYN-PWY | UDP-N-acetyl-D-glucosamine biosynthesis | Escherichia coli | CVN |
| UDPNAGSYN-PWY | UDP-N-acetyl-D-glucosamine biosynthesis | Fusobacterium nucleatum | CVN |
| UDPNAGSYN-PWY | UDP-N-acetyl-D-glucosamine biosynthesis | Fusobacterium periodonticum | CVN |
| UDPNAGSYN-PWY | UDP-N-acetyl-D-glucosamine biosynthesis | Haemophilus haemolyticus | CVN |
| UDPNAGSYN-PWY | UDP-N-acetyl-D-glucosamine biosynthesis | Haemophilus influenzae | CVN |
| UDPNAGSYN-PWY | UDP-N-acetyl-D-glucosamine biosynthesis | Lactobacillus fermentum | CVN |
| UDPNAGSYN-PWY | UDP-N-acetyl-D-glucosamine biosynthesis | Listeria monocytogenes | CVN |
| UDPNAGSYN-PWY | UDP-N-acetyl-D-glucosamine biosynthesis | Neisseria meningitidis | CVN |
| UDPNAGSYN-PWY | UDP-N-acetyl-D-glucosamine biosynthesis | Neisseria subflava | CVN |
| UDPNAGSYN-PWY | UDP-N-acetyl-D-glucosamine biosynthesis | Pseudomonas aeruginosa | CVN |
| UDPNAGSYN-PWY | UDP-N-acetyl-D-glucosamine biosynthesis | Pseudomonas synxantha | CVN |
| UDPNAGSYN-PWY | UDP-N-acetyl-D-glucosamine biosynthesis | Salmonella enterica | CVN |
| UDPNAGSYN-PWY | UDP-N-acetyl-D-glucosamine biosynthesis | Staphylococcus aureus | CVN |
| UDPNAGSYN-PWY | UDP-N-acetyl-D-glucosamine biosynthesis | Staphylococcus epidermidis | CVN |
| UDPNAGSYN-PWY | UDP-N-acetyl-D-glucosamine biosynthesis | Streptococcus anginosus | CVN |
| UDPNAGSYN-PWY | UDP-N-acetyl-D-glucosamine biosynthesis | Streptococcus constellatus | CVN |
| UDPNAGSYN-PWY | UDP-N-acetyl-D-glucosamine biosynthesis | Streptococcus cristatus | CVN |
| UDPNAGSYN-PWY | UDP-N-acetyl-D-glucosamine biosynthesis | Streptococcus gordonii | CVN |
| UDPNAGSYN-PWY | UDP-N-acetyl-D-glucosamine biosynthesis | Streptococcus infantis | CVN |
| UDPNAGSYN-PWY | UDP-N-acetyl-D-glucosamine biosynthesis | Streptococcus intermedius | CVN |
| UDPNAGSYN-PWY | UDP-N-acetyl-D-glucosamine biosynthesis | Streptococcus mitis oralis pneumoniae | CVN |
| UDPNAGSYN-PWY | UDP-N-acetyl-D-glucosamine biosynthesis | Streptococcus oligofermentans | CVN |
| UDPNAGSYN-PWY | UDP-N-acetyl-D-glucosamine biosynthesis | Streptococcus pseudopneumoniae | CVN |
| UDPNAGSYN-PWY | UDP-N-acetyl-D-glucosamine biosynthesis | Streptococcus pyogenes | CVN |
| UDPNAGSYN-PWY | UDP-N-acetyl-D-glucosamine biosynthesis | Streptococcus salivarius | CVN |
| UDPNAGSYN-PWY | UDP-N-acetyl-D-glucosamine biosynthesis | Streptococcus sanguinis | CVN |
| UDPNAGSYN-PWY | UDP-N-acetyl-D-glucosamine biosynthesis | Streptococcus thermophilus | CVN |
| UDPNAGSYN-PWY | UDP-N-acetyl-D-glucosamine biosynthesis | Streptococcus tigurinus | CVN |
| UDPNAGSYN-PWY | UDP-N-acetyl-D-glucosamine biosynthesis | unclassified | CVN |
| PWY-5030 | L-histidine degradation | Fusobacterium nucleatum | CVN |
| PWY-5030 | L-histidine degradation | Streptococcus gordonii | CVN |
| PWY-5030 | L-histidine degradation | Streptococcus parasanguinis | CVN |
| PWY-5030 | L-histidine degradation | Streptococcus sanguinis | CVN |
| PWY-5030 | L-histidine degradation | unclassified | CVN |
| MET-SAM-PWY | superpathway of S-adenosyl-L-methionine biosynthesis | Escherichia coli | CVN |
| MET-SAM-PWY | superpathway of S-adenosyl-L-methionine biosynthesis | Oribacterium sinus | CVN |
| MET-SAM-PWY | superpathway of S-adenosyl-L-methionine biosynthesis | Salmonella enterica | CVN |
| MET-SAM-PWY | superpathway of S-adenosyl-L-methionine biosynthesis | Selenomonas flueggei | CVN |
| MET-SAM-PWY | superpathway of S-adenosyl-L-methionine biosynthesis | Streptococcus infantis | CVN |
| MET-SAM-PWY | superpathway of S-adenosyl-L-methionine biosynthesis | Streptococcus mitis oralis pneumoniae | CVN |
| MET-SAM-PWY | superpathway of S-adenosyl-L-methionine biosynthesis | Streptococcus sanguinis | CVN |
| MET-SAM-PWY | superpathway of S-adenosyl-L-methionine biosynthesis | unclassified | CVN |
| METSYN-PWY | L-homoserine and L-methionine biosynthesis | Escherichia coli | CVN |
| METSYN-PWY | L-homoserine and L-methionine biosynthesis | Oribacterium sinus | CVN |
| METSYN-PWY | L-homoserine and L-methionine biosynthesis | Salmonella enterica | CVN |
| METSYN-PWY | L-homoserine and L-methionine biosynthesis | Selenomonas flueggei | CVN |
| METSYN-PWY | L-homoserine and L-methionine biosynthesis | Streptococcus infantis | CVN |
| METSYN-PWY | L-homoserine and L-methionine biosynthesis | Streptococcus mitis oralis pneumoniae | CVN |
| METSYN-PWY | L-homoserine and L-methionine biosynthesis | Streptococcus sanguinis | CVN |
| METSYN-PWY | L-homoserine and L-methionine biosynthesis | unclassified | CVN |
| PWY-7219 | adenosine ribonucleotides de novo biosynthesis | Actinomyces graevenitzii | NA |
| PWY-5100 | pyruvate fermentation to acetate and lactate II | Actinomyces graevenitzii | NA |
| THRESYN-PWY | superpathway of L-threonine biosynthesis | Actinomyces graevenitzii | NA |
| PWY-6151 | S-adenosyl-L-methionine cycle I | Actinomyces graevenitzii | NA |
| PWY-7221 | guanosine ribonucleotides de novo biosynthesis | Actinomyces graevenitzii | NA |

| | | | |
|---|---|---|---|
| PWY-7111 | pyruvate fermentation to isobutanol (engineered) | Actinomyces graevenitzii | NA |
| VALSYN-PWY | L-valine biosynthesis | Actinomyces graevenitzii | NA |
| PWY-6122 | 5-aminoimidazole ribonucleotide biosynthesis II | Actinomyces graevenitzii | NA |
| PWY-6277 | superpathway of 5-aminoimidazole ribonucleotide biosyn | Actinomyces graevenitzii | NA |
| PWY-6121 | 5-aminoimidazole ribonucleotide biosynthesis I | Actinomyces graevenitzii | NA |
| PWY-5188 | tetrapyrrole biosynthesis I (from glutamate) | Actinomyces graevenitzii | NA |
| PWY-7197 | pyrimidine deoxyribonucleotide phosphorylation | Actinomyces graevenitzii | NA |
| PANTO-PWY | phosphopantothenate biosynthesis I | Actinomyces graevenitzii | NA |
| PWY-7208 | superpathway of pyrimidine nucleobases salvage | Actinomyces graevenitzii | NA |
| PWY-7228 | superpathway of guanosine nucleotides de novo biosynth | Actinomyces graevenitzii | NA |
| PWY-7220 | adenosine deoxyribonucleotides de novo biosynthesis II | Actinomyces graevenitzii | NA |
| PWY-7222 | guanosine deoxyribonucleotides de novo biosynthesis II | Actinomyces graevenitzii | NA |
| PWY-6125 | superpathway of guanosine nucleotides de novo biosynth | Actinomyces graevenitzii | NA |
| PWY-5686 | UMP biosynthesis | Actinomyces graevenitzii | NA |
| NONMEVIPP-PWY | methylerythritol phosphate pathway I | Actinomyces graevenitzii | NA |
| COA-PWY-1 | coenzyme A biosynthesis II (mammalian) | Actinomyces graevenitzii | NA |
| PEPTIDOGLYCANSYN-PWY | peptidoglycan biosynthesis I (meso-diaminopimelate cont | Actinomyces graevenitzii | NA |
| PWY-6386 | UDP-N-acetylmuramoyl-pentapeptide biosynthesis II (lysir | Actinomyces graevenitzii | NA |
| PWY-6387 | UDP-N-acetylmuramoyl-pentapeptide biosynthesis I (meso | Actinomyces graevenitzii | NA |
| GALACTUROCAT-PWY | D-galacturonate degradation I | Prevotella salivae | NA |
| PWY-5989 | stearate biosynthesis II (bacteria and plants) | Prevotella salivae | NA |
| PWY-5695 | urate biosynthesis/inosine 5'-phosphate degradation | Prevotella salivae | NA |
| PWY-2942 | L-lysine biosynthesis III | Prevotella salivae | NA |
| PWY-7663 | gondoate biosynthesis (anaerobic) | Prevotella salivae | NA |
| FASYN-ELONG-PWY | fatty acid elongation -- saturated | Prevotella salivae | NA |
| PWY-1269 | CMP-3-deoxy-D-manno-octulosonate biosynthesis I | Prevotella salivae | NA |
| COBALSYN-PWY | adenosylcobalamin salvage from cobinamide I | Prevotella salivae | NA |
| PWY-6609 | adenine and adenosine salvage III | Prevotella salivae | NA |
| PWY-5973 | cis-vaccenate biosynthesis | Prevotella salivae | NA |
| PWY-6282 | palmitoleate biosynthesis I (from (5Z)-dodec-5-enoate) | Prevotella salivae | NA |
| PWY-7664 | oleate biosynthesis IV (anaerobic) | Prevotella salivae | NA |
| PWY-6386 | UDP-N-acetylmuramoyl-pentapeptide biosynthesis II (lysir | Prevotella salivae | NA |
| PWY-5097 | L-lysine biosynthesis VI | Prevotella salivae | NA |
| PWY0-862 | (5Z)-dodec-5-enoate biosynthesis | Prevotella salivae | NA |
| PEPTIDOGLYCANSYN-PWY | peptidoglycan biosynthesis I (meso-diaminopimelate cont | Prevotella salivae | NA |
| ASPASN-PWY | superpathway of L-aspartate and L-asparagine biosynthes | Prevotella salivae | NA |
| PWY-6387 | UDP-N-acetylmuramoyl-pentapeptide biosynthesis I (meso | Prevotella salivae | NA |
| GALACTUROCAT-PWY | D-galacturonate degradation I | Prevotella salivae | NA |
| PWY-7111 | pyruvate fermentation to isobutanol (engineered) | Prevotella salivae | NA |
| VALSYN-PWY | L-valine biosynthesis | Prevotella salivae | NA |
| PWY-7221 | guanosine ribonucleotides de novo biosynthesis | Prevotella salivae | NA |
| GALACT-GLUCUROCAT-PW | superpathway of hexuronide and hexuronate degradation | Prevotella salivae | NA |
| PWY-5686 | UMP biosynthesis | Prevotella salivae | NA |
| PWY-6700 | queuosine biosynthesis | Prevotella salivae | NA |
| DTDPRHAMSYN-PWY | dTDP-L-rhamnose biosynthesis I | Prevotella salivae | NA |
| NONMEVIPP-PWY | methylerythritol phosphate pathway I | Prevotella salivae | NA |
| PWY-7242 | D-fructuronate degradation | Prevotella salivae | NA |
| PWY-6163 | chorismate biosynthesis from 3-dehydroquinate | Prevotella salivae | NA |
| GLUCUROCAT-PWY | superpathway of β-D-glucuronide and D-glucuronate | Prevotella salivae | NA |
| GALACT-GLUCUROCAT-PW | superpathway of hexuronide and hexuronate degradation | Prevotella salivae | NA |
| PWY-5667 | CDP-diacylglycerol biosynthesis I | Prevotella salivae | NA |
| PWY0-1319 | CDP-diacylglycerol biosynthesis II | Prevotella salivae | NA |
| COA-PWY | coenzyme A biosynthesis I | Prevotella salivae | NA |
| PWY-6147 | 6-hydroxymethyl-dihydropterin diphosphate biosynthesis | Prevotella salivae | NA |
| PWY-6507 | 4-deoxy-L-threo-hex-4-enopyranuronate degradation | Prevotella salivae | NA |
| PWY-7219 | adenosine ribonucleotides de novo biosynthesis | Prevotella salivae | NA |
| PWY-7199 | pyrimidine deoxyribonucleosides salvage | Prevotella salivae | NA |
| PWY-6507 | 4-deoxy-L-threo-hex-4-enopyranuronate degradation | Prevotella salivae | NA |
| PWY-7242 | D-fructuronate degradation | Prevotella salivae | NA |
| GLUCUROCAT-PWY | superpathway of β-D-glucuronide and D-glucuronate | Prevotella salivae | NA |
| PWY-6151 | S-adenosyl-L-methionine cycle I | Prevotella salivae | NA |
| PWY-6124 | inosine-5'-phosphate biosynthesis II | Prevotella salivae | NA |
| PWY-6123 | inosine-5'-phosphate biosynthesis I | Prevotella salivae | NA |
| COA-PWY-1 | coenzyme A biosynthesis II (mammalian) | Prevotella salivae | NA |
| PWY-7111 | pyruvate fermentation to isobutanol (engineered) | Megasphaera micronuciformis | NA |
| VALSYN-PWY | L-valine biosynthesis | Megasphaera micronuciformis | NA |
| PWY-6163 | chorismate biosynthesis from 3-dehydroquinate | Megasphaera micronuciformis | NA |
| PWY-7221 | guanosine ribonucleotides de novo biosynthesis | Megasphaera micronuciformis | NA |
| PWY-5695 | urate biosynthesis/inosine 5'-phosphate degradation | Megasphaera micronuciformis | NA |
| COA-PWY | coenzyme A biosynthesis I | Megasphaera micronuciformis | NA |
| THRESYN-PWY | superpathway of L-threonine biosynthesis | Megasphaera micronuciformis | NA |
| PWY-7219 | adenosine ribonucleotides de novo biosynthesis | Megasphaera micronuciformis | NA |
| PEPTIDOGLYCANSYN-PWY | peptidoglycan biosynthesis I (meso-diaminopimelate cont | Megasphaera micronuciformis | NA |
| PWY-4242 | pantothenate and coenzyme A biosynthesis III | Megasphaera micronuciformis | NA |
| PWY-6608 | guanosine nucleotides degradation III | Megasphaera micronuciformis | NA |
| PWY-6387 | UDP-N-acetylmuramoyl-pentapeptide biosynthesis I (meso | Megasphaera micronuciformis | NA |
| COA-PWY-1 | coenzyme A biosynthesis II (mammalian) | Megasphaera micronuciformis | NA |
| PWY-6897 | thiamin salvage II | Megasphaera micronuciformis | NA |
| PWY-5686 | UMP biosynthesis | Megasphaera micronuciformis | NA |
| PWY-2942 | L-lysine biosynthesis III | Megasphaera micronuciformis | NA |
| PWY-5097 | L-lysine biosynthesis VI | Megasphaera micronuciformis | NA |
| PWY0-1296 | purine ribonucleosides degradation | Megasphaera micronuciformis | NA |
| PWY-7357 | thiamin formation from pyrithiamine and oxythiamine (ye | Megasphaera micronuciformis | NA |
| PWY-6122 | 5-aminoimidazole ribonucleotide biosynthesis II | Megasphaera micronuciformis | NA |
| PWY-6277 | superpathway of 5-aminoimidazole ribonucleotide biosyn | Megasphaera micronuciformis | NA |
| PWY-724 | superpathway of L-lysine, L-threonine and L-methionine b | Megasphaera micronuciformis | NA |
| PWY-6121 | 5-aminoimidazole ribonucleotide biosynthesis I | Megasphaera micronuciformis | NA |
| PWY-5667 | CDP-diacylglycerol biosynthesis I | Megasphaera micronuciformis | NA |
| PWY0-1319 | CDP-diacylglycerol biosynthesis II | Megasphaera micronuciformis | NA |
| PWY-6386 | UDP-N-acetylmuramoyl-pentapeptide biosynthesis II (lysir | Megasphaera micronuciformis | NA |
| PWY-6385 | peptidoglycan biosynthesis III (mycobacteria) | Megasphaera micronuciformis | NA |
| PWY-6700 | queuosine biosynthesis | Megasphaera micronuciformis | NA |
| PWY-6703 | preQ0 biosynthesis | Megasphaera micronuciformis | NA |
| PWY-6147 | 6-hydroxymethyl-dihydropterin diphosphate biosynthesis | Megasphaera micronuciformis | NA |
| PWY-7219 | adenosine ribonucleotides de novo biosynthesis | Atopobium parvulum | NA |
| PWY0-1296 | purine ribonucleosides degradation | Atopobium parvulum | NA |
| DTDPRHAMSYN-PWY | dTDP-L-rhamnose biosynthesis I | Atopobium parvulum | NA |
| PWY-2942 | L-lysine biosynthesis III | Atopobium parvulum | NA |
| PWY-6609 | adenine and adenosine salvage III | Atopobium parvulum | NA |
| PWY-5686 | UMP biosynthesis | Atopobium parvulum | NA |
| PWY-7221 | guanosine ribonucleotides de novo biosynthesis | Atopobium parvulum | NA |
| ASPASN-PWY | superpathway of L-aspartate and L-asparagine biosynthes | Atopobium parvulum | NA |
| PWY-6386 | UDP-N-acetylmuramoyl-pentapeptide biosynthesis II (lysir | Atopobium parvulum | NA |
| PWY0-1586 | peptidoglycan maturation (meso-diaminopimelate contair | Atopobium parvulum | NA |
| COA-PWY-1 | coenzyme A biosynthesis II (mammalian) | Atopobium parvulum | NA |
